# Quantifying the impact of cascade inequalities: a modelling study on the prevention impacts of antiretroviral therapy scale-up in Eswatini

**DOI:** 10.1101/2024.02.16.24302584

**Authors:** Jesse Knight, Huiting Ma, Bheki Sithole, Lungile Khumalo, Linwei Wang, Sheree Schwartz, Laura Muzart, Sindy Matse, Zandile Mnisi, Rupert Kaul, Michael Escobar, Stefan Baral, Sharmistha Mishra

## Abstract

**Background:** Inequalities in the antiretroviral therapy (ART) cascade across subpopulations remain an ongoing challenge in the global HIV response. Eswatini achieved the UNAIDS 95-95-95 targets by 2020, with differentiated programs to minimize inequalities across subpopulations, including for female sex workers (FSW) and their clients. We sought to estimate additional HIV infections expected in Eswatini if cascade scale-up had not been equal, and under which epidemic conditions these inequalities could have the largest influence.

**Methods:** Drawing on population-level and FSW-specific surveys in Eswatini, we developed a compartmental model of heterosexual HIV transmission which included eight subpopulations and four sexual partnership types. We calibrated the model to stratified HIV prevalence, incidence, and ART cascade data. Taking observed cascade scale-up in Eswatini as the basecase — reaching 95-95-95 in the overall population by 2020 — we defined four counterfactual scenarios in which the population overall reached 80-80-90 by 2020, but where FSW, clients, both, or neither were disproportionately left behind, reaching only 60-40-80. We quantified relative additional cumulative HIV infections by 2030 in counterfactual *vs* base-case scenarios. We further estimated linear effects of viral suppression gap among FSW and clients on additional infections by 2030, plus effect modification by FSW/client population sizes, rates of turnover, and HIV prevalence ratios.

**Results:** Compared with the base-case scenario, leaving behind neither FSW nor their clients led to the fewest additional infections by 2030: median (95% credible interval) 14.9 (10.4, 18.4) % *vs* 26.3 (19.7, 33.0) % if both were left behind — a 73 (40, 149) % increase. The effect of lower cascade on additional infections was larger for clients *vs* FSW, and both effects increased with population size and relative HIV incidence.

**Conclusions:** Inequalities in the ART cascade across subpopulations can undermine the anticipated prevention impacts of cascade scale-up. As Eswatini has shown, addressing inequalities in the ART cascade, particularly those that intersect with high transmission risk, could maximize incidence reductions from cascade scale-up.

## 1 Introduction

Early HIV treatment via antiretroviral therapy (ART) is a lifesaving intervention increasing both the quantity and quality of life [1]. A secondary benefit of early ART given Undetectable = Untransmittable (U=U) is that transmission risks are mitigated in serodifferent partnerships [2]. To realize these benefits, massive efforts are underway to achieve the UNAIDS 95-95-95 ART cascade targets [3] — *i*.*e*., to have: 95% diagnosed among people living with HIV, 95% on ART among those diagnosed, and 95% virally suppressed among those on ART. Botswana, Eswatini, Rwanda, Tanzania, and Zimbabwe have already surpassed 95-95-95 nationally [3], and and achieving these targets is expected to help reduce HIV incidence towards elimination.

Yet, there are growing concerns that inequalities in the ART cascade could undermine the population-level prevention impacts of ART anticipated from individual-level and model-based studies [4–6]. Specifically, available data suggest that cascade attainment is often lower among subpopulations at greater risk of HIV acquisition and/or transmission, including key populations, younger men and women, and highly mobile populations [5,7]. These inequalities can be driven by systemic barriers to engagement in care faced by marginalized populations, which intersect with individual, network, and structural determinants of HIV risk, such as economic insecurity, mobility, stigma, discrimination, and criminalization [8–11]. Moreover, cascade data may be lacking entirely for subpopulations experiencing the greatest barriers to care — *i*.*e*., the lowest ART cascades likely remain unmeasured [7].

Numerous transmission modelling studies have sought to estimate the prevention impacts of achieving 90-90-90+ across Sub-Saharan Africa [12,13]. Modelled populations are often stratified by risk, including key populations like female sex workers (FSW) and their clients, to capture important epidemic dynamics related to risk heterogeneity [14]. However, these studies have generally assumed that ART cascade attainment (*i*.*e*., proportions diagnosed, treated, and virally suppressed) or progression (*i*.*e*., rates of diagnosis, treatment initiation, and viral suppression) were equal across modelled subpopulations. For example, among the studies in [13] (see also Box 1), key populations were usually assumed to have equal cascade progression with the population overall, or greater in some scenarios, but never lesser. Thus, the potential influence of intersecting risk and cascade inequalities on ART prevention impacts has not been explored.

We therefore examined the following questions in an illustrative modelling analysis:

1. How are estimates of population-level ART prevention impacts influenced by inequalities in ART cascade across subpopulations?
2. Under which epidemic conditions do such inequalities have the largest influence?

We examined these questions using a deterministic compartmental model of heterosexual HIV transmission in Eswatini, focusing on inequalities related to sex work. Eswatini has the highest national HIV prevalence in the world, and recently surpassed 95-95-95 with minimal inequalities across subpopulations [3,15]. As such, we used observed ART cascade scale-up in Eswatini as a *base case* reflecting exemplary and evidently attainable scale-up, and explored *counterfactual* scenarios in which scale-up was slower, and where specific subpopulations could have been left behind.

## 2 Methods

We constructed a deterministic compartmental model of heterosexual HIV transmission, stratified by subpopulations defined by sex and sexual activity, and health states reflecting HIV natural history and ART cascade of care. The model includes eight subpopulations, including FSW at higher *vs* lower risk, and likewise for clients of FSW, and four partnership types, including main/spousal, casual, repeat sex work, and one-off sex work (Figure 1). We calibrated the model to reflect the HIV epidemic and ART scale-up in Eswatini (*base case*). We then explored *counterfactual* scenarios in which ART cascade was reduced among various combinations of subpopulations, and quantified ART prevention impacts by comparing *base case* and *counterfactual* scenarios.

**Figure 1:**
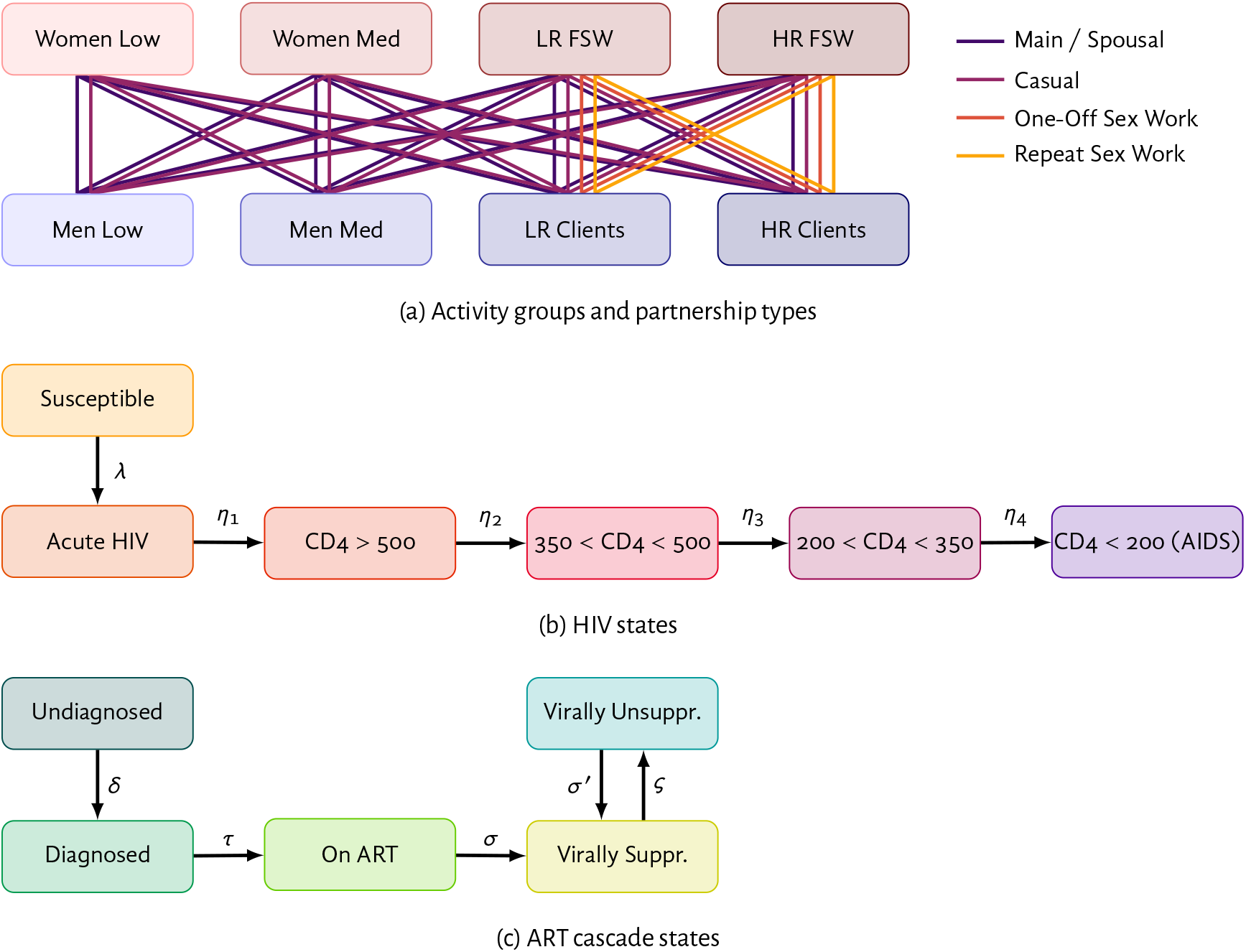
Model structure and transitions Low: lowest activity; Med: medium activity; LR/HR: lower/higher risk; FSW: female sex workers; Clients: of FSW; CD4: CD4+ T-cell count per mm^3^; ART: antiretroviral therapy; rates — *λ*: force of infection; *η*: HIV progression; *δ*: diagnosis; *τ*: ART initiation; *σ*: viral suppression; *σ*^′^: viral re-suppression; *ς*: ART failure / discontinuation; not shown: turnover amongst activity groups in (a).

### 2.1 Model Parameterization & Calibration

Complete details of the model structure, parameterization, and calibration are given in Appendix A.

#### HIV

HIV natural history included acute infection and stages defined by CD4-count. We modelled relative rates of infectiousness by stage as an approximation of viral load [16–18], as well as rates of HIV-attributable mortality by stage [19].

#### Risk heterogeneity

We captured risk heterogeneity through subpopulation-level factors, including subpopulation size, turnover among subpopulations, genital ulcer disease (GUD) prevalence, and rates / types of partnership formation; and partnership-level factors, including assortative mixing, partnership duration, frequency of sex, and levels of condom use. Table 1 summarizes key parameter values and sampling distributions related to risk heterogeneity. To parameterize FSW at higher *vs* lower risk, we analyzed individual-level survey data from Swati FSW in 2011 [20] and 2014 [21] (Appendix A.3.9). We parameterized the remaining subpopulations using reported data from national studies in 2006 [22], 2011 [23], and 2016 [24]. We modelled increasing condom use (Figure B.10), increasing voluntary medical male circumcision (Figure B.9), and decreasing GUD prevalence over time, We did not model other interventions (*e*.*g*., current pre-exposure prophylaxis scale-up) nor non-heterosexual HIV transmission.

**Table 1:** Selected model parameters related to risk heterogeneity.

| Parameter | Stratification | Posterior |  | Ref. |
| --- | --- | --- | --- | --- |
|  |  | Mean | (95% CI) |  |
| Subpopulation size<br>(% of total) | FSW, of women overall | 4.4 | (2.8, 6.7) | § A.3.11.1 |
|  | Clients, of men overall | 10.0 | (4.7, 15.7) | § A.3.11.2 |
|  | HR of FSW / clients overall | 20 | — | § A.3.9 |
| Duration in subpop<br>(mean years) | FSW overall | 3.5 | (2.7, 4.9) | § A.3.12.2 |
|  | Clients overall | 8.9 | (4.9, 15.4) | § A.3.12.2 |
|  | HR sex work and clients | 0.7 | (0.3, 1.2) | § A.3.12.2 |
| Sex work clients<br>per year | HR FSW, one-off sex work | 84 | (53, 126) | § A.3.13.1 |
|  | LR FSW, one-off sex work | 20 | (13, 27) | § A.3.13.1 |
|  | HR FSW, repeat sex work | 50 | (23, 86) | § A.3.13.1 |
|  | LR FSW, repeat sex work | 11 | (6, 21) | § A.3.13.1 |
| Sex work visits<br>per year | HR clients | 90 | (44, 154) | § A.3.13.1 |
|  | LR clients | 44 | (22, 74) | § A.3.13.1 |
| Total partner change<br>rate per year | Medium activity women | 0.99 | (0.72, 1.31) | § A.3.13.2 |
|  | Medium activity men | 0.67 | (0.28, 1.27) | § A.3.13.2 |
|  | Lowest activity women & men | 0.23 | (0.15, 0.35) | § A.3.13.2 |
| Any GUD p12m<br>prevalence (%)<sup>a</sup> | HR FSW | 38 | (25, 53) | § A.3.5.3 |
|  | LR FSW & HR clients | 30 | (20, 41) | § A.3.5.3 |
|  | LR clients | 24 | (15, 33) | § A.3.5.3 |
|  | Medium activity | 18 | (8, 29) | § A.3.5.3 |
|  | Lowest activity | 7 | — | § A.3.5.3 |
| Relative infectiousness | Acute infection | 10.6 | (5.3, 18.1) | § A.3.4.1 |
|  | Any GUD p12m | 2.1 | (1.3, 3.2) | § A.3.4.5 |
| Relative susceptibility | Receptive vaginal sex | 1.7 | (1.2, 2.0) | § A.3.4.2 |
|  | Receptive anal sex | 10.0 | — | § A.3.4.2 |
|  | Any GUD p12m | 3.0 | (1.2, 6.3) | § A.3.4.5 |
| Sex acts per<br>partnership-year | Main/spousal | 77 | (40, 125) | § A.3.14 |
|  | Casual | 120 | (62, 202) | § A.3.14 |
|  | One-off sex work | 12 | — | § A.3.14 |
|  | Repeat sex work | 18 | (12, 30) | § A.3.14 |
| Anal sex acts<br>(% of all acts) | Main/spousal & casual | 11 | (3, 21) | § A.3.14 |
|  | One-off & repeat sex work | 17 | (4, 34) | § A.3.14 |
| Condom use in 2020<br>(% of acts protected) | Main/spousal | 41 | (31, 51) | § A.3.5.2 |
|  | Casual | 70 | (62, 77) | § A.3.5.2 |
|  | One-off sex work | 88 | (78, 95) | § A.3.5.2 |
|  | Repeat sex work | 78 | (64, 87) | § A.3.5.2 |
|  | Anal vs vaginal sex | 74 | (63, 88) | § A.3.5.2 |
| Partnership duration<br>(years) | Main/spousal | 16.4 | (14.6, 18.5) | § A.3.15 |
|  | Casual | 0.78 | (0.45, 1.24) | § A.3.15 |
|  | One-off sex work | 0.83 | — | § A.3.15 |
|  | Repeat sex work | 0.43 | (0.19, 0.83) | § A.3.15 |

Low: lowest activity; Med: medium activity; LR/HR: lower/higher risk; FSW: female sex workers; Clients: of FSW; p12m: past 12 months;
<sup>a</sup> GUD prevalence declines universally after 2010 as described in § A.3.5.3.

#### ART cascade

We modelled rates of HIV diagnosis among people living with HIV as monotonically increasing over time. We defined a base rate for women with low/medium sexual activity, and constant relative rates for men with low/medium sexual activity (*RR* < 1), clients (*RR* < 1), and FSW (*RR* > 1), reflecting increased HIV testing access via antenatal care among women *vs* men, and enhanced screening among FSW [20]. We modelled ART initiation, starting in 2003, similarly except: the relative rate for ART initiation among FSW was *RR* < 1, reflecting specific barriers to uptake and engagement in care [8]; and we defined additional relative rates by CD4 count (*RR* ≤ 1) to reflect historical ART eligibility criteria (Figure A.5) [24]. We modelled viral suppression using a fixed rate for all subpopulations, corresponding to an average of 4 months from ART initiation [25]. We modelled treatment failure / discontinuation with a single monotonically decreasing rate applied to all subpopulations in the base case, reflecting improving treatment success / retention over time [24]. Individuals with treatment failure / discontinuation could re-initiate ART at a fixed rate, reflecting re-engagement in care or detection of treatment failure and initiation of alternative regimens. We modelled rapid CD4 recovery during the first 4 months of ART, followed by slower recovery while virally suppressed [26]. We modelled reduced HIV-attributable mortality among individuals on ART, in addition to mortality benefits of CD4 recovery.

#### Calibration

We calibrated the model to reflect available data from Eswatini on HIV prevalence, HIV incidence, and ART cascade of care overall and stratified by subpopulation where possible (Tables A.6–A.10) [15,20– 24,27] using an adapted version of Incremental Mixture Importance Sampling (IMIS) [28]. Full methodology is given in Appendix A.4, while calibration results are given in Appendix B.1.

### 2.2 Scenarios & Analysis

#### 2.2.1 Objective 1: Influence of ART cascade differences between subpopulations

For Objective 1, We defined the *base case* scenario to reflect observed ART cascade scale-up in Eswatini, reaching 95-95-95 for the population overall by 2020 [15], and 88-98-xx among FSW specifically [27].^1^ Next, we defined four *counterfactual* scenarios in which the overall population cascade reached 80-80-90 by 2020, and where FSW, clients, both, or neither were disproportionately left behind. In these counterfactual scenarios, we altered cascade attainment among FSW, clients, and/or the remaining population (“all others”) by calibrating fixed subpopulation-specific relative rates of: diagnosis (0 ≤ *RR*_*d*_ ≤ 1), treatment initiation (0 ≤ *RR*_*t*_ ≤ 1), and treatment failure / discontinuation (1 ≤ *RR*_*u*_ ≤ 20). When FSW and/or clients were left behind, we calibrated their *RR*s such that these subpopulations attained approximately 60-40-80 by 2020, reflecting some of the lowest cascades recently observed among key populations [7]. By contrast, we calibrated *RR*s for the remaining population such that the Swati population *overall* attained 80-80-90 in all 4 counterfactual scenarios, thus ensuring that a consistent proportion of the population overall attained viral suppression.

Table 2 summarizes these scenarios, while Figure B.17 plots the modelled cascades over time. When cascade rates among FSW and/or clients were unchanged from the base case, the cascade these subpopulations attained could be lower than in the base case due to subpopulation turnover and higher incidence. All cascades continued to increase beyond 2020 due to assumed fixed rates of diagnosis, treatment initiation, and treatment failure / discontinuation thereafter.

**Table 2:** Modelling scenarios for Objective 1 defined by 2020 calibration targets.

| Scenario | ART cascade in 2020<sup>a</sup> |  |  | Re-scaled cascade rates<sup>b</sup> |  |  |
| --- | --- | --- | --- | --- | --- | --- |
|  | FSW | Clients | Overall | FSW | Clients | All Others |
| *Base Case* | 88-98-xx | xx-xx-xx | 95-95-95 | — | — | — |
| *Leave Behind: FSW* | 60-40-80 | — | 80-80-90 | ✓ | ✗ | ✓ |
| *Leave Behind: Clients* | — | 60-40-80 | 80-80-90 | ✗ | ✓ | ✓ |
| *Leave Behind: FSW & Clients* | 60-40-80 | 60-40-80 | 80-80-90 | ✓ | ✓ | ✓ |
| *Leave Behind: Neither* | — | — | 80-80-90 | ✗ | ✗ | ✓ |

<sup>a</sup> Cascade: % diagnosed among PLHIV; % on ART among diagnosed; % virally suppressed among on ART; xx: no data available; <sup>b</sup> Rates of: diagnosis; ART initiation; treatment failure; FSW: female sex workers; Clients: of FSW; All Others: all women and men not involved in sex work. Figure B.17 plots the modelled cascades over time.

We quantified ART prevention impacts via relative cumulative additional infections (CAI) and additional incidence rate (AIR) in the counterfactual scenarios (*k*) *vs* the base case (0), over multiple time horizons up to 2030, starting from *t*0 = 2000:

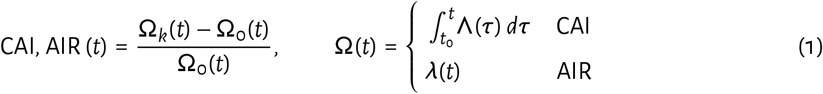

where: Λ denotes absolute numbers of infections per year, and *λ* denotes incidence rate per susceptible per year. For each scenario, we computed these outcomes (CAI and AIR) for each model fit *j*, and reported median (95% credible interval, CI) values across model fits, reflecting uncertainty.

#### 2.2.2 Objective 2: Conditions that maximize the influence of ART cascade differences

For Objective 2, we estimated via linear regression: the effects of lower ART cascade among FSW and clients on relative CAI and AIR, plus potential effect modification by epidemic conditions. The hypothesized causal effects are illustrated as a directed acyclic graph in Figure B.20.

For this regression, we generated 10,000 synthetic samples as follows. We explored a wider range of counterfactual scenarios *vs* Objective 1 by randomly sampling the relative rates for diagnosis and treatment initiation *RR*_*d*_, *RR*_*t*_ ∼ Beta (*α* = 3.45, *β* = 1.85) and treatment failure *RR*_*u*_ ∼ Gamma (*α* = 3.45, *β* = 1.88) for each of: FSW, clients, and the remaining population (9 total values). These sampling distributions had 95% CI: (0.25, 0.95) and (1.5, 15), respectively, and were chosen to obtain cascades in 2020 spanning approximately 60-60-90 through 85-90-95 (Figure B.19). For each of *N*_*f*_ = 1000 model fits, we generated *N*_*k*_ = 10 counterfactual scenarios per fit using Latin hypercube sampling of *RR*s [29], yielding *N*_*f*_ *N*_*k*_ = 10,000 total counterfactual samples for the regression.

For each of these 10,000 samples, we defined relative CAI and AIR by 2030 *vs* the base case, as in Objective 1. For each sample, we further defined *U*_*fki*_ for subpopulations *i* ∈ {1 : FSW, 2 : clients, * : overall} as the proportions *not* virally suppressed among those living with HIV by 2020, reflecting a summary measure of ART cascade gaps. Using *U*_*fki*_, we defined the main regression predictors as: *D*_*fk*_ = *U*_*fk*\*_ – *U*_*f* 0*_ > 0, reflecting differences in *population overall* viral non-suppression in sample *k* ∈ [1, 10] *vs* the base case (denoted *k* = 0); and *d*_*fki*_ = *U*_*fki*_ – *U*_*fk*\*_ ≶ 0, reflecting differences in *subpopulation-i-specific* viral non-suppression in sample *k vs* the population overall in sample *k* — *i*.*e*., viral non-suppression inequalities.

Next, we defined the following measures of epidemic conditions (*C*_*fj*_) related to sex work, as hypothesized modifiers of the effect of unequal non-suppression on relative CAI and RAI: FSW and client population sizes (% of population overall); average rate of turnover among FSW and clients (per year, reciprocal of duration selling / buying sex); and HIV incidence ratios in the year 2020 among FSW *vs* other women, and among clients *vs* other men. To calculate the HIV incidence ratios, we combined FSW at higher and lower risk, and likewise for clients at higher and lower risk. We used HIV incidence ratios in 2020 to reflect summary measures of risk heterogeneity, rather than including all risk factors from the transmission model, which could lead to overfitting and improper inference due to effect mediation.

Finally, we defined a general linear model for each outcome (CAI, AIR) as:

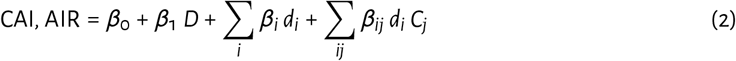

such that each outcome was modelled as a linear sum of the effects of: an intercept term; differential population-level non-suppression in the counterfactual *vs* the base scenario (*D*); differential non-suppression among FSW and clients *vs* the population overall within the counterfactual scenario (*d*_*i*_); and effect modification of *d*_*i*_ by epidemic conditions (*C*_*j*_). We fitted this model for each outcome using generalized estimating equations [30] to control for repeated use of each model fit *f*. We standardized all model variables (*D, d*_*i*_, *C*_*j*_) via 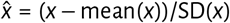 to avoid issues of different variable scales and collinearity in interaction terms. Effect sizes can thus be interpreted as the expected change in outcome per standard deviation change in the variable.^2^

## 3 Results

We first summarize modelled patterns of HIV transmission in the base case (1000 model fits), calibrated to reflect the Eswatini epidemic up to 2020. Our model suggests that transmission within repeat sex work partnerships was a dominant driver of early epidemic growth (Figures B.14 and B.15). However, from approximately 1994 onward, the majority of new yearly infections were transmitted within casual partnerships, including 50% (median) of infections in 2020 in the base case.^3^ By 2020, the majority of cumulative infections acquired were among women with lowest sexual activity (Figure B.12b), while the majority of cumulative infections transmitted were from clients (Figure B.12a). Overall HIV prevalence in 2020 was median (95% CI): 23.8 (22.4, 24.7) % (Figure B.4a), and overall incidence was 6.6 (5.3, 7.6) per 1000 person-years (Figure B.4c). The prevalence ratio between FSW and women overall was 1.78 (1.70, 1.87), and between clients and men overall it was 1.92 (1.49, 2.49) (Figure B.4b). Due to turnover and higher HIV incidence among FSW, achieving similar rates of diagnosis among FSW *vs* other women (Figure B.6a) required approximately twice the rate of testing. Sex work contributed a growing proportion of infections over 2020–2040: from 32% to 42% (Figure B.14).

### 3.1 Objective 1: Influence of cascade differences between subpopulations

Figure B.17 illustrates ART cascade attainment over time in each of the four counterfactual scenarios (80-80-90 overall by 2020), and the base case (95-95-95 overall by 2020). Figure B.16 illustrates overall HIV incidence in each scenario. Figure 2 then illustrates cumulative additional infections (CAI) and additional incidence rate (AIR) in each counterfactual scenario *vs* the base case. Leaving behind neither FSW and nor clients led to the fewest CAI: median (95% CI) 14.9 (10.4, 18.4) % more than the base case by 2030. By contrast, leaving behind both FSW and clients led to the most CAI: 26.3 (19.7, 33.0) % more than the base case by 2030 — a 73 (40, 149) % increase. Leaving behind either FSW or clients resulted in 19.7 (15.4, 23.6) % or 22.7 (17.6, 26.7) % cumulative additional infections *vs* the base case, respectively. Results were similar for additional incidence rate (AIR). In all counterfactual scenarios, the majority of additional infections were transmitted via casual partnerships (Figure B.18a), and acquired among non-FSW women (Figure B.18c).

**Figure 2:**
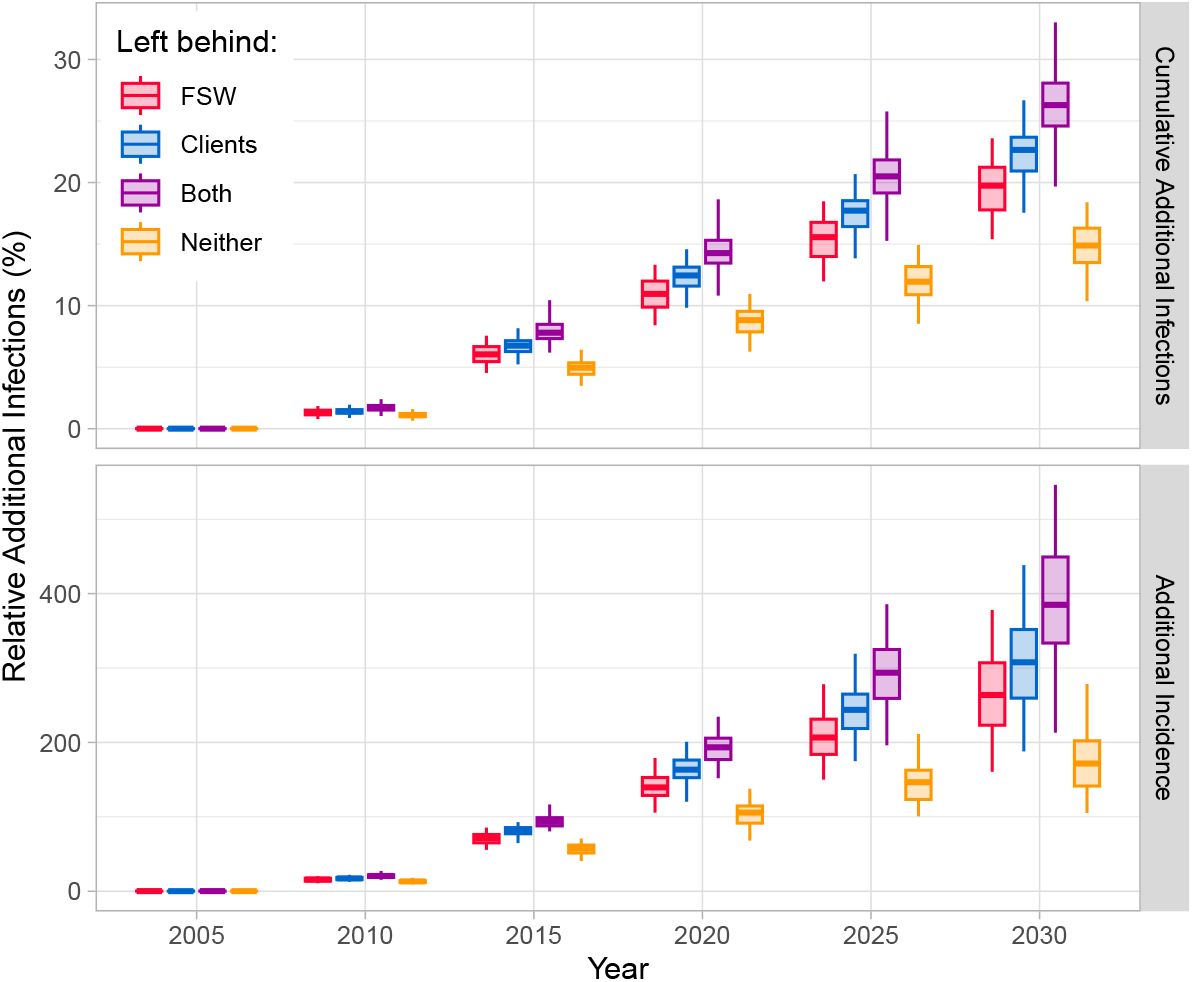
Relative additional infections under counterfactual scenarios *vs* the base case Base case: 95-95-95 by 2020; counterfactual scenarios: 80-80-90 overall by 2020, with reduced cascade (60-40-80: left behind) among FSW, clients of FSW, both, or neither; whiskers, boxes, and midlines: 95% CI, 50% CI, median of model fits.

Patterns of onward transmission were also similar across scenarios (Figure B.18b), though subpopulation contributions increased if they were left behind.

### 3.2 Objective 2: Conditions that maximize the influence of cascade differences

The fitted regression models Eq. (2) indicated that population-overall viral non-suppression (*D*) and relative non-suppression among FSW and clients (*d*_*i*_) each had strong and positive effects on 2030 CAI and AIR outcomes (*p* < 10^−5^). These effects corroborate the results of Objective 1. Regression residuals are shown in Figure B.21, while Figure 3 plots the estimated effects of subpopulation-specific non-suppression *d*_*i*_, plus effect modification by epidemic conditions *C*_*j*_. The effect of relative non-suppression on CAI and AIR among both FSW and clients generally increased with: larger FSW and client population sizes, and larger HIV incidence ratio among clients vs other men. Effects of relative non-suppression among clients further increased with duration buying sex (slower turnover), while effects of relative non-suppression among both groups on CAI specifically further increased with HIV incidence ratio among FSW vs other women.

**Figure 3:**
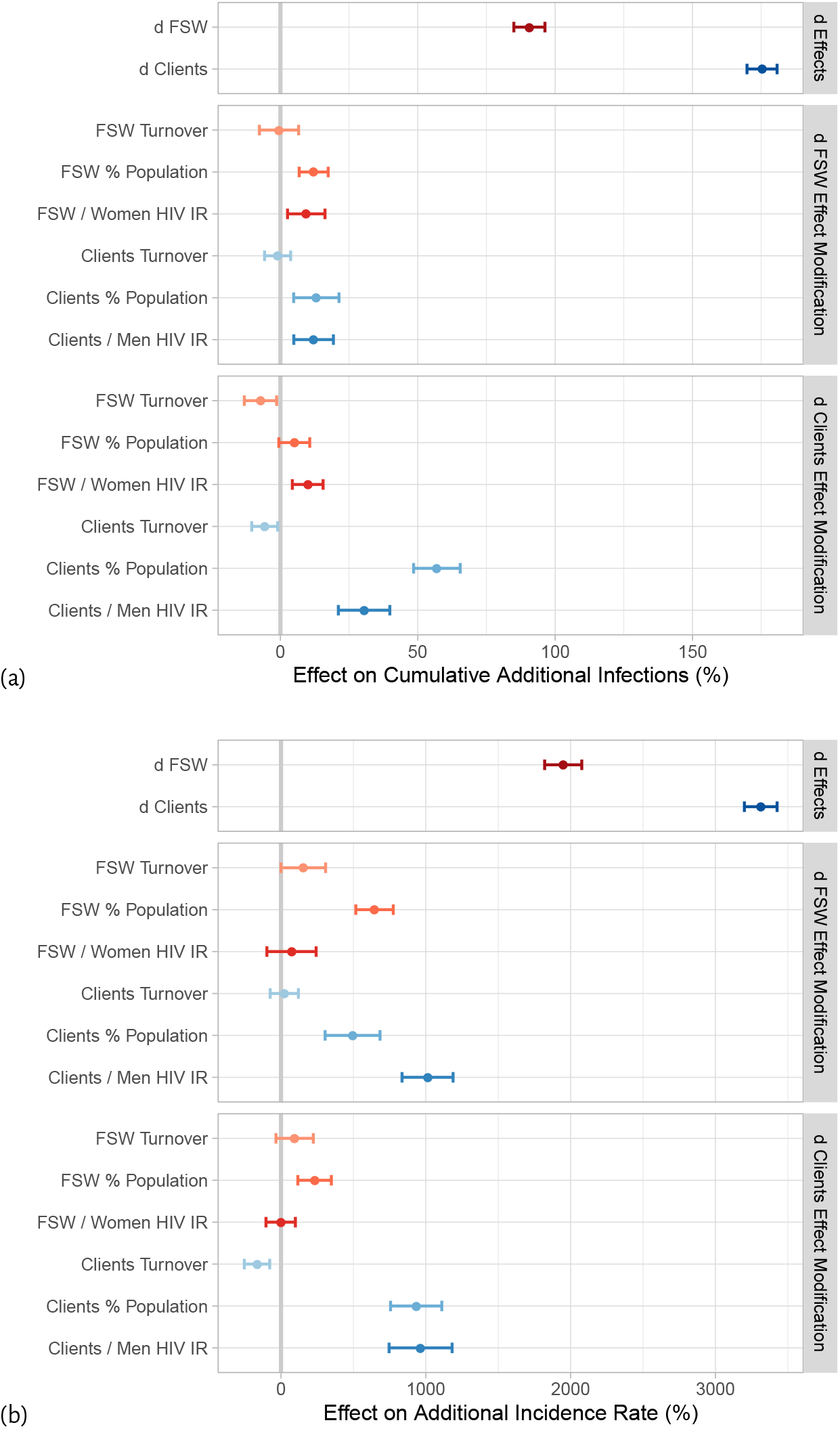
Estimated effects on relative additional infections of disproportionate viral non-suppression (*d*) among FSW and clients *vs* population overall, plus effect modification by epidemic conditions (a) cumulative additional infections, (b) additional incidence rate by 2030 *vs* base case; FSW: female sex workers; Clients: of FSW; IR: incidence ratio in 2020; *di*: difference in subpopulation-*i*-specific viral non-suppression *vs* population overall within counterfactual scenario; points and error bars: mean and 95% CI for each effect estimated via Eq. (2).

## 4 Discussion

We sought to explore how inequalities in the ART cascade that intersect with HIV risk heterogeneity may influence the model-estimated prevention impacts of ART. In our applied analysis of Eswatini, we found that slower cascade scale-up that left behind female sex workers (FSW) and their clients could have resulted in 40–149% more infections by 2030 *vs* slower scale-up alone. We also found that the impact of leaving behind FSW and/or clients was largely determined by their population sizes and HIV incidence relative to the wider population.

Eswatini has recently surpassed 95-95-95 for the population overall [15]. These targets were achieved through numerous initiatives coordinated across sectors, including those led by the *MaxART* program [31,32]. Multiple stakeholders, including people living with HIV, healthcare providers, traditional and religious leaders, community groups, and researchers were engaged via multiple channels, such as Technical Working Groups, Community Advisory Boards, and specific meetings for prioritized groups (men and adolescents) [31,32]. Drawing on this engagement and social science research to understand barriers to care, cascade services were comprehensively strengthened via investments in training, infrastructure, anti-stigma communication, demand creation, and monitoring [31,32].

Among FSW living with HIV in Eswatini, data suggest that 88% were diagnosed and 86% were on ART in 2021 (*i*.*e*., ART coverage was 98% among those diagnosed) [27]. Data on viral suppression among FSW were lacking, and so assumed to be similar to other women in the simulated epidemic (see § A.3.8 for more details). Although lower than 95-95-95, this cascade among FSW living with HIV is higher than in many other regions [7,10]. Strong programs are required to attain high cascades among FSW, considering that women enter and exit sex work (turnover) and likely experience highest risk of HIV acquisition during sex work. That is, programs must ensure higher rates of HIV testing, ART initiation, and retention among FSW *vs* other women to achieve similar cascades. For example, we inferred that rates of HIV testing in 2016 was 80–227% higher among FSW *vs* other women to reproduce observed cascade data during model calibration.

In Eswatini, programs for key populations include safe access to tailored services via drop-in centers (locally known as TRUE), mobile outreach, venue-based, and one-on-one options [27]. Health and clinical services are also integrated with efforts to reduce structural vulnerabilities, including experiences of harassment, violence, and fear of seeking healthcare, through community empowerment, psycho-social and legal supports, and sensitization and training for police and healthcare workers [27]. These programs have been designed and refined with ongoing community leadership and engagement, allowing them to better meet the specific needs of key populations, for whom barriers to engagement in HIV care often intersect with drivers of HIV risk, including economic insecurity, mobility, stigma, discrimination, and criminalization [4,8–11,33]. Our data-informed modeling of cascade scale-up in Eswatini confirms that such an equity-focused approach to ART cascade scale-up can maximize prevention impacts, and accelerate overall reductions in HIV incidence.

Our study highlights the importance of reaching both FSW and their clients, echoing recent modelling studies of South Africa and Cameroon [34,35]. These studies found that gaps in HIV prevention and treatment for clients were among the largest contributors to onward transmission in recent years. Such findings reiterate the need for improved data on both FSW and clients, including estimates of population size, sexual behaviour, and ART cascade attainment. These estimates may be difficult to obtain because individuals are unlikely to report buying or selling sex in population-level surveys due to stigma and criminalization [36] and because many clients are highly mobile (including transient seasonal/occupational migration) [33]. Thus, innovative study designs, bias adjustments, and services may be needed to understand and meet clients’ needs.

While numerous modelling studies have examined the potential prevention impacts of ART cascade scale-up [13] (Appendix B.3), our study is the first to explore the impact of inequalities in ART cascade across subpopulations with consistent population overall cascade across scenarios. Similar work by Marukutira et al. [37] illustrated the limited impact of achieving 95-95-95 for only citizens and not immigrants in Botswana, while Maheu-Giroux et al. [38] illustrated the high cost-effectiveness of prioritizing key populations (including clients) for ART in Côte d’Ivoire. Indeed, our findings are likely generalizable to other epidemic contexts. HIV prevalence ratios between key populations and the population overall are relatively low in Eswatini *vs* elsewhere [39,40]; thus, the impact of cascade inequalities among key populations in other contexts would likely be even greater than we found for Eswatini. Moreover, as HIV incidence declines in many settings, transmissions may become concentrated among key populations [41,42], further magnifying the impact of cascade inequalities.

A primary strength of our analysis is the use of observed ART cascade scale-up to 95-95-95 in Eswatini as the base case, with plausible cascade inequalities explored in counterfactual scenarios. As noted above, the available data suggest that Eswatini has minimized cascade inequalities which persist elsewhere [7]. Thus, our counterfactual scenarios directly estimate the consequences of failing to address these inequalities. Second, drawing on our conceptual framework for risk heterogeneity [13, Table 1] and multiple sources of context-specific data [20–24,27], we captured several dimensions of risk heterogeneity, including: heterosexual anal sex, four types of sexual partnerships, sub-stratification of FSW and clients into higher and lower risk strata, and subpopulation turnover (Table 1, Appendix A). Accurate modelling of risk heterogeneity has been shown to mediate model-estimated ART prevention impacts [43], and is especially important when considering differential ART scale-up across subpopulations. Finally, our analytic approach to Objective 2, in which epidemic conditions are conceptualized as potential effect modifiers represents a unique methodological contribution to the HIV modelling literature.

Our study also has limitations. First, we did not model pre-exposure prophylaxis (PrEP). However, our analyses focus on the time period prior to widespread PrEP availability in Eswatini. Second, we did not consider transmitted drug resistance (TDR). However, drug resistance is more likely to emerge in the context of barriers to viral suppression [44]; thus, lower cascade among those at higher risk would likely accelerate emergence of transmitted drug resistance, and thereby magnify our findings. Finally, our model structure did not include age, and we only considered heterosexual HIV transmission in Eswatini. Future work can explore adaptation of the model to consider PrEP, TDR, age stratification, and additional modes of HIV transmission. While the magnitude of our results may change with such adaptations, we do not expect that the qualitative interpretation would change. In fact, our findings would likely generalize to other transmission networks and determinants of risk heterogeneity, including other key populations and subpopulations such as highly mobile populations and young women [33,45].

In conclusion, the HIV response must remain rooted in context-specific understandings of inequalities in HIV risk and in access to HIV services, which often stem from common upstream factors. Thus, differences in ART cascade within and between subpopulations at higher risk of HIV must be monitored, characterized, and addressed to fully realize the anticipated benefits of ART at both the individual and population levels.

## Data Availability

All code and selected results are available on GitHub.

https://github.com/mishra-lab/hiv-model-eswatini

## Acknowledgements

We thank: Kristy Yiu, Samantha Lo (Unity Health Toronto) for research coordination support; Amrita Rao, Carly Comins (Johns Hopkins University), Alex Whitlock, Korryn Bodner (Unity Health Toronto), and Leigh Johnson (University of Cape Town) for helpful discussions and feedback on model design.

## Footnotes

1 Data on viral suppression for FSW were not available, which we denote as “xx”; no cascade data were available for clients specifically.

2 However, regression coefficient magnitudes should not be compared to indicate variable “importance”, because the standardization applied to each variable is driven by the (arbitrary) variance before standardization. For further discussion on interpretation of standardized regression coefficients, see: stats.stackexchange.com/questions/29781 and links therein.

3 In our model, casual partnerships can be formed by any subpopulation, and these partnerships subsume transactional partnerships.

