## Appendix for "Quantifying the impact of cascade inequalities: a modelling study on the prevention impacts of antiretroviral therapy scale-up in Eswatini"

**Date** 2024 May 10

A Model Details 4

A.1 Model Structure & Notation 4

A.2 Force of Infection 5
A.2.1 Conceptual Development 7
A.2.2 Equations 7
A.2.3 Transmission via Multiple Partnerships 9
A.2.4 Transmission-Driven Seroconcordance 10

A.3 Model Parameterization 10
A.3.1 Preliminaries 10
A.3.2 Data Sources for Eswatini 11
A.3.3 Initialization 13
A.3.4 Probability of HIV Transmission 13
A.3.4.1 HIV Infection Stage 14
A.3.4.2 Sex Act Types 14
A.3.4.3 Circumcision 15
A.3.4.4 Condoms 15
A.3.4.5 Genital Ulcer Disease 15
A.3.5 Prevalence of Transmission Modifiers 16
A.3.5.1 Circumcision 16
A.3.5.2 Condom Use 16
A.3.5.3 Genital Ulcer Disease 19
A.3.6 HIV Progression & Mortality 19
A.3.6.1 HIV Progression 19
A.3.6.2 HIV Mortality 20
A.3.7 Antiretroviral Therapy 20
A.3.7.1 Probability of HIV Transmission on ART 20
A.3.7.2 HIV Progression & Mortality on ART 20
A.3.8 Rates of HIV Diagnosis, ART Initiation, Viral Un-suppression & Re-suppression 21
A.3.8.1 HIV Diagnosis 21
A.3.8.2 ART Initiation 23
A.3.8.3 ART Failure 24
A.3.8.4 Viral Re-suppression 25
A.3.9 Risk Differences Within Sex Work 26
A.3.10 Wider Population: Bias Adjustment 26
A.3.10.1 Reported Partner Numbers 28
A.3.10.2 Bias Adjustment: Rationale 28
A.3.10.3 Bias Adjustment: Approach 30
A.3.10.4 Bias Adjustment: Results 31
A.3.11 Activity Group Sizes 31
A.3.11.1 Female Sex Workers 33

#### Appendix A

### Model Details

**Implementation** The model was implemented in Python v3.8.10 with Numpy v1.24.4, and solved numerically using 4th order Runge-Kutta [1] with a timestep of 0.05 years. Post-hoc analyses were conducted in R v3.6.3. All code and most results are available on GitHub: `github.com/mishra-lab/hiv-model-eswatini`.[4]

#### A.1 Model Structure & Notation

The model aims to capture heterosexual HIV transmission among the Swati population aged 15–49. The modelled population is stratified along five dimensions (Table A.1 and Figure A.1), including: 2 sexes ($s$), 4 activity groups ($i$), 6 HIV states ($h$), and 5 cascade states ($c$); the fifth dimension tracks seroconcordant HIV+ partnerships and includes strata for each of 4 partnership types ($p$) plus 1 extra stratum — see § A.2 for full details. In total, $2 \cdot 4 \cdot (1 + 5 \cdot 5 \cdot 5) = 1008$ states are modelled, since the cascade and partnership dimensions are only applicable to people living with HIV ($h > 1$). Two types of sex acts ($a$) are also considered.

**Sexual Activity** Sexual activity groups were defined to reflect common stratifications in the available data, and persistent differences in HIV incidence and prevalence [2–5]. The lowest sexual activity group ($i = 1$) comprises individuals who had 0-1 sexual partners in the past 12 months (p12m), but did not engage in sex work. The medium activity group ($i = 2$) similarly comprises individuals who had 2+ sexual partners in p12m but did not engage in formal sex work. The highest two activity groups among women ($i = 3, 4$) comprise lower and higher risk FSW (see § A.3.9 for more details), and the highest two activity groups among men ($i = 3, 4$) likewise comprise lower and higher risk clients of FSW.

**Partnership Types** Four types of sexual partnerships are modelled, capturing differences in partnership durations, and trends in condom use relevant to inferred transmission dynamics. The four partnership types are: long-term/spousal partnerships ($p = 1$, lowest condom use, long duration); short-term partnerships ($p = 2$, medium condom use, medium duration); one-off sex work partnerships ($p = 3$, highest condom use, 1 sex act); and repeat sex work partnerships ($p = 4$, medium condom use, medium duration). Figure A.1a illustrates the modelled activity groups and possible partnership types between them.

**HIV Infection & Treatment** HIV infection is stratified into acute-HIV and stages defined by CD4 count (Figure A.1b) to reflect changes in mortality [6], historical ART eligibility [7–10], and, with CD4 as a proxy for viral load, infectiousness [11]. The modelled ART cascade (Figure A.1c) includes the steps associated with the "90-90-90" targets, plus a generic "virally un-suppressed" state reflecting any combination of treatment failure, discontinuation, or loss to follow-up after achieving viral suppression. Loss to follow-up prior to viral suppression is not explicitly modelled, but subsumed into rates of ART initiation and viral suppression.

[4] In the code: R uses 1-based indexing, which match the notation here directly, while Python uses 0-based indexing, which therefore appear as $i \rightarrow i - 1$ in the code. Also, the model code reorders states in the ART cascade dimension for computational efficiency, with $c$ = 1: Undiagnosed; 2: Diagnosed; 3: Virally Un-suppressed; 4: On ART; 5: Virally Suppressed.

Table A.1: Overview of model dimensions and stratifications

| Dimension | Index | | Strata |
|---|---|---|---|
| Sex | ($s$) | 1 | Heterosexual Women |
| | | 2 | Heterosexual Men |
| Activity group | ($i$) | 1 | Lowest Activity |
| | | 2 | Medium Activity |
| | | 3 | Lower Risk Sex Work |
| | | 4 | Higher Risk Sex Work |
| HIV status | ($h$) | 1 | Susceptible |
| | | 2 | Acute HIV |
| | | 3 | CD4 > 500 |
| | | 4 | 350 < CD4 < 500 |
| | | 5 | 200 < CD4 < 350 |
| | | 6 | CD4 < 200 (AIDS) |
| ART cascade | ($c$) | 1 | Undiagnosed |
| | | 2 | Diagnosed |
| | | 3 | On ART |
| | | 4 | Virally Suppressed |
| | | 5 | Virally Un-suppressed |
| Partnership types | ($p$) | 1 | Main / Spousal |
| | | 2 | Casual |
| | | 3 | One-Off Sex Work |
| | | 4 | Repeat Sex Work |
| Sex act types | ($a$) | 1 | Vaginal |
| | | 2 | Anal |

See footnote 4 regarding indices in the code.

#### A.2 Force of Infection

The force of infection equation defines the rate at which susceptible individuals acquire infection. This equation thus integrates mechanistic assumptions and data regarding sexual partnership dynamics and modifiers of transmission risk.

**Conventional Approach** In compartmental HIV transmission models, sexual partnerships are typically quantified using a "change rate" because each "compartment" represents a homogeneous and memoryless population, and so individual partnerships cannot be modelled [12]. In this approach, the probability of transmission per sex act is adjusted (reduced) to account for additional sex with the same partner after transmission has occurred — which we call "post-transmission sex acts" [13]. This adjustment takes the general form:

$$B = 1 - (1 - \beta)^A \tag{A.1}$$

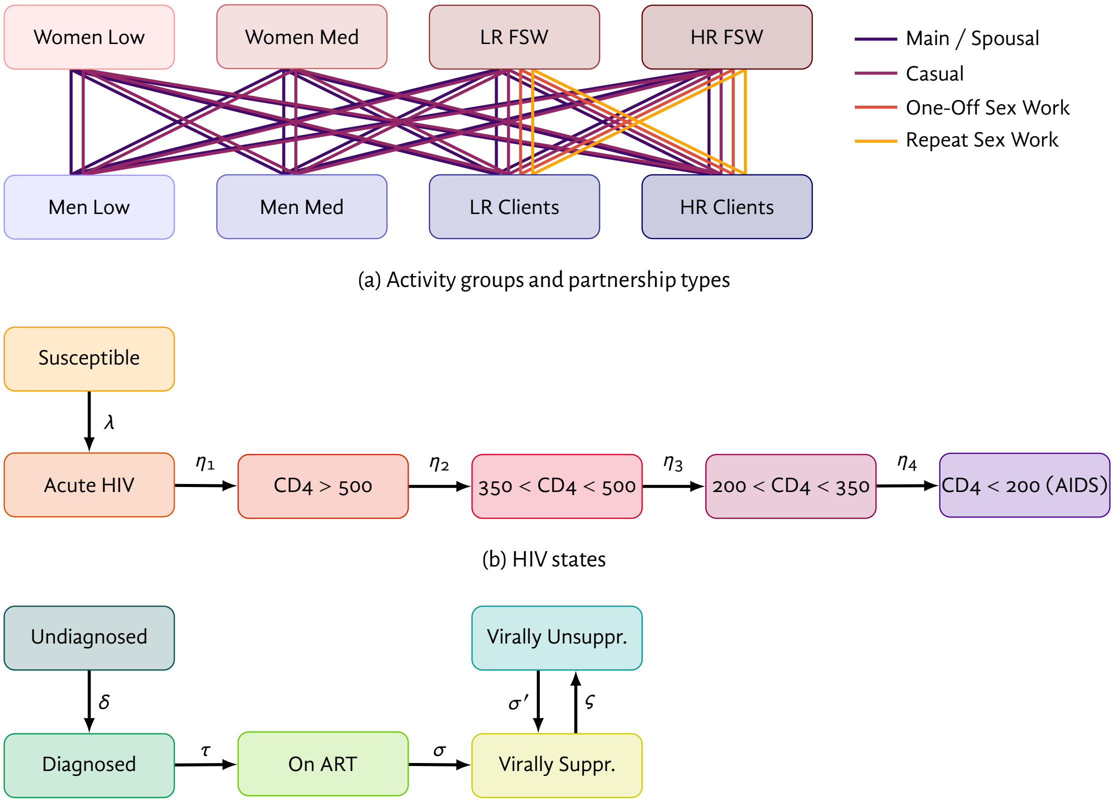

(a) Activity groups and partnership types

(b) HIV states

(c) ART cascade states

Figure A.1: Model structure and transitions

Low: lowest activity; Med: medium activity; LR/HR: lower/higher risk; FSW: female sex workers; Clients: of FSW; CD4: CD4+ T-cell count per $mm^3$; ART: antiretroviral therapy; rates — $\lambda$: force of infection; $\eta$: HIV progression; $\delta$: diagnosis; $\tau$: ART initiation; $\sigma$: viral suppression; $\sigma'$: viral re-suppression; $\varsigma$: ART failure / discontinuation; not shown: turnover amongst activity groups in (a).

where: $\beta$ is the probability of transmission per sex act, $A$ is the number of sex acts per partnership, and $B$ is the total probability of transmission per-partnership. However, this adjustment entails a trade-off [13]:

- If $A$ is chosen to reflect the complete partnership duration, then the probability of transmission is reduced "now" in anticipation of future post-transmission sex acts, thereby underestimating early transmission within partnerships; moreover, the true probability of transmission per sex act $\beta$ likely evolves over long partnerships due to HIV progression and/or prevention interventions, but this evolution is rarely included in Eq. (A.1).
- If $A$ is chosen to reflect an average "partnership-year", then early transmission is not underestimated, but transmission within longer partnerships can be overestimated, because the adjustment in Eq. (A.1) may actually have little effect.

Considering these limitations, we developed a new approach to account for post-transmission sex acts in the force of infection equation, which we call the *Effective Partnerships Adjustment* [13], described below.

##### A.2.1 Conceptual Development

The conventional adjustment Eq. (A.1) estimates the proportion of sex acts which occur after transmission within each partnership. An alternate approach could estimate the proportion of partnerships where transmission has already occurred. In a compartmental model, these partnerships can be tracked as proportions of individuals: namely, all individuals who recently acquired infection *and* all individuals who recently transmitted infection. Here, we use "recent" to mean "before individuals change partners". If some individuals have multiple concurrent partners, then these individuals should not be removed entirely, but their numbers of "effective partnerships" should be reduced by 1. If multiple types of partnerships are considered, then only the partnership type involved in the transmission should be reduced. This adjustment to "effective partnerships" can then be applied until these individuals change partners — at a rate inversely related to the partnership duration: $\delta^{-1}$. However, during this period, these individuals can/should still be modelled to progress as usual through different stages of infection, treatment, etc.

Using this conceptual basis, the *Effective Partnerships Adjustment* introduces a new stratification of the modelled population, denoted $\bar{p}$. The stratum $\bar{p} = 0$ corresponds to no recent transmission, or all "new" (potentially serodiscordant) partnerships. Other strata $\bar{p} \neq 0$ correspond to recent transmission via (to or from) partnership type $\bar{p}$. Figure A.2 illustrates the new stratification together with with the existing HIV infection stratification (Figure A.1b). Following infection, all individuals enter a stratum $\bar{p} \neq 0$ corresponding to the partnership type $p$ by which they were infected. Thus, the rate of entry to this stratum from $S_i$ is defined by the force of infection without aggregating across partnership types: $\lambda_{ip}$. Individuals may then transition from $\bar{p} \neq 0$ to $\bar{p} = 0$ upon forming a new partnership, at a rate $\delta_p^{-1}$. Finally, individuals may re-enter any stratum $\bar{p} \neq 0$ if they transmit infection via partnership type $p$. We denote the corresponding rate as $\lambda'_{ip}$, representing the per-person rate of *transmission*, not *acquisition* as in $\lambda_{ip}$. This rate $\lambda'_{ip}$ is not defined or needed in prior models but we develop the necessary equations below in § A.2.2. The issue of transmission via multiple partnerships is discussed in § A.2.3.

##### A.2.2 Equations

Since partnership duration is now considered separately and explicitly, we do not define any per-partnership probability of transmission $B$. Rather, we define the force of infection to directly include the frequency of sex per partnership $F$ and probability of transmission per sex act $\beta$. However, the mixing is slightly more complicated than usual, since the number of "effective partnerships" depends on infection status. In addition, these partnerships are now defined as numbers of concurrent partners $K$, rather than rates of partnership formation $Q$.

Let $M_{pii'}$ be the total (population-level, not per-person) number of type-$p$ partnerships between group $i$ and group $i'$ (here we omit sex notation $s, s'$ for simplicity). This "mixing matrix" $M_{pii'}$ can be defined in several

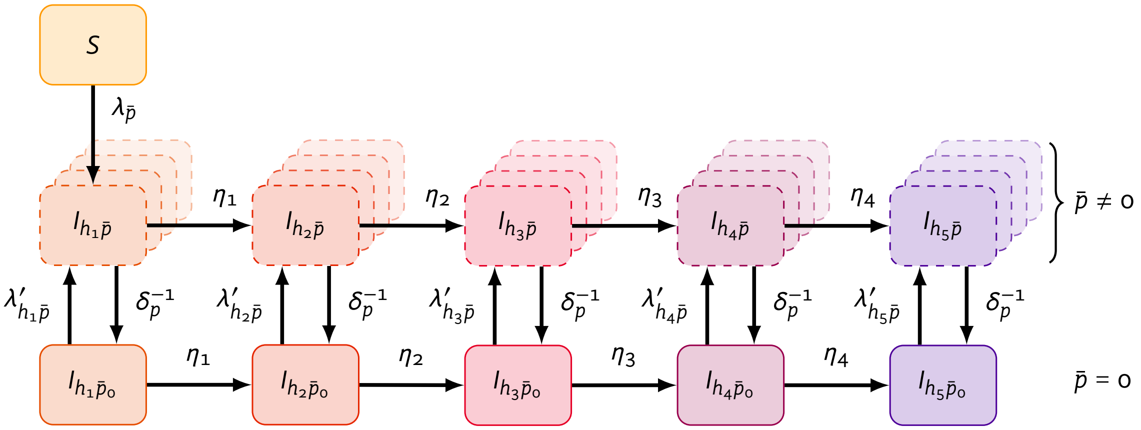

Figure A.2: Modelled states and transitions related to HIV infection, and a new stratification $\bar{p}$ to track the proportions of individuals in partnerships where transmission already occurred

$S$: susceptible; $I_h$: infectious in stage $h$; $p$: partnership type; $\bar{p}$: new stratification, where $\bar{p} = 0$ reflects no recent transmission (all new partnerships), and $\bar{p} \neq 0$ reflects recent transmission via a type-$p$ partnership; $\lambda$: force of infection per susceptible; $\lambda'$: force of infection per infectious; $\eta$: rate of progression between infection stages; $\delta$: partnership duration.

ways (*e.g.*, § A.3.16), based on the total numbers of "effective partnerships" among each group: $M_{pi}$, $M_{pi'}$, plus some parameter(s) specifying mixing patterns. Working backwards, we start by defining $M_{pi}$ (and likewise $M_{pi'}$) via the sum across health statuses — *i.e.*, susceptible, and different stages of infection $h$:

$$M_{pi} = M_{S,pi} + \sum_h M_{I,pih} \tag{A.2}$$

We then define the total numbers of partnerships among susceptible individuals as:

$$M_{S,pi} = S_i K_{pi} \tag{A.3}$$

and likewise for individuals in infection stage $h$:

$$M_{I,pih} = I_{ih,\bar{p}=p}(K_{pi} - 1) + \sum\nolimits_{\bar{p}\neq p} I_{ih\bar{p}}\, K_{pi} \tag{A.4}$$

This Eq. (A.4) is the key equation whereby the numbers of "effective type-$p$ partnerships" among individuals in stratum $\bar{p}$ are reduced by 1. This reduction is then propagated through the mixing patterns when defining $M_{pii'}$. Next, we define the total (population-level, not per-person) rate of transmission from group $i'$ and infection stage $h'$ to group $i$ via type-$p$ partnerships as:

$$\Lambda_{pii'h'} = F_p \beta_{pii'h'} M_{pii'} \left(\frac{M_{S,pi}}{M_{pi}}\right) \left(\frac{M_{I,pi'h'}}{M_{pi'}}\right) \tag{A.5}$$

where the two fractions represent the proportions of all partnerships $M_{pii'}$ formed between susceptible individuals from group $i$ ($M_{S,pi}$) and infectious individuals in group/infection stage $i'h'$ ($M_{I,pi'h'}$). The per-

person transmission rates to group $i$, and from group $i'h'$ can then be defined as:

$$\lambda_{pi} = \sum_{i'h'} \Lambda_{pii'h'} \, S_i^{-1} \tag{A.6}$$

$$\lambda'_{pi'h'} = \sum_{i} \Lambda_{pii'h'} \, I_{i'h'}^{-1} \tag{A.7}$$

For the purposes of solving the model, we can skip division by $S_i$ and $I_{i'h'}$ in Eqs. (A.6) and (A.7), since $\lambda'_{pi}$ and $\lambda'_{pi'h'}$ are immediately multiplied by $S_i$ and $I_{i'h'}$, respectively, in the system of differential equations — *i.e.*, we need absolute, not per-person, rates of transmission.

Finally, and to reiterate from above, infected individuals in stratum $I_{ih\bar{p}}$ are assumed to form new partnerships at a rate $\delta_p^{-1}$, and thereby transition to stratum $I_{ih\bar{p}_0}$ ("all new partners"); and otherwise transition between infection stages, cascade of care, activity groups, etc. as usual, as illustrated in Figure A.2.

##### A.2.3 Transmission via Multiple Partnerships

We do not explicitly model the proportions of infected individuals who recently acquired and/or transmitted infection via two different partnership types, or two partnerships of the same type. To do so, the required size of the new dimension $\bar{p}$ would be at least $2^P$, not $P + 1$, where $P$ is the number of different partnership types modelled. For transmission via three different partnerships, the required size would be at least $3^P$, and so on. Indeed, this exponential relationship is related to the challenge of specifying all possible combinations of partnership states in pair-based models [14]. However, under frequentist assumptions, we can equivalently model two transmissions by one individual as one transmission each by two individuals. Thus, we can transfer two individuals from $I_{ih\bar{p}_0}$ to $I_{ih\bar{p}_1}$ and $I_{ih\bar{p}_2}$ (one each) under the $P$+1 stratification, instead of just one individual from $I_{ih\bar{p}_0}$ to "$I_{ih\bar{p}_{12}}$" under one of the exponential ($x^P$) stratifications.

In fact, $I_{ih\bar{p}_0}$ can be *negative* (but only for $\bar{p} = 0$), because the dimension $\bar{p}$ is only relevant to Eq. (A.4); in all other contexts and equations, we use $I_{ih} = \sum_{\bar{p}} I_{ih\bar{p}}$, which must be positive as usual. Moreover, we can also have $I_{ih\bar{p}} > I_{ih}$, provided that:

$$I_{ih\bar{p}} \leq I_{ih} K_{pi} \tag{A.8}$$

reflecting the situation where 100% of $I_{ih}$ have recently acquired and/or transmitted infection via at least one type-$p$ partnership, or 50% via at least two partnerships, etc. This situation can therefore only arise in the context of multiple concurrent type-$p$ partnerships: $K_{pi} > 1$. If $I_{ih\bar{p}} > I_{ih}$, then $I_{ih\bar{p}_0}$ *must* be negative, but we can show that Eq. (A.4) still yields the correct value of $M_{I,pih}$. With this perspective, the constraint in Eq. (A.8) may be more intuitive: we cannot "remove" more than the total number of partnerships. This constraint should also be easy to guarantee for a small enough timestep, because in Eq. (A.4), $M_{I,pih}$ approaches zero as $I_{ih\bar{p}}$ approaches $I_{ih}K_{pi}$ — i.e. all type-$p$ partnerships become HIV+ seroconcordant, and no more transmission can occur via these partnerships until partners change.

##### A.2.4 Transmission-Driven Seroconcordance

Another benefit of the proposed approach is that we can quantify the proportion of partnerships that are seroconcordant due to prior transmission within the partnership. We call these partnerships "transmission-driven seroconcordant" to distinguish them from seroconcordant partnerships formed by chance — or due to serosorting [15,16] — among two previously infected individuals. For this proportion, the numerator is the population size of stratum $\bar{p} = p$, while the denominator is the total number of type-$p$ partnerships among (a) all individuals, or (b) infected individuals only:

$$\text{TDSC}_{p*} = \frac{I_{*,\bar{p}=p}}{K_{p*} \sum_{\bar{p}} X_{*\bar{p}}} \qquad \begin{cases} X = I + S & \text{(a)} \\ X = I & \text{(b)} \end{cases} \tag{A.9}$$

where $*$ could specify any subset of the population defined by modelled strata — *e.g.*, sex, activity group, HIV state, etc. While denominator (a) may be interesting conceptually, denominator (b) can be directly interpreted as a relative reduction in onward transmission risk among infected individuals. The actual reduction would be weighted by the probability of transmission per sex act, sex frequency, etc. Note also that a single post-transmission partnership will be "double-counted" in the numerator of Eq. (A.9), because both infected individuals cannot transmit via this partnership.

#### A.3 Model Parameterization

Model parameterization involves specification of parameter values (model inputs), such as proportions, probabilities, rates, and ratios. This section describes the data, analyses, and assumptions used to derive these inputs. A summary of calibrated parameters, calibration targets, and methodology is given in § A.4.

##### A.3.1 Preliminaries

**Deriving Prior Distributions** Uncertainty distributions for most parameters and calibration targets were estimated by fitting a parametric distribution to specified quantiles. Let $f(x \mid \theta)$ be the probability density function of random variable $x$ (parameter or calibration target) given distribution parameters $\theta$. Then $F(x \mid \theta) = \int_0^x f(\tau)\, d\tau$ is the cumulative distribution function, and $Q(p \mid \theta) = F^{-1}(p \mid \theta)$ is the quantile function. Our objective is to estimate $\theta$, given a set of quantiles (*e.g.*, $q = \{q_{2.5}, q_{97.5}\}$ for the 95% CI). For each estimation, we minimized the following error function, using the L-BFGS-B algorithm [17]:

$$J(\theta) = \sum_i \left| q_i - Q(p_i \mid \theta) \right|^{\omega} \tag{A.10}$$

where $\omega$ can specify absolute differences ($\omega = 1$) or squared differences ($\omega = 2$) to improve convergence. Distribution fit was validated visually using a plot of the distribution quantiles $Q(p_i \mid \theta)$ *vs* the target quantiles $q_i$, overlaid on the density distribution $f(x \mid \theta)$; *e.g.*, Figure A.3.

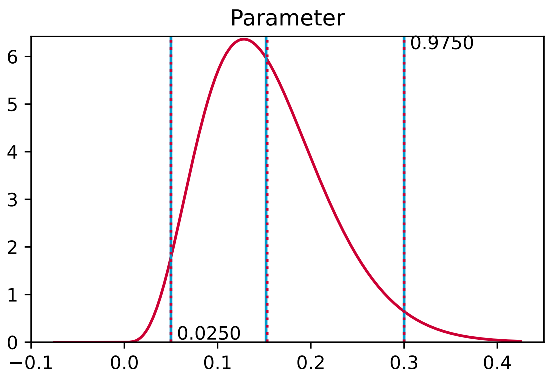

Figure A.3: Example distribution fitting validation plot

BAB distribution fit to $\{q_{2.5} = .05, q_{97.5} = .30\}$; blue solid lines: target quantiles $q_i$; red dotted lines: distribution quantiles $Q(p_i \mid \theta)$; red solid line: density distribution $f(x \mid \theta)$.

**Beta Approximation of the Binomial (BAB) Distribution** Numerous model parameters and calibration targets represent population proportions. Such proportions can be estimated as $\rho = n/N$, where $N$ is the sample size and $n$ is the number of individuals with the characteristic of interest. The uncertainty around $n$ is then given by the binomial distribution:

$$p(n) = \binom{N}{n} \rho^n (1-\rho)^{N-n} \tag{A.11}$$

However, Eq. (A.11) is only defined for discrete values of $n$. It is more convenient to have a continuous distribution for $\rho$, for sampling parameters and evaluating the likelihood of calibration targets, since compartmental models can have non-whole-number population sizes. For this purpose, we use a beta approximation of the binomial distribution (BAB):

$$p(\rho) = \frac{\Gamma(\alpha+\beta)}{\Gamma(\alpha)\,\Gamma(\beta)} \rho^{\alpha-1} (1-\rho)^{\beta-1} \tag{A.12}$$

with $\alpha = N\rho$ and $\beta = N(1-\rho)$. Unlike the approximation by a normal distribution, the beta distribution ensures that $\rho \in [0,1]$. Figure A.4 illustrates the approximation for $N = \{10, 20, 40\}$ and $\rho = \{0.01, 0.1, 0.5\}$.

##### A.3.2 Data Sources for Eswatini

Major HIV data sources for Eswatini are summarized in Table A.2, and briefly described as follows. Summary statistics were extracted from reports and publications in all cases, except two FSW surveys [18,19], for which individual-level data were obtained and analyzed directly in § A.3.9.

**General Population** The 2006–07 Demographic and Health Survey (DHS) [2] was the first nationally representative, household-based survey in Eswatini covering numerous demographic and health topics. The survey included dried blood spot HIV testing, covering 88.1% of women and 81.1% of men. Adjusted HIV prevalence was stratified by sex, age, and other demographic factors, as well as marital status and numbers of sexual partners in the past 12 months (p12m). The survey also included data on sexual health and behaviour,

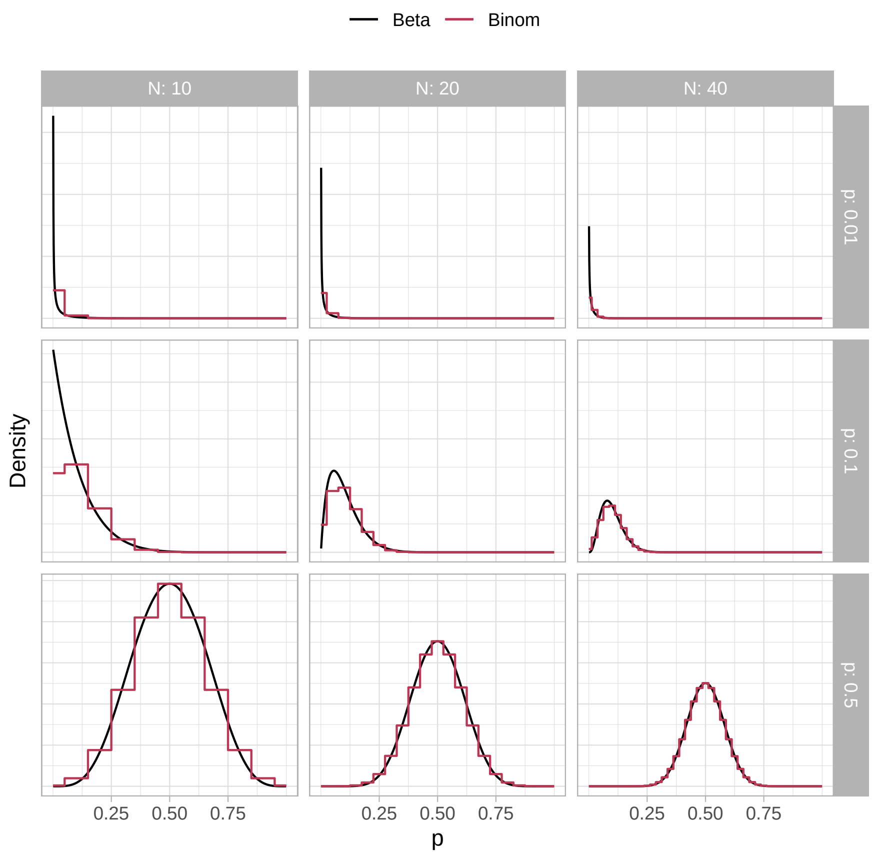

Figure A.4: Beta approximation of the binomial distribution (BAB)

Table A.2: Main data sources for Eswatini

| Ref | ID | Dates [a] | Population [b] | N [c] | HIV [d] |
|---|---|---|---|---|---|
| [2] | DHS'06 | 07/06 – 02/07 | GP 15+ | 9,143 | P |
| [20] | SHIMS1 | 12/10 – 06/11 | GP 18–49 | 18,169 | P, I |
| [5] | SHIMS2 | 08/16 – 03/17 | GP 15+ | 9,146 | P, I |
| [21] [e] | SHIMS3 | 05/21 – 11/21 | GP 15+ | 12,043 | P, I |
| [18] | KP'11 | 09/11 – 10/11 | KP 15+ | 328 | P |
| [19] | KP'14 | 09/14 – 01/15 | KP 18+ | 781 | — |
| [22] | KP'21 | 10/20 – 01/21 | KP 18+ | 676 | P, I |

[a] Baseline data collection (MM/YY); [b] GP: general population; KP: key populations (female sex workers, men who have sex with men); [c] Respondents aged xx–49 who completed baseline survey; [d] Estimates of HIV via blood test, P: prevalence, I: incidence; [e] Preliminary findings only.

including condom use at last sex, STI symptoms in p12m, and HIV testing history. The first two SHIMS in 2010–11 [20] and 2016–17 [5] were conducted with the aim of estimating population-level incidence before and after *Soka Uncobe*. Similar to the DHS, these SHIMS were nationally representative, household-based surveys; however, SHIMS focused specifically on HIV variables, and additionally estimated ART cascade steps and HIV incidence. In SHIMS1 [20], a large prospective 6-month cohort was used to estimate incidence and validate recency testing [23] as a cross-sectional measure of incidence, whereas in SHIMS2 [5], incidence was estimated via the validated recency test. Compared to the DHS, participation rates were lower in SHIMS1 (81.7% and 65.0% among women and men, for the baseline survey), and similar in SHIMS2 (88.0% and 78.5%). SHIMS3 (2021) was recently completed, but so far only preliminary findings relevant to calibration targets are available in a summary report [21].

**Female Sex Workers** The first behavioural surveillance survey among FSW in Eswatini reached only 37 FSW during 2001–02 and did not include HIV testing [22]. In 2011, a larger survey reached 328 FSW via respondent-driven sampling and included HIV testing and detailed behavioural data [18,24]. This study found unadjusted HIV prevalence of 70.3%, highlighting a concentrated sub-epidemic among this key population even within the high-prevalence Eswatini epidemic [18]. A follow-up study in 2014 aimed to estimate FSW and MSM population sizes, identify venues for HIV service delivery, and provide additional data on service gaps [19]; this study used location-based snowball sampling [25] to reach 781 FSW, but did not include HIV testing. Finally, a fourth survey in 2020–21 sought to estimate FSW and MSM population sizes, HIV prevalence and incidence, prevalence of viral suppression, as well as identify behavioural and structural factors associated with HIV [22]; the study recruited 676 FSW via respondent-driven sampling.

##### A.3.3 Initialization

The first cases of HIV and AIDS in Eswatini were diagnosed in 1986 and 1987, respectively [26], although HIV may have been present several years earlier [27]. As such, we initialize the model in 1980 with no HIV, and simulate introduction of HIV at a random year between 1980 and 1985 (uniform prior). HIV introduction is modelled as exogenous infection of 0.01% (~ 24) individuals in the model,[5] distributed across activity groups in proportion to their size, comprising: 5% acute HIV ($h = 2$), 65% with CD4 > 500 ($h = 3$) and 30% with 350 < CD4 < 500 ($h = 4$), all undiagnosed ($c = 1$). The population size of EmaSwati aged 15–49 in 1980 was defined as 243,000 from [28].

##### A.3.4 Probability of HIV Transmission

We parameterized the overall probability of transmission per sex act $\beta$ as the product of a base rate $\beta_0$, and independent relative effects corresponding to multiple factors. Such factors (indexed $f$) included: sex act type $a$, condom use, prevalence of circumcision among susceptible men, partner HIV infection stage $h'$ and viral suppression via ART $c'$, as well as prevalence of STI co-infection/symptoms among both partners. Thus, $\beta$ was

[5] No further import/export of HIV to/from Eswatini is considered thereafter in the model. HIV transmission between Eswatini and neighbouring countries, including South Africa and Mozambique, has likely continued throughout the epidemic due to labour migration and other factors [27]. However, we assume that such transmissions have low overall influence on epidemic dynamics.

defined as:

$$\beta_{asis'i'h'c'} = \beta_0 \, R_{\beta,f_1} \ldots R_{\beta,f_N} \tag{A.13}$$

The impact of each factor (except ART) on the probability of HIV transmission is described in the following subsections, while the prevalence of each factor is given in § A.3.5. The impact of ART on transmission is described in § A.3.7.1.

###### A.3.4.1 HIV Infection Stage

Boily et al. [11] synthesized per-act transmission probability in the absence of ART from 43 studies in 25 populations. Among 7 studies reporting stage of HIV infection (early, asymptomatic, late), infection stage explained 95% of variance in per-act probability of transmission in [11]. Such differences in transmission are most likely due to differences in viral load, which is associated with HIV stage [29,30]. The probability of transmission during the middle asymptomatic period, was reported as mean (95% CI) 0.072 (0.053, 0.097)% per act, reflecting $\beta_0$. To improve model fit (see § A.4), the 95% CI was increased to (0.053, 0.15)%, which was used to define a gamma prior distribution for $\beta_0$. This probability was assumed to apply to vaginal intercourse, based on the studies considered.

For early infection ($h = 2$), Boily et al. [11] estimated the relative infectiousness of the first 5 months of infection as 9.2 (4.5, 18.8) times higher than the asymptomatic period. However, both the duration and infectiousness of the acute phase have been long debated [31–33]. In a recent reanalysis of the Rakai cohort data, Bellan et al. [34] estimate a much smaller contribution of the acute phase to overall infection, summarized as 8.4 (0, 63) "excess hazard-months". This excess risk represents the joint uncertainty and collinearity in the estimated duration of 1.7 (.55, 6.8) months and relative infectiousness of 5.3 (.79, 57). Thus, we sampled the duration $\delta_{h=2}$ from a gamma prior with mean (95% CI) 1.7 (.55, 6) months, and relative infectiousness $R_{\beta,h'=2}$ from a gamma prior with 5.3 (1, 15) times the asymptomatic period (confidence intervals were adjusted to fit the gamma distributions, and to ensure 1 < excess hazard-months < 63).

For late-stage disease, defined as 6-15 months before death in [11], Boily et al. estimated the relative rate of transmission as 7.3 (4.5, 11.9). However, we defined later HIV stages by CD4 count, including 200 < CD4 < 350 ($h = 5$) and CD4 < 200 ($h = 6$, AIDS), which reflects closer to 50 and 18 months before death in the absence of ART, respectively. Therefore, we combined estimates from several sources [11,30,35] to define two gamma prior distributions with mean (95 CI%) 1.6 (1.3, 1.9) and 8.3 (4.5, 13), for the relative rate of HIV transmission in these two stages ($h = 5, 6$), respectively. For CD4 > 350 ($h = 3, 4$), we assumed no change from the baseline probability $\beta_0$.

###### A.3.4.2 Sex Act Types

The model considers vaginal and anal intercourse, further stratified by sex (male-to-female/insertive *vs* female-to-male/receptive). For vaginal intercourse, evidence for differential risk by sex is mixed, with some studies reporting no difference [35,36], and others reporting up to 2-times higher male-to-female ($s' = 2, s = 1$) transmission *vs* female-to-male ($s' = 1, s = 2$) [11,37]. To reflect this uncertainty, we sampled the relative rate of male-to-female *vs* female-to-male transmission from Unif [1, 2]; in applying this relative rate, both male-to-female and female-to-male transmission probabilities were adjusted such that the overall mean was preserved.

Baggaley et al. [38] synthesized the per-act transmission probability for anal intercourse, with most data from MSM studies. Analyses in [38] were not stratified by HIV stage, so we assumed the same relative rates derived in § A.3.9 applied equally to vaginal and anal intercourse. Overall female-to-male (insertive) per-act transmission probabilities were similar for anal intercourse [39] (without ART): 0.14 (0.04, 0.29)% *vs* vaginal intercourse [11] (without commercial sex exposure): 0.164 (0.056, 0.481)%; thus we assumed that female-to-male (insertive) transmission probabilities for anal *vs* vaginal intercourse were equal. By contrast, male-to-female (receptive) per-act transmission probabilities were approximately 10 higher in anal intercourse [38] (without ART): 1.67 (0.44, 3.67)% *vs* vaginal intercourse [11] (without commercial sex exposure): 0.143 (0.088, 0.233)%; thus we assumed a fixed 10-fold increase in male-to-female transmission probability for anal *vs* vaginal intercourse. See § A.3.14 for sex act frequency within each partnership type.

###### A.3.4.3 Circumcision

Relative risk in per-act HIV female-to-male transmission for circumcised *vs* uncircumcised men via vaginal intercourse has been estimated as approximately 0.50, with 95% CI spanning (0.29, 0.96) [11,36,40]. Since circumcision status is unrelated to the research question, we fixed this effect at 50% relative risk. For anal intercourse, Wiysonge et al. [41] estimated that circumcision resulted in .27 (.17, .44) the odds of HIV acquisition for the insertive partner. It can be shown that relative reduction in incidence represents a lower bound on relative reduction in per-act transmission probability.[6] Thus, for anal intercourse, we similarly fixed the per-act effect at 27%. Finally, there is inconclusive evidence to suggest that circumcision status affects male-to-female/receptive transmission [41,42], so we assumed no effect. See § A.3.5.1 for prevalence of circumcision in Eswatini over time.

###### A.3.4.4 Condoms

The most recent meta-analysis of condom effectiveness (when used) in heterosexual couples by Giannou et al. [43] estimated a relative risk of approximately 0.26 (0.13, 0.43). No significant differences were noted between female-to-male *vs* male-to-female transmission. A recent study among men who have sex with men found a similar effect for anal sex [44]. Thus, condom effectiveness was fixed at 74%. See § A.3.5.2 for the proportions of sex acts where condoms are used in Eswatini over time (parameterized separately).

###### A.3.4.5 Genital Ulcer Disease

Genital ulcer disease (GUD) is another another established risk factor for HIV transmission [45,46]. Some, but not all GUD is associated with sexually transmitted infections (STIs), and some, but not all STIs can

---

[6] See § A.3.9 for more discussion.

cause GUD [46]. GUD is thought to increase both HIV susceptibility and infectiousness through a variety of mechanisms [46–48], but HIV may also facilitate transmission of various STIs through immunosuppression [49]. The meta-analysis by Boily et al. [11] found that presence of STI alone was not associated with increased HIV transmission: RR 1.11 (0.30, 4.14), but GUD was: RR 5.29 (1.43, 19.6), with most studies examining GUD among the HIV-susceptible partner. One study [50] estimated RR 2.58 (1.03, 5.69) of transmission for GUD among the HIV-positive partner. Most studies defined GUD status as any experience of symptoms during the study period (*e.g.*, past 12 months, p12m), since precise delineation of GUD episodes is challenging. Morover, individuals may take action to reduce onward STI transmission, such as accessing treatment, having less sex, and using condoms [2]. Thus, the true effect of GUD on HIV transmission via unprotected sex during active GUD episodes may be larger [51]. However, if estimates of GUD prevalence and GUD effect (on HIV transmission) use consistent definitions (*e.g.*, any GUD in p12m), then the time-averaged effect can be applied without need to estimate GUD episode duration. On the other hand, association of GUD and HIV transmission may not reflect causation, but rather confounding by uncontrolled exposure risk. As such, we applied factors for increased susceptibility and infectiousness due to GUD in accordance with group-specific p12m GUD prevalence (see § A.3.5.3), with median 95% CI (1.2, 7.0) and (1.2, 3.4) (gamma priors), respectively.

##### A.3.5 Prevalence of Transmission Modifiers

###### A.3.5.1 Circumcision

Traditional (non-medical) circumcision in Eswatini is rare, reported as approximately 0.7% of men aged 15-49 in 2016 [5]. Voluntary medical male circumcision (VMMC) increased circumcision coverage to 8.2% by 2007, following demand for mainly hygienic reasons [2]. In 2007, the government further increased scale-up of VMMC services as part of HIV prevention efforts [2], leading to 17.1% coverage in 2011 [20], 30.0% in 2017 [5], and 37% in 2021 [52]. Since VMMC continues to be a key element of Eswatini's HIV response [52], we assumed that coverage could reach and plateau at 50–90% (95% CI) by 2050. There is minimal evidence of differential condom use by circumcision status [20], so we assumed no differences. Similarly, while circumcision differed by union status in [5] (*e.g.*, 22.1% circumcised among men in a union *vs* 31.7% among men not in a union), differences did not persist after re-stratifying these men into groups with 0-1 *vs* 2+ partners per year, as described in § A.3.10. In Zambia, circumcision status was not associated with paying for sex [53].

###### A.3.5.2 Condom Use

Condom use is typically reported as either categorical for a recent period, usually 30 days, *e.g.*, "*never, rarely, sometimes, often, always*"; or binary for the most recent sex act. Both report types may be subject to reporting bias, but the "last sex" more directly translates into a proportion of sex acts. The direction of reporting bias may vary with social context, with [54] suggesting over-reporting of condom use, and [55] suggesting under-reporting of condom use. As such, we made no systemic adjustments to the available condom use data. Table A.3 summarizes the available condom use data for Eswatini, deriving from [2,5,18,19,56,57].

Table A.3: Estimates of condom use in Eswatini

| Partnership Type | Year | Population | Type | % | (95% CI) | Ref | Notes |
|---|---|---|---|---|---|---|---|
| Main | 2006 | Women | last sex | 23.5 | (23.2, 23.9) | [2] | a |
| | | Men | last sex | 23.1 | (19.4, 26.9) | [2] | a |
| | 2016 | Women | last sex | 52.7 | (52.5, 52.9) | [5] | a |
| | | Men | last sex | 33.7 | (30.8, 36.7) | [5] | a |
| Main or Casual | 1988 | Women | currently | 0.6 | (0.4, 1.3) | [56] | b |
| | | Men | currently | 7.3 | (5.9, 12.1) | [56] | b |
| | 2002 | FSW | last sex | 60 | — | [57] | cd |
| | | | always | 45.8 | — | [57] | cd |
| | 2006 | Women | last sex | 36.5 | — | [2] | |
| | | Men | last sex | 47.2 | — | [2] | |
| | 2011 | Women | always | 30 | — | [2] | |
| | | Men | always | 34 | — | [2] | |
| | | FSW | last sex | 51.1 | (41.8, 60.4) | [18] | de |
| | | | always | 20.8 | (14.7, 26.9) | [18] | de |
| | 2014 | FSW | last sex | 80.6 | (64.7, 89.6) | [19] | g |
| | 2016 | Women | last sex | 58.3 | — | [5] | |
| | | Men | last sex | 53.1 | — | [5] | |
| Casual | 2006 | Women | last sex | 53.5 | — | [2] | |
| | | Men | last sex | 66.0 | — | [2] | |
| | 2016 | Women | last sex | 64.9 | — | [5] | |
| | | Men | last sex | 73.7 | — | [5] | |
| Sex Work Unspecified | 2002 | FSW | last sex | 90 | — | [57] | d |
| | | | always | 74.4 | — | [57] | d |
| | 2020 | FSW | always | 50 | — | [22] | |
| New Sex Work | 2011 | FSW | last sex | 84.8 | (57.9, 92.4) | [18] | ef |
| | | | always | 56.7 | (47.8, 65.6) | [18] | d |
| | 2014 | FSW | last sex | 88.5 | (54.9, 95.9) | [19] | g |
| Regular Sex Work | 2011 | FSW | last sex | 82.9 | (56.8, 90.0) | [18] | ef |
| | | | always | 38.6 | (29.5, 47.7) | [18] | e |
| | 2014 | FSW | last sex | 85.6 | (47.9, 95.0) | [19] | g |

[a] Back-calculated as described in § A.3.5.2; [b] 95% CI from urban & rural data; [c] Described as "non-paying partners" in the survey; [d] Two major cities only (Manzini & Mbambane); [e] RDS-adjusted; [f] 95% CI lower bound reduced by 25% due to possible reporting bias; [g] 95% CI bounds from regions with lowest and highest reported condom use.

**Main/Spousal & Casual** No direct estimates of condom use in main/spousal partnerships are available; condom use at last sex (with a non-paying partner) was either reported overall or for casual partners only.[7] However, the proportions of individuals with various relationship statuses (*e.g.*, polygynous union, non-polygynous union, not in a union, see § A.3.10) can be used to back-calculate condom use in main/spousal partnerships for both 2006 [2] and 2016 [5]. To do so, we assumed whether "last sex" among individuals in unions with 2+ partners was with their main/spousal partner or with a casual partner; or more generally, what proportion of most recent sex acts was with a casual partner. We repeated the back-calculation assuming 5% and 95%, yielding the confidence intervals shown in Table A.3. Estimates of condom use in non-paying partners were lower among FSW *vs* the wider population in 2011 (20.8% *vs* ~32% "always"), but higher in 2014-16 (80.1% *vs* ~55.7% "last sex"). Therefore, we assumed no differences in condom use among FSW *vs* the wider population for main/spousal or casual partnerships.

**Sex Work** All data on sex work partnerships in Eswatini is from FSW (*i.e.*, not their clients). A 2001 study in Ghana [58] suggested that FSW were more likely than their clients to report having used a condom. As such, we adjusted the lower bound of 95% CI for condom use in sex work partnerships ($p = 3, 4$) as either 75% of the reported lower bound, or the lowest reported region-specific estimate. Estimates for 2002 [57] were obtained from two major cities only (Manzini and Mbambane); since early condom availability was mainly urban, treated these estimates as 95% CI upper-bounds, and defined the lower bound as 20% of the reported values.

**Anal Sex** Owen et al. [59] estimate that among FSW globally, condom use in anal sex is approximately 79 (66, 94)% that of condom use in vaginal sex.[8] In Eswatini [18,19], relative condom use in anal sex *vs* vaginal sex ranged from 44% among new clients in 2011 to 88% among regular clients in 2014. So, we sampled relative condom use in anal *vs* vaginal sex from a BAB prior distribution with 95% CI: (50, 95)%.

**Sampling & Trends** While levels of condom use reported by men and women do not always agree, the levels should agree in simulated partnerships. To reflect uncertainty due to the discrepancy, we sampled condom use for each year and partnership type from BAB prior distributions having 95% CI that spans the range of estimates from men and women (where applicable), including the widest points of all confidence intervals. We further expanded the confidence intervals in some cases by enforcing a maximum value of $N = 100$ for the BAB distribution. We assume that condom use was effectively zero in 1980 [56]. We also assume and enforce two conditions that: condom use must be monotonic increasing over time; and condom use must be highest in new sex work partnerships, and lowest in main partnerships, for all sampled parameter values. For each available year, we simultaneously sample condom use for all partnership types, and samples failing the condition are discarded. As illustrated in § A.4.1, this sampling strategy minimizes differences between the prior and sampled-with-constraint distributions. For each partnership type, we then smoothly interpolate between sampled levels of condom use over the available years using monotone piecewise cubic interpolation [60].

[7] "Higher risk" partners were defined in [2] as: "*Sexual intercourse with a partner who was neither a spouse nor lived with the respondent*", effectively matching the model definition of "casual" partnerships.

[8] We integrated the reported confidence intervals using the delta method after assuming binomial-distributed proportions.

###### A.3.5.3 Genital Ulcer Disease

Self-reported prevalence of GUD in p12m among sexually active women and men aged 15–49 was approximately 7% in 2006 [2, Table 13.14]. This prevalence was not stratified by numbers of partners, so we modelled GUD prevalence among the lowest risk women and men as 7%. Among the medium risk groups, we sampled GUD prevalence uniformly between 7% and the prevalence modelled among lower risk FSW (below).

The 2011 and 2014 FSW surveys did not ask respondents about GUD specifically, but about any STI symptoms in p12m.[9] In the wider population [2], approximately 60% of women self-reporting any STI symptoms specifically reported GUD in p12m; thus, self-reported STI symptoms among FSW may overestimate p12m GUD prevalence. Approximately 50% and 25% of FSW reported STI symptoms in 2011 and 2014, respectively. Reflecting uncertainty related to self-reported estimates, STI *vs* GUD, and sampling bias, we sampled p12m GUD prevalence among lower risk FSW from a BAB distribution with 95% CI (20, 40)%. Per analysis in § A.3.9, we assumed that STI (and thus GUD) prevalence was approximately 1.3 (1.0, 1.6) times higher among higher risk FSW (gamma prior). FSW data also suggest declining STI prevalence between 2011 and 2014, which could reflect scale-up of STI testing and treatment [61]. However, STI prevalence among Swati youth in 2017–18 remained high [62]. Thus, to reflect uncertainty in STI/GUD prevalence trends, we sampled a relative reduction in GUD prevalence for all populations between 2010 and 2030 from a uniform distribution spanning [0.2, 1].

Finally, no Eswatini-specific data are available for clients of FSW, but studies in Zimbabwe [63], Senegal [64] and Zambia [53] have found 2.5–3.7 (95% CI span 1.4–5.0) the odds of STI symptoms during the past 6–12 months among clients *vs* non-clients. Thus, we defined GUD prevalence among lower risk clients as midway between medium risk groups and lower risk FSW, and among higher risk clients as equal to lower risk FSW.

##### A.3.6 HIV Progression & Mortality

###### A.3.6.1 HIV Progression

The length of time spent in each HIV stage is related to rates of progression between stages $\eta_h$, rates of additional HIV-attributable mortality by stage $\mu_{\text{HIV},h}$, and treatment via antiretroviral therapy (ART). Lodi et al. [65] estimate median times from seroconversion to CD4 < 500, < 350, and < 200 cells/mm$^3$, while Mangal [6] directly estimate the rates of progression between CD4 states $\eta_h$ in a simple compartmental model. Based on these data, we modelled mean durations ($1/\eta_h$) of:[10] 0.142 years in acute infection ($h = 2$, from § A.3.4.1); 3.35 years in CD4 > 500 ($h = 3$); 3.74 years in 350 < CD4 < 500 ($h = 4$); and 5.26 years in 200 < CD4 < 350 ($h = 5$); plus the remaining time until death in CD4 < 200 ($h = 6$, AIDS). Since the duration in acute infection ($h = 2$) is randomly sampled, the remaining duration in CD4 > 500 ($h = 3$) is adjusted accordingly.

[9] The survey question about STI symptoms was: "*In the last 12 months, have you had symptoms of a sexually transmitted infection including discharge from your vagina or sores on or around your vagina or anus*".
[10] Assuming exponential distributions for durations in each CD4 state [66].

###### A.3.6.2 HIV Mortality

Mortality rates by CD4-count in the absence of ART were estimated in multiple African studies [6,67,68]; based on these data, we estimated yearly HIV-attributable mortality rates $\mu_{\text{HIV},h}$ as: 0 during acute phase ($h$ = 2); 0.4% during CD4 > 500 ($h$ = 3); 2% during 350 < CD4 < 500 ($h$ = 4); 4% during 200 < CD4 < 350 ($h$ = 5); and 20% during CD4 < 200 ($h$ = 6, AIDS).

##### A.3.7 Antiretroviral Therapy

Viral suppression via antiretroviral therapy (ART) influences the probability of HIV transmission, as well as rates of HIV progression and HIV-related mortality. The model considers individuals on ART before ($c$ = 3) and after ($c$ = 4) achieving full viral load suppression (VLS), as defined by undetectable HIV RNA in blood samples. Among retained patients initiating ART (see § A.3.8.2 for rates), time to VLS is usually described as "within 6 months" [69]. Mujugira et al. [70] estimated the median time to VLS as 3.1 [IQR: 2.8, 5.5] months from 1592 HIV serodiscordant couples; however this time may be underestimated due to the trial conditions and population. The distribution of time to VLS (Figure 1 in [70]) also featured a heavy tail, suggesting heterogeneity in time to VLS (see § A.3.8.1 for implications). For example, time to VLS may be prolonged due to social and economic barriers to care [71,72]. Considering these data, we sampled the time to VLS (duration in cascade state $c$ = 3) from a gamma distribution with 95% CI (0.33, 1.0) years.

###### A.3.7.1 Probability of HIV Transmission on ART

All available evidence suggests that viral suppression by ART to undetectable levels prevents HIV transmission, *i.e.*, undetectable = untransmittable ("U=U") [73]. Thus, we assumed zero HIV transmission from individuals with VLS ($c$ = 4). However, HIV transmission may still occur during the period between ART initiation to viral suppression ($c$ = 3) [70]. Donnell et al. [30] estimate an adjusted incidence ratio of 0.08 (0.0, 0.57) for all individuals on ART. However, in [30] and [74], the 1 and 4 (respectively) genetically linked infections from individuals on ART all occurred within 90 days of ART initiation, suggesting that risk of transmission only persists before viral suppression. Adjusting the incidence denominator (person-time) to 90 days per individual who initiated ART in [30] results in approximately 3.13 times higher estimated incidence ratio: 0.25 for this specific period.[11] Thus, we sampled relative infectiousness on ART but before viral suppression ($c$ = 3) from a BAB distribution with mean (95% CI) of 0.25 (0.01, 0.67). Finally, we assumed that the virally un-suppressed state ($c$ = 5) had half the reduced infectiousness of $c$ = 3, yielding 95% CI: (0.50, 0.83).

###### A.3.7.2 HIV Progression & Mortality on ART

Effective ART stops CD4 cell decline and results in some CD4 recovery [75,76]. Most CD4 recovery occurs within the first year of treatment [75]. Due to the limited number of modelled treatment states, we model

[11] In [30], individuals who initiated ART contributed approximately 9.4 months per-person (273 persons / 349 person-years, Tables 2 and 3); thus the first 3 months of each individual represent 3/9.4 = 0.319 fewer person-months of follow-up.

this initial recovery to be associated with the pre-VLS ART state ($c = 3$). Lawn et al. [76] and Gabillard et al. [77] estimate an increase of between 25–39 cells/mm$^3$ per month during the first 3 months of treatment. After initial increases, CD4 recovery is modest and plateaus. Battegay et al. [75] report approximate increases of 22.4 cells/mm$^3$ per year between years 1 and 5 on ART. Since HIV states $h = 4, 5, 6$ correspond to 150, 150, and 200-wide CD4 strata, we model rates of movement along $h = 6 \longrightarrow 5 \longrightarrow 4 \longrightarrow 3$ as 0.167, 0.167, 0.125 per month, respectively, during pre-VLS ART ($c = 3$) and 0.1 per year after VLS ($c = 4$).

Since higher CD4 states are modelled to have lower mortality rates (see § A.3.6.2), the modelled recovery of CD4 cells via ART described above implicitly affords a mortality benefit. However, HIV infection is associated with increased risk of death by non-AIDS causes — *i.e.*, unrelated to CD4 count — including cardiovascular disease and renal disease [78]. Lundgren et al. [79] estimated 61% reduction in non-AIDS life-threatening events due to ART. For the same CD4 strata, Gabillard et al. [77] also report approximately 2-times higher mortality rates within the first year of ART *vs* thereafter, suggesting that VLS is associated with 50% mortality reduction independent of CD4 increase. Thus, we modelled an additional 50% reduction in mortality among individuals with VLS ($c = 4$), and half this (25%) reduction before achieving VLS ($c = 3$).

##### A.3.8 Rates of HIV Diagnosis, ART Initiation, Viral Un-suppression & Re-suppression

Rates of HIV diagnosis $\delta$, ART initiation $\tau$, viral un-suppression $\zeta$ (including treatment failure, discontinuation, or loss to follow-up), and viral re-suppression $\sigma'$ (Figure A.1c) were defined to reflect historical trends and ART eligibility for Eswatini [7–10], as described in detail below. These rates were further calibrated to reproduce observed cascade attainment over time in Eswatini (*e.g.*, proportion on ART among those diagnosed with HIV). Similar to condom use, rates were interpolated between specified years using monotone piecewise cubic interpolation [60].

###### A.3.8.1 HIV Diagnosis

Multiple Eswatini studies report the proportions of women and men who tested for HIV in the p12m. However, this proportion may not directly reflect the yearly rate of diagnosis, because individuals may test more frequently based on their perceived risk [80]. Indeed, EmaSwati living with HIV were more likely to have reported previously testing for HIV in 2006 [2, Table 14.9], 2011 [81, Table 5], and 2016 [5, Table 7.3]. Additionally, the proportion tested in p12m likely underestimates the *rate* of testing due to repeat testers. Assuming an exponentially-distributed time spent untested in the period under consideration (consistent with inherent compartmental modelling assumptions), the testing rate $\lambda$ can be calculated from the proportion tested $\rho$ over period $T$ via:

$$\begin{aligned} \rho &= 1 - \exp(-\lambda T) \\ \lambda &= -\log(1 - \rho)/T \end{aligned} \tag{A.14}$$

Moreover, [55] found approximately 70% underreporting of ever testing for HIV in face-to-face interviews *vs* anonymous polling booth surveys, with consistent results across married and unmarried women and men.

Yet, preliminary model calibration using reported HIV testing rates (with 95% CI) described below as HIV diagnosis rates directly caused the model to overestimate HIV+ status awareness *vs* the available data (see § A.4.2.3, Table A.10). This apparent discrepancy between reported population-level testing rates and HIV+ status awareness is in fact common, and could be explained by testing rate heterogeneity [82] — *i.e.*, the existance of "fixed" sub-populations who test frequently and those who test rarely or never. Without further stratifying the modelled population along this testing frequency dimension, it is impossible to capture this heterogeneity directly. However, an alternative solution is to reduce modelled HIV diagnosis rates to reproduce the available data on HIV+ status awareness via model calibration. To this end, we parameterized HIV diagnosis rates over time based on reported testing rates (below), with a global reduction factor $f \sim \text{Unif}(0.5, 1)$. We further specified diagnosis rates using non-FSW women as a reference group, with separate time-varying *relative* rates defined for FSW and men. Confidence intervals for relative rates were assumed using a standard deviation of 0.2 for FSW and 0.1 for men (gamma priors).

**HIV Testing Rates** Early HIV testing in Eswatini was mainly available to pregnant women via antenatal clinics, though a small number of youth and men also accessed HIV testing services [83,84]. Based on antenatal clinic data [85], we modelled a gradual increase in rates of HIV diagnosis among women from zero to 95% CI (5, 15)% (gamma prior) per year from 1990 to 2002, when the national HIV testing and counselling program was formally introduced [61]. We assumed no initial differences between FSW and other women, due to the lack of specific key populations prevention programs [86]. We further assumed that HIV diagnosis among men initially occurred at 10% the rate of women.

By 2006, $\rho$ = 21.9 (20.6, 23.3)% of women and 8.9 (7.8, 10.0)% of men had tested for HIV and received the results in p12m [2][12] — relative rate for men *vs* women: 0.377 (0.207, 0.597). Further scale-up of HIV testing began in 2006 via provider-initiated testing and improved integration with the general health care system [61]. Between 2007 and 2010, such efforts doubled the number of testing locations (119 to 241) and tripled the number of total yearly tests (53,000 to 154,000) [61,87]. By 2011, an estimated $\rho$ = 46.8% of women, 28.4% of men, and 61.7 (55.6, 67.5)% of FSW had tested for HIV in p12m [18,81],[13] yielding testing rates of $\lambda$ = 0.631, 0.333, and 0.962 per year, respectively — relative rates: 0.529 (0.352, 0.743) for men, and 1.521 (1.206, 1.980) for FSW.

Phase 1 of the MaxART program [88] ran from 2011 to 2014, with a primary objective to increase HIV testing. An estimated 284,680 people were reached with 389,658 tests by the end of Phase 1 (2014). By 2016, 57.1% of women and 47.8% of men had tested in p12m [5], yielding testing rates of $\lambda$ = 0.846 and 0.650 per year, respectively. The relative rate for men increased to 0.770 (0.587, 0.978); however, this increase was *not* applied (2011 relative rate maintained) to improve model fit (see § A.4). In 2014 [19] and 2020 [22] approximately $\rho$ = 75% of FSW had tested in p12m ($\lambda$ = 1.386) as such, we applied a relative rate of 1.62, (1.29, 2.07) for 2016. We held all rates of HIV diagnosis after 2016 fixed.

[12] Unless otherwise noted, "*tested for HIV*" will imply "*and received the results*" throughout this section.
[13] The adjustment for missing ages 15–17 in [81] from § A.4.2.1 was applied to the reported 50.1% of women and 31.7% of men aged 18–49 who tested in p12m, assuming 20% of women and 10% of men aged 15–17 tested in p12m.

###### A.3.8.2 ART Initiation

Rates of ART initiation $\tau$ were modelled to reflect time-varying eligibility, availability, loss to follow-up, and differences between sex/activity groups.

**Eligibility** Historical ART eligibility in Eswatini has generally followed the evolving World Health Organization (WHO) guidelines [89–92]. Initial eligibility included one of [7]:

- CD4 < 200 cells/mm$^3$ and any WHO clinical stage
- CD4 < 350 cells/mm$^3$ and WHO clinical stage III
- any CD4 count and WHO clinical stage IV

Eligibility was revised in 2010 [8] to:

- CD4 < 350 cells/mm$^3$ and any WHO clinical stage
- any CD4 count and WHO clinical stage III or IV

and again in 2015 [9] to:

- CD4 < 500 cells/mm$^3$ and any WHO clinical stage
- in a discordant partnership or having a specified illness (any CD4 count or WHO clinical stage)

before adoption of the current "*ART for all*" guidelines in late 2016 (modelled as effectively January 2017) [10,93]. Phase 2 of MaxART also began in 2015, offering immediate ART via 14 health facilities in a stepped wedge design (6 facilities added per year) [93]. Relative to the 114 total facilities offering ART nationally at this time [94], we assumed this trial had minimal direct impact on population-level ART initiation — notwithstanding valuable insights gained regarding effective implementation [93].

We implemented the CD4-only eligibility criteria directly in the model, which is structured to match these 200, 350, and 500 CD4 cells/mm$^3$ thresholds (Figure A.1b). For eligibility by WHO clinical stages (not explicitly modelled), we estimated relative rates of ART initiation based on the following data from South Africa [95, Table 4] and Saudi Arabia [96, Table 2], respectively:

- 43/111 (39%) and 14/46 (30%) of PLHIV with 200 < CD4 < 350 were at stages III or IV;
  assumed: 35% PLHIV with 200 < CD4 < 350 were eligible for ART pre-2010
- 13/79 (16%) and 6/76 (8%) of PLHIV with CD4 > 350 were at stage III;
  assumed: 15% PLHIV with 350 < CD4 < 500 were eligible for ART pre-2010 (5% with CD4 > 500)
- 5/79 (6%) and 1/76 (1%) of PLHIV with CD4 > 350 were at stage IV;
  assumed: 20% PLHIV with 350 < CD4 < 500 were eligible for ART 2010–2015 (5% with CD4 > 500)

We assumed that roll-out of eligibility changes in 2010, 2015, and 2017 each occurred over a 1-year period. Figure A.5 illustrates the resulting modelled relative rates of ART initiation for each HIV stage over time.

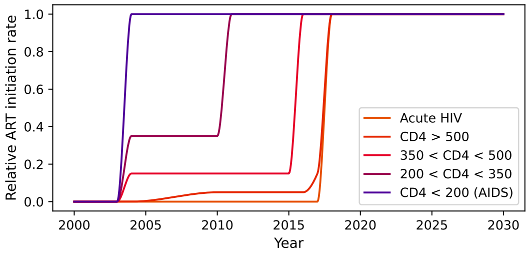

Figure A.5: Modelled relative rates of ART initiation by HIV stage, reflecting eligibility changes over time

**Availability and Initiation** ART first became available in Eswatini in late 2003 via a one-hospital pilot project [61]. Early ART scale-up was modest, with 31 facilities offering ART by the end of 2009 [97]; however, this number increased rapidly to 110 facilities by the end of 2011 [61]. Phase 1 of MaxART (2011–2014) sought to further increase ART coverage among eligible PLHIV [88], including decentralization to lower level facilities, bringing the total number of facilities to 170 by 2015 [98]. Finally, national adoption of "*Test and Start*" in 2017 likely further reduced delays in ART initiation, while loss to follow-up was reduced throughout the years of ART scale-up [93].

Considering these data, we modelled the yearly ART initiation rate among eligible diagnosed PLHIV as: effectively $\tau = 0$ in 2003, gradually increasing to 1.5 (0.5, 3.0) by 2010; then to 9 (6, 12) by 2012; and stabilizing at 12 by 2018. This maximum rate of $\tau = 12$ corresponds to a mean effective delay of one month between diagnosis and ART initiation; this value was chosen in part to avoid numerical instability when solving the model with very high rates.

**Group Differences** In 2011, conditional ART coverage (among diagnosed) was greater among men *vs* women (Table A.10), suggesting greater ART initiation among men *vs* women. Yet, unconditional ART coverage (among PLHIV, regardless of diagnosis) were approximately equal (31.4 and 33.2%, respectively), and so conditional differences may be explained by the fact that women were more likely to be diagnosed at an earlier HIV stage via antenatal care, and thereafter not yet eligible for ART. Thus, we assumed no differences in ART initiation among men *vs* women. A similar mechanism could partially explain differences in conditional coverage between FSW *vs* women overall (36.9 *vs* 48.0%), as FSW were more slightly likely to know their status (74.1 *vs* 69.1%). However, FSW face unique barriers to accessing ART related to stigma and material insecurity [99]; as such, we sampled a relative rate for ART initiation among FSW from [0.5, 1] (uniform prior).

###### A.3.8.3 ART Failure

The modelled virally un-suppressed state ($c = 5$) reflects any combination of treatment failure (*i.e.*, due to resistance mutations), discontinuation, or loss to follow-up (LTFU) after achieving viral suppression. The model does not explicitly simulate emergence and/or transmission of drug resistance, nor multiple unique ART regimens. As of 2016, resistance mutations to at least 1 of 3 drugs in combination regimens were identified in 10% ART-naive PLHIV in Eswatini, and 16% PLHIV with prior ART exposure [100]. However, the extent to which these individual mutations can cause complete treatment failure remains unclear. Additionally, while transmissible resistance mutations could become more prevalent over time, emergence of new drugs can combat the population-level impacts of this resistance [101].

All available data suggests that retention in ART care — *i.e.*, not discontinued or LTFU — has improved over time in Eswatini [5,102,103]. Assuming an exponentially-distributed retention time (consistent with inherent compartmental modelling assumptions), we averaged the available data [103, Table 6] to calculated the effective yearly ART attrition rate as: 16.5% in 2008, 13.8% in 2010, 14.1% in 2012, and 8.3% in 2014. One-year LTFU was reported as 1% in 2016 [5], but it's not clear whether this definition was consistent with the earlier estimates. Many measures of LTFU may also overestimate true LTFU by failing to account for transfers between clinics and deaths [104,105]; it's not clear whether the reported measures for Eswatini account for transfers or deaths.

LTFU was estimated to be 1.3 times higher among men *vs* women in South Africa [104], which would be consistent with observed lower viral suppression among men *vs* women on ART in Eswatini (Table A.10) [104]. The same study estimated that LTFU did not significantly differ by the modelled CD4-strata [104]. No estimates of LTFU were available for FSW specifically in Eswatini, but among 354 FSW on ART in [22] (2021), 103 knew the results of viral load monitoring in p12m, of whom only 8 self-reported undetectable viral load. Such data may again reflect the unique barriers to accessing ART faced by FSW [99].

Considering all of the above data, assumed: a yearly rate of viral un-suppression $\zeta$ among non-FSW women of 15% until 2010, decreasing to 5% by 2018; plus relative rates for men and FSW: [1, 1.5] (uniform priors).

###### A.3.8.4 Viral Re-suppression

The rate of viral re-suppression $\sigma'$ aims to reflect the average delay associated with the steps of switching regimens (in case of treatment failure), or the steps of re-engaging in HIV care (in case of LTFU).

For treatment failure, viral un-suppression must first be identified. Availability of viral load monitoring in Eswatini was limited until at least 2010 [8], but incorporated into standard of care by 2015 (yearly testing) [9]. Without viral load testing, treatment failure can still be indicated clinically [8]. After suspecting treatment failure, at least three months of additional monitoring is typically required to rule-out issues of adherence [8–10], before another regimen is started. Moreover, second/third-line regimen options were limited in Eswatini until at least 2014 [94,106]. Upon switching to an improved regimen, assume that viral suppression occurs at the same rate as among ART-naive PLHIV (see § A.3.7).

For LTFU, no data directly indicate the average duration out of care in Eswatini. A recent model-based analysis of Kenyan data [107] suggests an average between 8 months and 2 years. Considering large-scale, multisectorial efforts to improve ART care in Eswatini, it is likely that duration out of care has declined since 2010. Thus, sampled the initial rate of viral re-suppression $\sigma'$ from a gamma prior with 95% [0.5, 1.0], which increased by a factor of 1.5 over 2010–2018. We assumed no differences between groups.

##### A.3.9 Risk Differences Within Sex Work

Compartmental HIV transmission models which include FSW have rarely sub-stratified FSW (besides age) [108–111], such as to reflect differential HIV risk or distinct typologies of sex work [112,113]; yet such heterogeneities may influence transmission dynamics. Our model structure (Figure A.1a) was designed to capture *within*-FSW risk heterogeneity. The objective of the following analysis was therefore to parameterize higher *vs* lower risk sex work. For this analysis, we used individual-level data from two biobehavioural surveys among Swati FSW in 2011 [18,24] (N = 325) and 2014 [19] (N = 781). More details about each study are given in § A.3.2.

Based on community input,[14] we conceptualized risk differences within sex work as transient periods of higher *vs* lower risk, rather than distinct types of sex worker. As such, we modelled rapid turnover between higher and lower risk FSW (see § A.3.12), and distinguished these states via the total numbers of clients in p1m. Specifically, we stratified survey respondents into the top 20% / bottom 80% in total numbers of new and regular clients reported for p1m. We then summarized key variables within the two strata (Table A.4) and estimated the ratio of means per [114]. We repeated this analysis using 2011, 2014, and combined datasets.

Years selling sex, non-paying partners, condom use, anal sex, and HIV status (2011 data only) did not differ substantially between strata, while reported numbers of clients and STI symptoms did. We sampled reported numbers of clients in p1m from gamma distributions with $\alpha = 25$, reflecting an assumption of mean / 5 = standard deviation; means were specified as: 14 and 21 for new and regular clients in higher risk sex work, and 3.5 and 6 for new and regular clients in lower risk sex work. We further use data in Table A.4 regarding: partners and clients in § A.3.13, STI symptoms in § A.3.4.5, condom use in § A.3.5.2, anal sex in § A.3.14, years selling sex in § A.3.12.

##### A.3.10 Wider Population: Bias Adjustment

We stratified the remaining women and men (besides FSW and clients) by numbers of partners in the past 12 months (p12m): 0–1 and 2+. The 2006-07 DHS [2], 2011 SHIMS [20], and 2016-17 SHIMS2 [5] surveys provide the numbers of respondents who reported 2+ partners in the past 12 months (p12m): 13.5, 18.2, 14.5% among men, and 1.6, 3.8, 4.1% among women, respectively.[15] However, such reports are likely substantially biased by social desirability bias due to the face-to-face interview format [55,115–117]. Moreover, these data do not provide information on the *types* of partners reported — *i.e.*, those reporting 1 partner in p12m are not necessarily in a main/spousal (vs casual) partnership, and neither are those reporting 2+ partners in p12m. Here we develop and apply a new method to adjust for these potential reporting biases, and simultaneously estimate the numbers of main/spousal and casual partners among each stratum. The results of these analyses then directly inform activity group sizes in § A.3.11.3 and numbers of partners in § A.3.13.2.

[14] Personal communication: Lungile Khumalo, *Voice of Our Voices*, Eswatini

[15] From Tables 14.7.1 and 14.7.2 (ages 15-49) in [2], Table 3 (ages 18-49) in [20], Table 15.3.A (ages 15+) in [5], with manual adjustment for survey skip patterns in [2,5].

Table A.4: Ratios of variables among higher *vs* lower risk FSW in Eswatini

| | | Higher | | Lower | | Ratio | |
|---|---|---|---|---|---|---|---|
| Year | Variable | mean | (range) | mean | (range) | mean | (95% CI) |
| 2011 | Age | 24.5 | (17, 41) | 26.6 | (16, 49) | 0.92 | (0.87, 0.98) |
| | Years selling sex | 4.82 | (0, 18) | 5.76 | (0, 30) | 0.84 | (0.63, 1.06) |
| | Non-paying partners p1m | 1.32 | (0, 5) | 1.45 | (0, 6) | 0.91 | (0.71, 1.12) |
| | New clients p1m | 18.9 | (0, 60+) | 3.87 | (0, 15) | 4.87 | (3.55, 6.27) |
| | Regular clients p1m | 23.9 | (3, 60+) | 5.71 | (0, 20) | 4.19 | (3.40, 5.04) |
| | Non-paying partner condom use [a] | 0.51 | — | 0.49 | — | 1.04 | (0.74, 1.39) |
| | New client condom use [a] | 0.93 | — | 0.87 | — | 1.07 | (0.98, 1.17) |
| | Regular client condom use [a] | 0.78 | — | 0.83 | — | 0.94 | (0.80, 1.08) |
| | Any anal sex p1m [a] | 0.37 | — | 0.47 | — | 0.79 | (0.50, 1.11) |
| | Any STI symptoms p12m [a] | 0.60 | — | 0.48 | — | 1.24 | (0.95, 1.57) |
| | HIV status [ab] | 0.72 | — | 0.70 | — | 1.03 | (0.85, 1.22) |
| 2014 | Age | 27.1 | (18, 44) | 27.6 | (18, 50) | 0.98 | (0.95, 1.01) |
| | Years selling sex | 6.12 | (0, 22) | 6.44 | (1, 26) | 0.95 | (0.83, 1.08) |
| | Non-paying partners p1m | 1.43 | (0, 17) | 1.15 | (0, 10) | 1.25 | (0.91, 1.61) |
| | New clients p1m | 12.2 | (0, 60+) | 3.35 | (0, 16) | 3.65 | (3.10, 4.23) |
| | Regular clients p1m | 20.0 | (0, 60+) | 6.21 | (0, 20) | 3.22 | (2.89, 3.58) |
| | Non-paying partner condom use [a] | 0.71 | — | 0.83 | — | 0.85 | (0.73, 0.98) |
| | New client condom use [a] | 0.89 | — | 0.89 | — | 1.00 | (0.93, 1.07) |
| | Regular client condom use [a] | 0.82 | — | 0.87 | — | 0.94 | (0.86, 1.02) |
| | Any anal sex p1m [a] | 0.13 | — | 0.08 | — | 1.69 | (0.93, 2.70) |
| | Any STI symptoms p12m [a] | 0.30 | — | 0.22 | — | 1.34 | (0.98, 1.77) |
| Both | Age | 26.2 | (17, 44) | 27.3 | (16, 50) | 0.96 | (0.93, 0.99) |
| | Years selling sex | 5.63 | (0, 20) | 6.27 | (0, 30) | 0.90 | (0.80, 1.01) |
| | Non-paying partners p1m | 1.39 | (0, 17) | 1.25 | (0, 10) | 1.12 | (0.90, 1.35) |
| | New clients p1m | 14.2 | (0, 60+) | 3.53 | (0, 20) | 4.03 | (3.45, 4.63) |
| | Regular clients p1m | 21.2 | (0, 60+) | 6.05 | (0, 20) | 3.51 | (3.18, 3.85) |
| | Non-paying partner condom use [a] | 0.62 | — | 0.71 | — | 0.88 | (0.76, 1.01) |
| | New client condom use [a] | 0.90 | — | 0.88 | — | 1.02 | (0.96, 1.08) |
| | Regular client condom use [a] | 0.80 | — | 0.86 | — | 0.94 | (0.87, 1.01) |
| | Any anal sex p1m [a] | 0.19 | — | 0.19 | — | 1.01 | (0.70, 1.36) |
| | Any STI symptoms p12m [a] | 0.40 | — | 0.30 | — | 1.33 | (1.08, 1.61) |
| | HIV status [ab] | 0.75 | — | 0.69 | — | 1.07 | (0.90, 1.27) |

Higher / Lower: top 20% / bottom 80% by total clients p1m; [a] proportion of respondents; [b] 2011 data only (serologic HIV status).

###### A.3.10.1 Reported Partner Numbers

Both the 2006 DHS [2, Tables 14.6.1 and 14.6.2] and 2016-17 SHIMS [5, Tables 15.4.A and 15.4.B] summarize the numbers of women and men by partners in p12m *and* by marital/union status, but summaries are stratified by each factor separately, not jointly. However, making the following assumptions, we estimated the jointly-stratified proportions of individuals. Let $W_{2+}$, $W_1$, and $W_0$ denote women reporting 2+, 1, and 0 partners, respectively, and likewise with $M_{2+}$, $M_1$, $M_0$ for men (all partners reflect p12m). The assumptions were:

- $W_{2+}$ included all women in non-polygynous unions (married or cohabiting) reporting sex with a "casual" (non-marital, non-cohabiting) partner
- $M_{2+}$ included all men in polygynous unions, plus all men in non-polygynous unions reporting sex with a casual partner
- the remaining $W_{2+}$ and $M_{2+}$ formed only casual partnerships
- all women and men in non-polygynous unions reporting no sex with a casual partner reported 1 partner ($W_1$ and $M_1$)
- the remaining $W_1$ and $M_1$ formed only casual partnerships

Figure A.6 illustrates the resulting proportions of women and men in each union / partners in p12m stratum in 2006-07 (a) and 2016-17 (b).

###### A.3.10.2 Bias Adjustment: Rationale

We were motivated to examine potential reporting bias because $M_{2+}$ is consistently much greater than $W_{2+}$. In fact, this difference is common in surveys [118,119], and could be explained by either: (a) a small number of women with many partners, such as FSW, who may also not be reached by the survey, or who may not fully report partner numbers; (b) over-reporting of partnerships by men; or (c) under-reporting of partnerships by women. Further stratification of women reporting 2+ partners in [2, Table 14.7.1] revealed that 94% reported exactly 2 whereas 6% reported 3+, suggesting that explanation (a) is less likely unless women with 3+ partners are under-reported or indeed missing from the survey.

Gregson et al. [120] (Zimbabwe), Nnko et al. [121] (Tanzania) and Clark, Kabiru, and Zulu [122] (Kenya) explored explanations (b) and (c) through measures of consistency; their results suggested that under-reporting of non-spousal partnerships by women (c) was more likely, perhaps due to social norms and pressures. Such norms in Eswatini are explored in [123–126]. In fact, a review comparing computer-based tools *vs* face-to-face interviews for surveying sexual behaviour [127] found that *both* women and men may under-report sexual partners, but women more so. A notable 2008 study in Benin [55] found that 7 times as many married women (21 *vs* 3%) and 3 times as many married men (53 *vs* 18%) reported any extramarital sex in p12m in a survey via anonymous polling booth *vs* face-to-face interview. Similarly, 5 times as many unmarried women (13.5 *vs* 2.8%) reported exchanging sex for money, gifts or favours in p12m, while 4 times as many unmarried men (62 *vs* 14%) reported non-transactional sex with a women in p12m. Such findings were similar to those from Zimbabwe (1990s) [120].

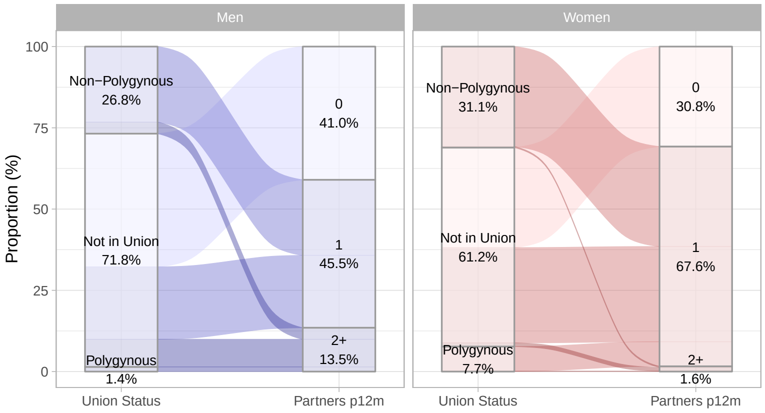

(a) 2006-07 [2]

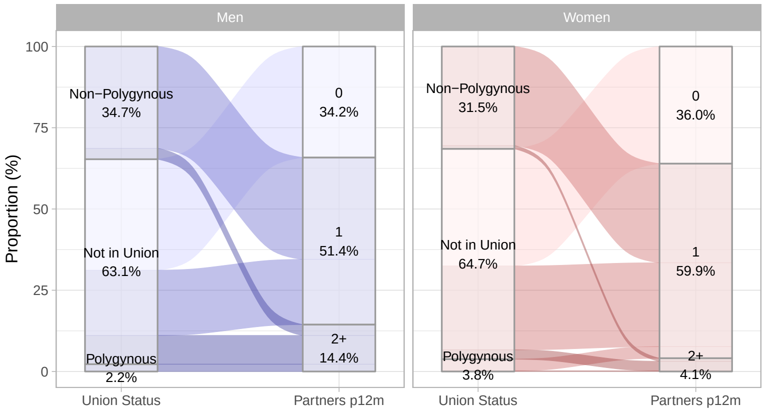

(b) 2016 [5]

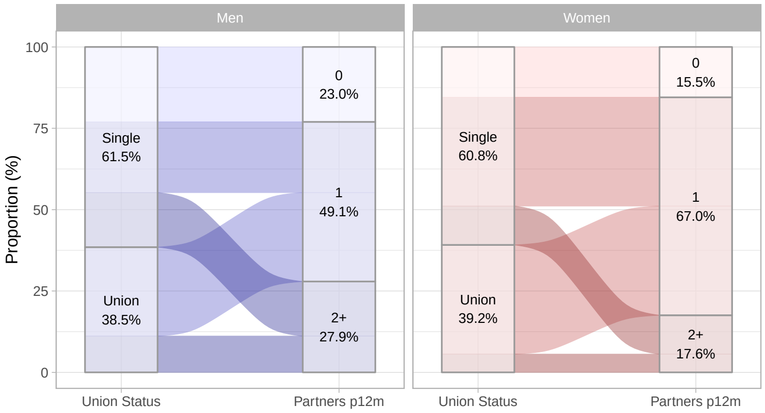

(c) Adjusted (mean)

Figure A.6: Reported proportions of women and men aged 15–49, stratified by union status and numbers of partners in the past 12 months

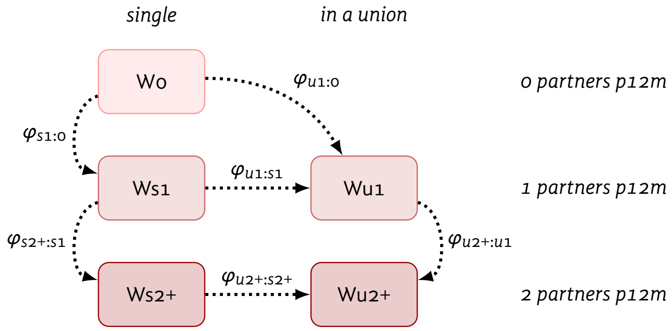

Figure A.7: Illustration of how the proportions of women (and equivalently men) are adjusted / reallocated between union / partners in p12m strata based on odds ratios $\varphi$

p12m: within the past 12 months; W0: 0 partners in p12m; Ws1: single (not married/cohabiting) and 1 partner in p12m; Wu1: in a union (married/cohabiting) and 1 partner in p12m; Ws2+: single and 2+ partners in p12m; Wu2+: in a union and 2+ partners in p12m. $\varphi$: odds of truly being in the second (arrowhead) *vs* first (tail) group.

###### A.3.10.3 Bias Adjustment: Approach

To account for the above potential reporting biases and qualitative insights from [123–126], We modelled the adjusted proportions of Swati women and men in each union / partners in p12m stratum as follows. Let $W_{s1}$ and $W_{u1}$ denote sub-proportions of $W_1$ who are single and in a union, respectively, and likewise for $W_{s2+}$, $W_{u2+}$, $M_{s1}$, $M_{u1}$, $M_{s2+}$, and $M_{u2+}$. Further, let $W_{s1}$ denote the reported proportion of women (average of 2006-07 and 2016-17), *vs* $W'_{s1}$ denoting the adjusted proportion. We assumed that a faction of $W_0$ belongs in $W'_{s1}$ — *i.e.*, a fraction of women reporting 0 partners in p12m truly had 1 casual (non-main/spousal) partner. We modelled this relationship through an odds ratio $\varphi_{W,s1:0}$, which is roughly equivalent in interpretation to the proportion ratios estimated by Béhanzin et al. [55]:[16]

$$\varphi_{Ws1:0} = \frac{W'_{s1}}{W'_0} \bigg/ \frac{W_{s1}}{W_0} \tag{A.15}$$

We defined similar odds ratios $\varphi_{Ws2+:s1}$, $\varphi_{Wu2+:u1}$, $\varphi_{Wu1:0}$, $\varphi_{Wu1:s1}$, and $\varphi_{Wu2+:s2+}$, and likewise for men. These strata and the corresponding adjustments / reallocations of women from reported to adjusted strata are illustrated in Figure A.7. To resolve the adjusted values $W'$ then requires solving the (nonlinear) system of 6 equations corresponding to the 6 odds ratios $\varphi$, subject to $\sum_i W'_i = 1$ and $0 \leq W'_i < 1$. An exact solution is not guaranteed, but the sum squared error from all equations can be minimized. The odds ratios $\varphi$ were then defined as follows, including sampling distributions.

**Union Status** We assumed that under-reporting of main/spousal partnerships was minimal, but that some "main" partnerships may not be captured in the definition "married/cohabiting" from [2,5]; thus $\varphi_{u1:0}$, $\varphi_{u1:s1}$, and $\varphi_{u2+:s2+}$ would be small but greater than 1 (horizontal transitions in Figure A.7). Moreover, based on the median age of marriage, 23–29 [2], approximately half of respondents aged 15–49 would have been married, whereas only 28–39% of women and men reported being in a union (Figure A.6a and A.6b), although

[16] Odds ratios ensure no proportions become greater than one or negative.

some marriages end in divorce/widowing [2]. Thus, we sampled each of $\varphi_{u1:0}$, $\varphi_{u1:s1}$, and $\varphi_{u2+:s2+}$ from 1 + Gamma ($\alpha, \beta$ = 1) with $\alpha$ = .5 for women and $\alpha$ = .3 for men, yielding mean (95% CI): 1.50 (1.00, 3.51) and 1.30 (1.00, 2.90), respectively.

**Partner Numbers** Next, regarding partner numbers, we defined $\varphi_{s1:0}$, $\varphi_{s2+:s1}$, and $\varphi_{u2+:u1}$ as follows (vertial transitions in Figure A.7). The median age of first sex in Eswatini was approximately 18 for women and 19.5 for men [2]. Thus, the 31–36% of women and 34–41% of men aged 15–49 reporting no partners in p12m (Figure A.6a and A.6b) is likely overestimated, although some individuals may be abstinent in p12m following sexual debut. We assumed that women had 3 and men had 2 times the odds of actually having 1 casual partner in p12m while reporting no partners. Thus, we sampled $\varphi_{s1:0}$ from 1 + Gamma ($\alpha, \beta$ = 1) with $\alpha$ = 2 for women and $\alpha$ = 1 for men, yielding mean (95% CI): 3.00 (1.24, 6.57) and 2.00 (1.03, 4.69), respectively. Drawing on [55], we assumed that "single" women and men (not married/cohabiting) were less likely to report multiple partners in p12m, but women more so. Thus, we sampled $\varphi_{s2+:s1}$ from 1 + Gamma ($\alpha, \beta$ = 1) with $\alpha$ = 4 for women and $\alpha$ = 1 for men, yielding 5.00 (2.09, 9.77) and 2.00 (1.03, 4.69). We made a similar assumption about married/cohabiting women and men, with the same odds for men, but even greater odds of non-reporting among women. We sampled $\varphi_{u2+:u1}$ from 1 + Gamma ($\alpha, \beta$ = 1) with $\alpha$ = 6 for women and $\alpha$ = 1 for men, yielding 7.00 (3.20, 12.67) and 2.00 (1.03, 4.69).

###### A.3.10.4 Bias Adjustment: Results

The mean resulting adjusted proportions $W'$ and $M'$ from solving the system with the assumed odds ratios $\varphi$ are illustrated in Figure A.6c, which can be compared to the reported proportions in (a) and (b). Figure A.8 also illustrates the empiric density distributions for each element $W'_i$ and $M'_i$. Numerically, the mean (95% CI) estimates were:

- $W'_0$ = 17 (9, 27)% of women and $M'_0$ = 25 (13, 35)% of men had 0 partners in p12m
- $W'_1$ = 66 (57, 75)% of women and $M'_1$ = 49 (37, 61)% of men had 1 partners in p12m
- $W'_{2+}$ = 17 (10, 27)% of women and $M'_{2+}$ = 26 (15, 44)% of men had 2+ partners in p12m
- $W'_{u1}/W'_{01}$ = 38 (21, 57)% women and $M'_{u1}/M'_{01}$ = 35 (23, 50)% men with 0–1 partners in p12m were in a main/spousal partnership
- $W'_{s1}/W'_{01}$ = 41 (19, 65)% women and $M'_{s1}/M'_{01}$ = 31 (15, 55)% men with 0–1 partners in p12m were in a single casual partnership
- $W'_{u2+}/W'_{2+}$ = 32 (9, 55)% women and $M'_{u2+}/M'_{2+}$ = 38 (13, 62)% men with 2+ partners in p12m were in a main/spousal partnership, and the rest had only casual partnerships.

##### A.3.11 Activity Group Sizes

We model population sizes of all activity groups as proportions of the total population, which are assumed to remain roughly constant. Individuals can, however, move between groups (see § A.3.12.2) — *i.e.*, groups

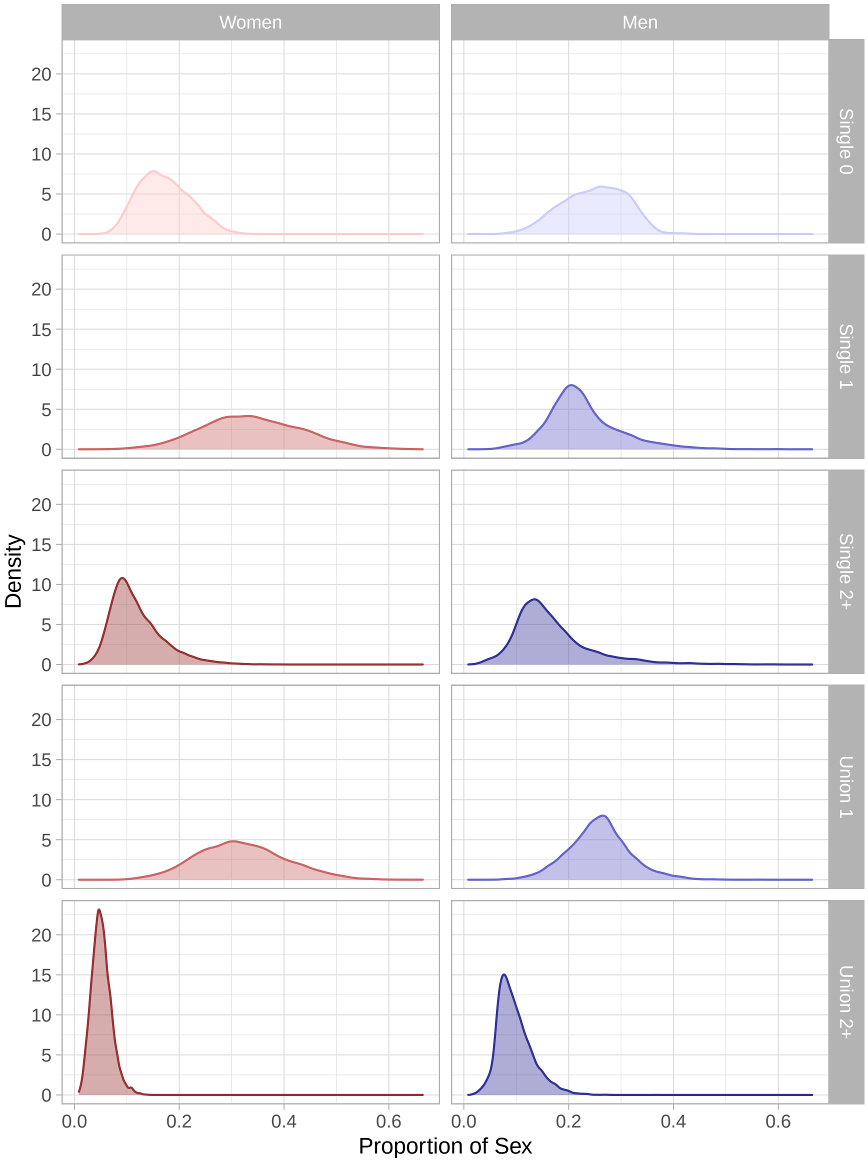

Figure A.8: Density distributions for adjusted proportions of women and men aged 15–49, stratified by union status and numbers of partners in the past 12 months are open populations — and disproportionate mortality due to HIV between groups may cause higher risk groups to shrink over time. Overall population growth is discussed in § A.3.12.1.

###### A.3.11.1 Female Sex Workers

The proportion of women who report sex work in national demographic and health surveys is generally considered unreliable due to social desirability bias, particularly if the survey is face-to-face and household-based [55,115,117,120,128]. Therefore, FSW population size estimates require targeted surveys and unique methodologies [129,130]. In both [19] and [22], the Swati FSW population size was estimated using a combination of unique object method, service multiplier method, prior survey participation, and network scale-up method (NSUM) [129]. In 2011 [19], regional FSW population size estimates ranged from 0.7% to 6.5% of all women, with overall population-weighted mean across regions of 2.9%; in 2021 [22], the mean (95% CI) estimates were 2.43 (1.17, 5.02)%. To reflect this uncertainty in the model, we fit a BAB distribution such that 95% of the probability fell between 0.7% and 6.5%, and used as the prior distribution for the proportion of women who are FSW. Then, following the analysis in § A.3.9, we fixed the proportion of all FSW in the higher risk FSW group at 20%, and likewise the lower risk group at 80%.

###### A.3.11.2 Clients of FSW

Similar to FSW, household-based surveys are not considered reliable data sources for estimating the population size of clients of FSW [55]. However, few surveys are designed to reach clients of FSW, and no direct estimates of FSW size exist for Eswatini. So, we use a common approach for inferring the FSW client size [58], similar to the "multiplier method" [131]. Given the FSW population proportion $P_{FSW}$, the average number of yearly new and regular sex work clients per FSW $\bar{Q}_{FSW}$, the frequency of sex per partnership-year $F_{SW}$, and the average total number of yearly sex acts per client $\bar{Q}_{CLI} F_{SW}$, we define the total client population $P_{CLI}$ as:

$$P_{CLI} = \frac{P_{FSW}\,\bar{Q}_{FSW}\,F_{SW}}{\bar{Q}_{CLI}\,F_{SW}} \tag{A.16}$$

Then, as with FSW, the proportion of total clients in the higher risk client group is defined as 20% of all clients, and likewise for the lower risk group at 80%. Using $\bar{Q}_{FSW}$, $\bar{Q}_{CLI}$, and $F_{SW}$ as defined below in § A.3.13.1, the prior client population size $P_{CLI}$ estimated by this method was 11.6 (2.1, 34.4)% of men.

###### A.3.11.3 Wider Population

Based on the results of § A.3.10, we defined the sizes of the modelled lower and medium activity groups, and the average numbers of main/spousal partnerships per person. We assumed that $W'_{2+}$ and $M'_{2+}$ included FSW and client population sizes, respectively. Thus, we defined the populations size of medium activity women as $W_M = W'_{2+} - W_{FSW}$. Sampling $W'_{2+}$ from a BAB distribution with 95% CI (10, 27)%, the resulting 95% CI for medium activity women $W_M$ was (6, 25)% of women. We then defined the lowest activity women population size as $W_L = 1 - W'_{2+}$, representing (72, 90)% of women. Since there is greater uncertainty in the client population size, the same approach for the medium activity men population size could yield negative values. Instead, we sampled the proportion of medium activity men $M_{\text{M}}$ directly from a BAB distribution with 95% CI (10, 17)%, yielding 95% CI for $M_{\text{M}} + M_{\text{CLI}}$ of (14, 52)% of men, which is close to (15, 44)% from $M_{2+}$. We then defined the lowest activity men were as $M_{\text{L}} = 1 - M_{\text{M}} + M_{\text{CLI}}$, representing (48, 86)% of men.

##### A.3.12 Turnover

###### A.3.12.1 Births & Deaths

The modelled population considers ages 15–49, reflecting commonly reported data and the majority of sexual activity. In the absence of mortality, individuals would therefore remain within the modelled "open cohort" population for 35 years. The estimated average yearly mortality rate for these ages was 1.44% around 2006 [2, Table 15.2]. However, this estimate includes HIV/AIDS-attributable mortality, which we model separately (see § A.3.6.2), accounting for approximately 64% of deaths around that time [132]. Thus, the overall exit rate from the modelled cohort due to reaching age 50 ("aging out") and non-HIV-attributable mortality was: $\mu = 1/35 + (1 - .64)1.44\% = 3.78\%$.

We estimated the rate of entry into the modelled population $\nu$ to fit population size of ages 15–49 in Eswatini [28], and approximate population growth rates [133], given that we model HIV/AIDS-attributable mortality separately. Specifically, we assumed a population growth rate $g = \nu - \mu$ in the absence of HIV/AIDS of 4% in 1980, 3% in 2000, 1.5% in 2010, and 1.5% in 2020 (monotonic cubic interpolation). We sampled $g$ in 2050 from a uniform prior with 95% CI (0.7%, 1.5%), reflecting uncertainty in estimated projections [133]. Finally, we calculated the population entry rate as $\nu = g + \mu$. These parameter values were informally validated by comparison of model outputs with Swati population sizes for ages 15–49 from [28]. The distribution of activity groups among individuals *entering* the model, denoted $E_{si}$, is different from the distribution among individuals *currently* in the model $P_{si}$, but $E_{si}$ is computed automatically as described below in § A.3.12.2.

###### A.3.12.2 Activity Group Turnover

In addition to overall population turnover (entry/exit from the open population), we model movement of individuals between activity groups within the model. Activity group turnover reflects the fact that risk is not constant over sexual life course, and reported duration in higher activity contexts can be short [113]. Previous modelling has shown that activity group turnover (sometimes called "episodic risk") can strongly influence parameter fitting and intervention impact [134,135]. We model turnover from activity group $si$ to $si'$ as a constant rate $\theta_{sii'}$, which implies an assumption that (in the absence of HIV) duration in group $si$ is exponentially distributed with mean $D_{si}$ [66]:

$$D_{si} = \frac{1}{\mu + \sum_{i'} \theta_{sii'}} \tag{A.17}$$

where $\mu$ is the overall exit rate from § A.3.12.1. As shown previously [135], the relative sizes of each sex-activity group $P_{si}$ can be maintained at fixed values by satisfying the following "mass-balance" equation:

$$\nu P_{si} = \nu E_{si} + \sum_{i'} \theta_{si'i} P_{si'} - \sum_{i} \theta_{sii'} P_{si} \tag{A.18}$$

Specific turnover rates $\theta_{sii'}$ and entrant activity group sizes $E_{si}$ can then be uniquely resolved by specifying $N_i\,(N_i - 1) = 12$ non-redundant and compatible constraints, where specifying each $D_{si}$ is one such constraint.

**Selling Sex** Estimating durations (*e.g.*, in sex work) from cross-sectional data should consider several potential sources of bias [136,137], including distributional, sampling, censoring, and measurement biases. We previously explored these biases using the 2011 Eswatini FSW survey data [18] and inferred adjusted estimates of sex work duration via a Bayesian Bayesian hierarchical model [137]. We estimated a mean duration of 4.06 (2.29, 6.34) years, with durations distributed approximately exponentially — compatible with the implicit assumption of compartmental models [138]. Thus, we sampled overall duration in sex work from a gamma prior with 95% CI (2.29, 6.34) years. As noted in § A.3.9, we conceptualized higher risk sex work as a transient period, with short duration. We sampled this duration from a gamma prior with 95% CI (2, 12) months. We then modelled turnover for higher risk sex work as exclusively coming from / going to lower risk sex work, including no direct entry from outside the model: $E_{si} = 0$. By contrast, women entering lower risk sex work could enter directly from outside the model ($E_{si} > 0$), and turnover from / to any other activity group.

**Buying Sex** Data to inform the average duration spent buying sex among clients is limited. Fazito et al. [136] estimated mean durations of 4.6–5.5 years based on studies in Benin [139] and Kenya [140]. Hodgins et al. [141, Table G] also gives pooled estimates for the proportions of men in Sub-Saharan Africa who paid for sex *ever vs* in *p12m* during 2000–2020. Estimates ranged from 8.8 (6.5, 11.7)% of men aged 25–34 who ever bought sex, to 2.2 (1.5, 3.2)% of men aged 35–54 who bought sex in p12m. Based on these data, we defined a gamma prior distribution for the total duration buying sex with 95% CI (4, 15) years. We conceptualized higher risk clients as a transient period with the same duration as higher risk sex work, and assumed an equal pattern of possible turnover between activity groups among men buying sex as women selling sex.

**Lowest & Medium Activity Groups** Data on individual-level changes to numbers of non-sex work partners in p12m is even more sparse than data related to sex work; so, it's unclear to what extent individuals move between the lowest and medium activity groups throughout their sexual life course. Data from Uganda, Zimbabwe, and South Africa [118] suggested that sexual activity (proportion sexually active and mean numbers of partners) was approximately stable with age (after sexual debut and and before age 49), with modest trends toward lower activity at older age. However, these population-level data do not necessarily suggest that the *same* individuals have multiple partnerships each year. Reflecting this uncertainty, we sampled the rate of turnover from medium to lowest activity for both women and men from a gamma prior with 95% CI (5, 50)% per year.

**Additional Turnover Assumptions** The above assumptions specify 8 constraints for each sex: 2 durations $D_{si}$, 1 entry rate $E_{si} = 0$, and 5 turnover rates $\theta_{sii'}$ (4 zero, 1 nonzero). Next, since FSW often enter sex work shortly after sexual debut [142,143], and sexual activity is roughly constant or slightly declining with age

[118], we assumed that $E_{si} = f\,P_{si}$,[17] with $f = 2$ for lower risk FSW, $f = 1.5$ for lower risk clients, and $f = 1$ for medium activity women and men (+2 constraints); then $f < 1$ for the lowest activity women and men is computed automatically. Finally, since exiting sex work is unlikely to be an abrupt transition to monogamous or zero sexual activity [113,144], we further assumed that (50, 90)% of women exiting sex work transition to the medium activity group (BAB prior) (+1 constraint); in the absence of relevant data, we made a similar assumption regarding clients, with (25, 90)% former clients transitioning to the medium activity group (+1 constraint). These 10 < 12 total constraints then allow two degrees of freedom to resolve the values of $\theta_{sii'}$ and $E_{si}$. A non-negative solution to the system of constraints is solved as described in [135],[18] repeated at each timestep since $\nu$ varies with time.

##### A.3.13 Partnership Numbers

This section summarizes the numbers of partnerships modelled among activity groups. Similar to group sizes, we draw on the analysis of FSW data in § A.3.9 and bias adjustment for wider population in § A.3.10, we well as further adjustments with regards to partnership duration [137].

**Adjusting for Partnership Duration** As noted in § A.2, sexual partnerships are usually quantified using a change rate Q, whereas our force of infection equation uses a number of current partners $K$. Either parameter may be estimated from survey questions like "*How many casual sexual partners have you had in the past 12 months?*" However, this estimation must account for both the recall period $\omega$ (*e.g.*, 12 months) and partnership duration $\delta$ (*e.g.*, 6 months) per [137]:

$$Q = \frac{x}{\omega + \delta} \tag{A.19}$$

$$K = \frac{x\delta}{\omega + \delta} = Q\delta \tag{A.20}$$

where $x$ is the mean number of partners reported in the recall period.

###### A.3.13.1 Sex Work Partnerships

**Female Sex Workers** Table A.4 summarizes the numbers of new and repeat clients *per month* reported by Swati FSW, stratified by higher *vs* lower risk per the analysis in § A.3.9. These data thus would provide $x$ for $\omega = 1$ month. However, based on the survey questions,[19] it's not clear whether these reported partner numbers represent the numbers of unique men or unique client visits.

We assumed that all *new* clients were one-off visits; thus the reported partner numbers effectively represented 1/12th of the total numbers of yearly partnerships $Q_{\text{SWO}}$. As such, we sampled the yearly rate of one-off sex work partnerships among lower risk FSW from a gamma distribution with mean (95% CI) as 3.5 (2.3, 5.0) × 12, and the rate among higher risk FSW from 14 (9, 20) × 12. Since each partnership is assumed to include only

[17] Subject to $f \le (\nu - \mu + D_{si}^{-1})\,\nu^{-1}$, which can be derived from Eq. (10) in [135].
[18] Using `docs.scipy.org/doc/scipy/reference/generated/scipy.optimize.nnls.html`
[19] The survey questions were: "*In the last 30 days, how many (new/regular) clients have you had sex with?*", or similar.

one sex act, the partnership duration $\delta_{SWO}$, frequency of sex $F_{SWO}$, and number of concurrent partnerships $K_{SWO}$ are ill-defined, but can be defined for convenience as $\delta_{SWO} = 1/12$ (years), $F_{SWO} = 12$ (per year), and $K_{SWO} = Q_{SWO}/12$.

For *repeat* sex work partnerships, uncertainties remain regarding partnership duration $\delta_{SWR}$ (see § A.3.15), frequency of sex per month $F_{SWR}/12$, and survey responses $x$ reflecting unique clients or total client visits per month. If $x$ reflects the numbers of unique clients, then $Q_{SWO}$ can be defined via Eq. (A.19) using $x$ directly; whereas if $x$ reflects the numbers of unique visits, then $Q_{SWO}$ should be defined using $x/(F_{SWO}/12)$. We assumed that 2/3 *vs* 1/3 of respondents interpreted the question as in the former *vs* latter case, such that:

$$x' = (2/3)\, x + (1/3)\, x/(F_{SWR}/12) \tag{A.21}$$

Taking $F_{SWR}/12 = 2$ as the prior mean from § A.3.14, Eq. (A.21) simplifies to $x' = \frac{5}{6}\, x$. Thus, we defined $K_{SWR}$ via Eqs. (A.20) and (A.21), with: $\omega = 1/12$ (1 month), $\delta_{SWR}$ as specified in § A.3.15, and $x_{SWR}$ from the gamma distributions given in § A.3.9.

**Clients** Across Sub-Saharan Africa, data for clients of FSW on the number of unique FSW visited and the frequency of sex is sparse. Among 64 clients in Kenya, the median number of sex work visits per week was 1.3 (68 per year); most clients (68%) had 1–3 regular FSW partners simultaneously, and visited 0–3 new FSW per year [140]. Among 261 truck drivers at sex work hotspots in Uganda, the mean number of sexual partners was 7.4 in the past 30 days and 44.7 in the past year [145]. Johnson and Dorrington [146] modelled yearly sex work visits among South African clients of FSW as gamma-distributed with age over 10, peaking at 64 visits per year for clients aged 37. To reflect these data, we specified clients overall to have mean (95% CI) 36 (18, 72) sex acts with FSW per year ($K_{SW}\, F_{SW}/12$, gamma prior). Then, the yearly sex acts among lower and higher risk clients are defined such that higher risk have 2.0 (1.6, 2.5) times the number among lower risk. Finally, since the distribution of sex acts between new *vs* regular sex work partnerships must match that among FSW, the specific values of $K_{SW}$ were computed automatically.

###### A.3.13.2 Main/Spousal & Casual Partnerships

Drawing on the results in § A.3.10.4, we defined the numbers of main/spousal and casual partners among each activity group as follows.

**Main/Spousal Partnerships** To simplify model fitting, we sampled a common proportion $x$ of individuals reporting a main/spousal partnership from a BAB distribution with 95% CI (25, 50)%, applied to all women and men in the lowest activity groups, as well as all women in the medium activity group. Then, we used Eq. (A.20) to define $K$ using $\omega = 1$ year and the main/spousal partnership duration from § A.3.15. Since FSW and clients had fewer main/spousal partnerships (see below), we calculated the proportion of men in the medium activity group having main/spousal partnerships to balance the total number of main/spousal partnerships among women and men.

**Casual Partnerships** We similarly defined a common proportion of women and men in the lowest activity groups reporting casual partnership $x_{CAS}$ with 95% CI (20, 55)%. However, the number of casual partnerships among $W_{2+}$ and $M_{2+}$ ramains uncertain. The analysis in § A.3.10 provides no information on these values, but the number of casual partners in p12m for the medium activity groups must be at least about 1.5 to ensure these women and men actually have 2+ partners in p12m. Thus, we sampled the number of casual partners reported by women in the medium activity group $x$ from a gamma distribution with 95% CI (1.2, 2), and computed $K$ via Eq. (A.20). As before, we calculated the numbers of casual partnerships among men in the medium activity group to balance total casual partnerships.

**Main/Spousal & Casual Partnerships among FSW & Clients** Among Swati FSW, the mean number of total non-paying partners in the past month was approximately 1–1.5 (Table A.4), which could include both main/spousal partners and casual partners. Among FSW in South Africa [147] and Kenya [148], while 54 and 72% (respectively) reported being in a relationship, only 6 and 3% were married, although many non-marital partners may still constitute effectively "main" partnerships with respect to condom use and duration. Thus, we assumed that: 50% of all FSW reported a main/spousal partner in p12m; lower risk FSW reported 0.5 casual partners; and higher risk FSW reported 1.0 casual partners, on average.

Available data suggest that about half of clients also report non-sex work partners, which are not always distinguished as main/spousal *vs* casual partnerships [64,139]. Non-paying partners of FSW are also often clients of other FSW [148,149]. Yet, clients of FSW also tend to be younger and more likely to be never/formerly married *vs* non-client men [139,150]. So, we assumed that clients reported half the numbers of main/spousal partnerships compared to lowest activity men, and 25–100% the numbers of casual partnerships compared to medium activity women (uniform prior). As before, we computed $K$ via Eq. (A.20) with partnership durations from § A.3.15.

##### A.3.14 Sex Frequency

The Eswatini general population data sources [2,5,20] did not report on frequency of sex. In South Africa, average numbers of sex acts per week per partnership (non-sex work) was reported as mean 2.5 (IQR: 1–3) [151], with consistent reports across main/spousal partnerships and casual partnerships. Sex frequency among South Africans per month overall (not per-partnership) is also summarized in [152, Figure 3.15], which is roughly consistent with [151], but motivates a smaller lower bound. Median sex frequency per partnership-year in 1998 Rakai, Uganda was approximately 90 acts with the "more frequent" of concurrent partners, and approximately 20 acts with the "less frequent" [153]. Considering these data, we sampled the number of sex acts per year in main/spousal partnerships from a gamma prior distribution with 95% CI (26, 156), and a relative rate for casual partnerships from Unif (0.5, 2). As described in § A.3.13.1, we defined $F_{\text{SWO}} = 12$ for one-off sex work partnerships, and $F_{\text{SWR}} \sim \text{Unif}(12, 36)$ for repeat sex work partnerships. We also constrained samples of $F_{p_4}$ such that higher risk FSW never have commercial sex more than twice daily, on average.

**Anal Sex** Among Eswatini data sources, only [19] (FSW, 2014) counted sex acts separately for anal and vaginal sex.[20] Among all FSW, the proportion of "average sex acts per week" that were anal (vs vaginal) was 2.9%. However, a previous coital diary study in neighbouring KwaZulu-Natal suggested much higher proportions

[20] Owen et al. [154] examined prevalence of anal sex in p1m among Swati FSW in 2011, but could not comment on frequency due the survey questions.

were anal [155], and face-to-face interview survey design may result in under-reporting [59]. Owen et al. review studies of anal sex in South Africa, and estimate that 0.6–16.5% of sex acts among the general population are anal [156], *vs* 2.4–15.9% among FSW [59]. To reflect this data, we sampled the proportions of sex acts which are anal in main/spousal and casual partnerships from a gamma prior distribution with 95% CI (0.6, 16.5)%, and a relative proportion in all sex work partnerships from Unif (1, 2).

##### A.3.15 Partnership Duration

Eswatini-specific data on partnership duration are lacking. Moreover, accurate estimation of partnership duration remains challenging even when data exist, due to censoring, truncation, and sampling biases [137,157]. Similar to challenges in estimating sex work duration, we must distinguish the definition of an "average partnership" as (a) among all partnerships in a population over a given *time period*, *vs* (b) among all partnerships in a population *cross-section*. Case (b) will be biased by partnership duration, so the estimated mean duration will longer, while case (a) reflects an unbiased estimate.[21] The difference between the exponential distribution mean ($1/\lambda$) and median ($\log 2/\lambda$) should also be kept in mind.

**Main/Spousal Partnerships** Detailed data on marriage in Eswatini was only captured in 2006 [2, Table 6.1]. The median age of first marriage was 24.3 among women and 27.7 among men (26.0 overall). Approximately 64% of women and 88% of men (76% overall) who were ever married or living together were in a union at age 50–54. However, no data indicated whether any respondents had remarried or entered into a secondary union. Among women aged 40–49, the most recent data on median age of first marriage and proportions ever remarried were 33 years old and 6.6% in South Africa, 20.9 and 3.7% in Lesotho, and 18.7 and 28.4% in Mozambique [158]; such data may not capture non-marital secondary unions. Thus, we assumed $\rho$ = 5–20% of unions among EmaSwati aged 50–54 were secondary. Considering that the modelled population only includes ages 15–49, we then defined the mean durations of main/spousal partnerships as $\delta_{\text{MSP}} = (0.76 - \rho)\,(49 - 26) \in (14.5, 18.5)$ years.

In some models, partnership duration is used to define both the total numbers of sex acts per partnership and the partnership change rate (see § A.2). This change rate might be overestimated by the above definition, since the rate should also consider whether and when divorced/separated individuals form *new* main/spousal partnerships. The change rate could even be tied to the modelled baseline and HIV-attributable mortality, given that the majority of Swati unions ended via spousal death (83% of unions among women and 56% among men by age 50–54) [2]. For simplicity and consistency with prior approaches, we used the effective duration of 14.5–18.5 years throughout (uniform prior).

**Casual Partnerships** No data is available regarding durations of non-marital sexual partnerships in Eswatini, and regional data on are also limited. We synthesized the available partnership duration data from South Africa [159–161], Rural Tanzania [121], and four cities in Kenya, Zambia, Benin, and Cameroon [162]. Based on these data, we defined a gamma prior distribution for the mean duration of casual partnerships $\delta_{\text{CAS}}$ with

[21] If case (a) durations are exponentially distributed, the durations in case (b) will be gamma-distributed with $\alpha = 2$, $\beta = \lambda$; thus the mean duration in case (b) will be $\alpha/\beta = 2\lambda$ (twice as long).

95% CI (0.25, 1.5) years, roughly consistent with prior models [163]. A gamma distribution was chosen *vs* uniform or normal to reflect non-uniform belief while preventing negative values.

**Sex Work Partnerships** As noted in § A.3.13.1, duration of one-off sex work partnerships is ill defined, but can be defined to comprise a single sex act with $F_{\text{SWO}}\delta_{\text{SWO}} = 1$. Data on repeat sex work partnerships is severely limited, and sometimes regular paying clients later become non-paying emotional partners [148,164]. Based on [140], I defined a gamma prior distribution for the mean duration of repeat sex work partnerships $\delta_{\text{SWR}}$ with 95% CI (2, 12) months.

##### A.3.16 Mixing

In addition to more transmission among FSW and their clients via one-off and repeat sex work partnerships — which are *only* formed among FSW and clients — other types of partnerships may be formed preferentially between particular activity groups. For example, FSW and clients may be more likely to form main partnerships with each other than with other activity groups. Such preferences are captured in a "mixing matrix" $M$, where $M_{pii'}$ denotes the total number of type-$p$ partnerships formed between groups $i$ and $i'$ in the population (ignoring sex indices $s, s'$ temporarily) — *i.e.*, who has sex with whom. The mixing matrix $M_{pii'}$ must be symmetric, and have row/column sums equal to the total numbers of partnerships "offered" by any group: $M_{pi} = P_i K_{pi}$ (group size × partnerships per-person).

###### A.3.16.1 Log-Linear Mixing

Many risk/activity-stratified compartmental transmission models parameterize mixing via a single parameter $\epsilon \in [0, 1]$, which controls the degree of like-with-like mixing [165,166]. However, the simplicity of this approach precludes more complex mixing patterns — such as preferential mixing among two of four total groups. A more general approach to mixing is developed in [167]. This "log-linear" approach defines the mixing matrix elements $M_{pii'}$ as follows. The expected total numbers of partnerships between risk groups under random mixing are defined as:

$$\Pi_{pii'} = \frac{M_{pi} M_{pi'}}{\sum_j M_{pj}} \tag{A.22}$$

Next, a matrix $\Phi_{pii'}$ is defined, representing the odds of a type-$p$ partnership forming between groups $i$ and $i'$, compared to random mixing. The matrix $\Phi$ must be symmetric, and can be estimated directly from the right kind of data (which is rarely available) [167]. Then, an initial estimate of $M_{pii'}$ is:

$$\begin{aligned} M^{(0)}_{pii'} &= \exp\left[\log\left(\Pi_{pii'}\right) + \Phi_{pii'}\right] \\ &= \Pi_{pii'} \exp\left(\Phi_{pii'}\right) \end{aligned} \tag{A.23}$$

However, this estimate changes the total numbers of partnerships formed by each group: $M^{(0)}_{pi} \neq \Pi_{pi}$, where $M_{pi} = \sum_{i'} M_{pii'}$ and $\Pi_{pi} = \sum_{i'} \Pi_{pii'}$. There is no *a priori* definition of $M_{pii'}$ or adjustment to $\Phi_{pii'}$ that can guarantee the numbers of partnerships will not change. However, an iterative proportional fitting procedure

[168] can resolve an estimate $M_{pii'}^{(\infty)}$ that maintains the total numbers of partnerships:

$$M_{pii'}^{(n+1)} = M_{pii'}^{(n)} \frac{\Pi_{pf}}{M_{pf}^{(n)}} \qquad f = \begin{cases} i & \text{if } n \text{ is even} \\ i' & \text{if } n \text{ is odd} \end{cases} \tag{A.24}$$

Each step of this procedure can be understood as a re-scaling of the current estimate $M_{pii'}^{(n)}$ row-wise ($i$) or column-wise ($i'$) to match the numbers of partnerships offered by individuals ($\Pi_{pi}$) or their partners ($\Pi_{pi'}$). Each row-step re-introduces discrepancies in the columns, and vice versa, but overall convergence is guaranteed [169].

In practice, Eq. (A.24) adds approximately one decimal of precision per $2n$ for the $4 \times 4$ case, thus 15–20 iterations is often sufficient to come within computational precision limits. Since the partnerships matrix $M_{pii'}$ should adapt to reflect changes in group sizes (*e.g.*, due to HIV mortality) or numbers of partnerships offered (*e.g.*, see § A.2), the matrix must be re-computed at every time point. Thus, the procedure Eq. (A.24) could be considered computationally expensive. However, this approach provides great flexibility and interpretability to specify complex mixing patterns via the odds matrix $\Phi_{pii'}$.

Two final adjustments are needed for the bipartite (*i.e.*, heterosexual) system, after adding back the sex dimension indices $i \longrightarrow si$, $i' \longrightarrow s'i'$. First, we ensure that $M_{s=s'} = \Pi_{s=s'} = 0$. Second, for the case when the total numbers of partnerships offered by women and men do not balance ($\sum_j M_{ps_1j} \neq \sum_j M_{ps_2j}$), We revise the denominator of Eq. (A.22) to $\sum_j \psi_s M_{psj}$, where $\psi_s$ are weights such that $\sum_s \psi_s = 1$. Similar to the "compromise" parameter $\theta$ in [166], if $\psi = \{1, 0\}$, then women's partnership numbers are matched exactly while men adapt their partner numbers to balance; and conversely for $\psi = \{0, 1\}$. We fixed $\psi = \{0.5, 0.5\}$ for equal adaptation among women and men.

###### A.3.16.2 Odds of Mixing

Despite the flexibility in the odds of mixing matrix $\Phi_{pii'}$, and the importance of mixing patterns for transmission dynamics [170], there are limited data to inform mixing patterns for Eswatini. In Kenya [148], Benin, Guinea, and Senegal [149], and Uganda [164], a disproportionate fraction of non-paying partners of FSW were former and/or current clients. However, its not clear whether such partnerships reflect main/spousal and/or casual partnerships. As such, we sampled a common value for both partnership types $\sim \exp[\text{Unif}\,(-2, +2)]$, applied equally to higher and lower risk FSW and clients. We further assumed that lowest activity women and men had greater odds of forming main/spousal partnerships with each other, based loosely on age cohorting effects [171], observed like-with-like sexual mixing preferences in other contexts [167,172,173], and prior models [108]. We sampled this odds ratio from an equal prior: $\exp[\text{Unif}\,(-2, +2)]$. We made no further assumptions about preferential mixing (*i.e.*, all other elements $\Phi = 0$). Thus, we assumed that one-off and repeat sex work partnerships form randomly with respect to higher *vs* lower FSW and their clients.

Table A.5: Definitions and distributions of calibrated parameters

| | | Prior | | | Posterior | |
|---|---|---|---|---|---|---|
| Parameter | Definition | Type | Mean | (95% CI) | Mean | (95% CI) |
| t0_hiv | year of HIV introduction to Eswatini | Uniform | 1982.5 | (1980.1, 1984.9) | 1982.6 | (1980.3, 1985.0) |
| PX_w_fsw | proportion of women who are FSW | Beta | 0.0288 | (0.00703, 0.065) | 0.0344 | (0.0207, 0.0526) |
| PX_w_h | proportion of women who have 2+ partners in p12m | Beta | 0.178 | (0.0961, 0.278) | 0.191 | (0.134, 0.245) |
| PX_m_m | proportion of men who have 2+ partners in p12m | Beta | 0.133 | (0.1, 0.17) | 0.137 | (0.103, 0.164) |
| dur_fsw | duration in sex work overall | Gamma | 4.07 | (2.29, 6.34) | 3.81 | (2.56, 5.01) |
| dur_sw_h | duration in higher risk sex work | Gamma | 0.5 | (0.17, 1.0) | 0.583 | (0.285, 0.936) |
| dur_cli | duration buying sex among clients | Gamma | 8.63 | (4.0, 15.0) | 8.79 | (4.89, 12.7) |
| turn_xm_xl | turnover rate from medium to lowest activity (women and men) | Gamma | 0.216 | (0.05, 0.5) | 0.231 | (0.11, 0.443) |
| Pturn_fsw_m:l | proportion of FSW who transition to medium activity | Beta | 0.724 | (0.503, 0.898) | 0.743 | (0.594, 0.881) |
| Pturn_cli_m:l | proportion of clients who transition to medium activity | Beta | 0.602 | (0.249, 0.9) | 0.6 | (0.361, 0.804) |
| growth_2050 | rate of Eswatini population growth in 2050 | Uniform | 0.011 | (0.0072, 0.0148) | 0.0108 | (0.00737, 0.0144) |
| C12m_msp_xl | number of main/spousal partners in p12m among lowest activity | Beta | 0.37 | (0.251, 0.498) | 0.371 | (0.277, 0.469) |
| C12m_cas_xl | number of casual partners in p12m among lowest activity | Beta | 0.366 | (0.201, 0.549) | 0.368 | (0.239, 0.497) |
| C12m_cas_wm | number of casual partners in p12m among medium activity women | Gamma | 1.58 | (1.2, 2.0) | 1.67 | (1.43, 1.95) |
| RC_cas_cli:wm | relative casual partners among clients vs medium activity women | Uniform | 0.625 | (0.269, 0.981) | 0.751 | (0.296, 1.0) |
| C1m_swo_fsw_l | number of one-off sex work partners in p1m among lower risk FSW | Gamma | 3.5 | (2.27, 5.0) | 3.28 | (2.45, 4.34) |
| C1m_swr_fsw_l | number of repeat sex work partners in p1m among lower risk FSW | Gamma | 6.0 | (3.88, 8.57) | 5.42 | (3.38, 7.69) |
| C1m_swo_fsw_h | number of one-off sex work partners in p1m among higher risk FSW | Gamma | 14.0 | (9.06, 20.0) | 13.7 | (9.67, 17.3) |
| C1m_swr_fsw_h | number of repeat sex work partners in p1m among higher risk FSW | Gamma | 21.0 | (13.6, 30.0) | 22.7 | (14.8, 29.2) |
| KF_swx_cli | rate of visiting FSW (sex acts) among clients overall | Gamma | 40.5 | (18.0, 71.9) | 51.4 | (32.6, 71.4) |
| RKF_swx_cli_h:l | relative visits (sex acts) among higher vs lower risk clients | Gamma | 2.03 | (1.6, 2.5) | 2.04 | (1.74, 2.38) |
| F_msp | rate of sex acts in main/spousal partnerships | Gamma | 77.3 | (26.0, 156.0) | 86.0 | (56.3, 123.0) |
| RF_cas:msp | relative rate of sex acts in casual vs main/spousal partnerships | Uniform | 1.25 | (0.538, 1.96) | 1.6 | (1.0, 2.0) |
| dur_msp | duration of main/spousal partnerships | Uniform | 16.5 | (14.6, 18.4) | 16.6 | (15.0, 18.5) |
| dur_cas | duration of casual partnerships | Gamma | 0.743 | (0.25, 1.5) | 0.78 | (0.484, 1.12) |
| dur_swr | duration of repeat sex work partnerships | Gamma | 0.495 | (0.166, 1.0) | 0.435 | (0.212, 0.78) |
| F_swr | rate of sex acts in repeat sex work partnerships | Uniform | 24.0 | (12.6, 35.4) | 20.7 | (12.0, 32.3) |
| PF_ai_mcx | proportion of anal sex acts in main/spousal and casual partnerships | Gamma | 0.0573 | (0.00603, 0.165) | 0.114 | (0.0584, 0.217) |
| RPF_ai_swx:mcx | relative proportion of anal sex acts in sex work vs other partnerships | Uniform | 1.5 | (1.02, 1.98) | 1.57 | (1.05, 1.98) |
| lpref_msp_xl | log-odds of main/spousal partnership formation among lowest activity | Uniform | 0 | (-1.9, 1.9) | -0.217 | (-1.93, 1.21) |
| lpref_mcx_swx | log-odds of non-sex work partnership formation among FSW and clients | Uniform | 0 | (-1.9, 1.9) | -0.0689 | (-1.79, 1.57) |

continued ...

continued …

| | | | | | | |
|---|---|---|---|---|---|---|
| `Rbeta_condom` | relative per-act probability of HIV transmission with a condom | Beta | 0.266 | (0.131, 0.429) | 0.304 | (0.194, 0.409) |
| `RPF_condom_a:v` | relative condom use in anal vs vaginal sex | Beta | 0.768 | (0.504, 0.949) | 0.733 | (0.553, 0.903) |
| `RPF_condom_1996` | elative condom use in all partnerships in 1996 vs 2002 or 2006 | Uniform | 0.5 | (0.025, 0.975) | 0.53 | (0.054, 0.998) |
| `PF_condom_msp_2006` | condom use in main/spousal partnerships in 2006 | Beta | 0.23 | (0.153, 0.317) | 0.23 | (0.172, 0.296) |
| `PF_condom_msp_2016` | condom use in main/spousal partnerships in 2006 | Beta | 0.416 | (0.308, 0.529) | 0.418 | (0.335, 0.504) |
| `PF_condom_cas_2006` | condom use in casual partnerships in 2006 | Beta | 0.598 | (0.501, 0.692) | 0.594 | (0.53, 0.671) |
| `PF_condom_cas_2016` | condom use in casual partnerships in 2016 | Beta | 0.694 | (0.601, 0.78) | 0.695 | (0.636, 0.767) |
| `PF_condom_swo_2002` | condom use in one-off sex work partnerships in 2002 | Beta | 0.432 | (0.148, 0.744) | 0.486 | (0.254, 0.762) |
| `PF_condom_swo_2011` | condom use in one-off sex work partnerships in 2011 | Beta | 0.777 | (0.581, 0.923) | 0.797 | (0.708, 0.879) |
| `PF_condom_swo_2014` | condom use in one-off sex work partnerships in 2014 | Beta | 0.787 | (0.547, 0.95) | 0.862 | (0.779, 0.939) |
| `PF_condom_swr_2002` | condom use in repeat sex work partnerships in 2002 | Beta | 0.337 | (0.118, 0.603) | 0.294 | (0.126, 0.485) |
| `PF_condom_swr_2011` | condom use in repeat sex work partnerships in 2011 | Beta | 0.754 | (0.568, 0.9) | 0.706 | (0.608, 0.788) |
| `PF_condom_swr_2014` | condom use in repeat sex work partnerships in 2014 | Beta | 0.759 | (0.481, 0.949) | 0.745 | (0.649, 0.84) |
| `PF_circum_2050` | prevalence of circumcision by 2050 | Beta | 0.724 | (0.503, 0.898) | 0.741 | (0.581, 0.871) |
| `beta_0` | per-act probability of HIV transmission $\beta$ for CD4 > 350 (REF) | Gamma | 0.00131 | (0.000498, 0.00251) | 0.00174 | (0.00123, 0.00247) |
| `Rbeta_acute` | relative $\beta$ during acute infection | Gamma | 6.01 | (1.11, 15.0) | 9.33 | (4.98, 15.8) |
| `Rbeta_350` | relative $\beta$ for 200 < CD4 < 350 | Gamma | 1.59 | (1.3, 1.9) | 1.55 | (1.32, 1.76) |
| `Rbeta_200` | relative $\beta$ for CD4 < 200 | Gamma | 8.2 | (4.5, 13.0) | 8.17 | (5.48, 11.1) |
| `Rbeta_vi_rec` | relative $\beta$ for receptive vaginal sex | Uniform | 1.5 | (1.02, 1.98) | 1.66 | (1.21, 1.99) |
| `aRbeta_gud_sus` | additional relative $\beta$ for GUD among susceptible partner | Gamma | 2.05 | (0.2, 6.0) | 2.55 | (0.415, 5.45) |
| `aRbeta_gud_inf` | additional relative $\beta$ for GUD among infectious partner | Gamma | 0.99 | (0.2, 2.4) | 1.04 | (0.272, 1.99) |
| `dur_acute` | duration of acute infection | Gamma | 0.172 | (0.0174, 0.5) | 0.288 | (0.111, 0.489) |
| `P_gud_fsw_l` | prevalence of GUD among lower risk FSW | Beta | 0.295 | (0.2, 0.4) | 0.294 | (0.218, 0.368) |
| `RP_gud_fsw_h:l` | relative prevalence of GUD among higher vs lower risk FSW | Gamma | 1.28 | (1.0, 1.6) | 1.31 | (1.07, 1.52) |
| `RP_gud_2030` | relative prevalence of GUD overall in 2030 vs 2010 | Uniform | 0.6 | (0.22, 0.98) | 0.817 | (0.366, 1.0) |
| `iP_gud_h:l` | interpolator for GUD among medium activity vs DHS and FSW | Uniform | 0.5 | (0.025, 0.975) | 0.518 | (0.0537, 0.953) |
| `Rbeta_uvls` | relative $\beta$ on ART but before VLS | Beta | 0.244 | (0.0139, 0.656) | 0.302 | (0.0656, 0.614) |
| `Rdx_global` | relative rate of diagnosis overall | Uniform | 0.75 | (0.512, 0.988) | 0.645 | (0.536, 0.775) |
| `dx_w_2002` | rate of diagnosis among women in 2002 | Beta | 0.094 | (0.0452, 0.158) | 0.0949 | (0.056, 0.135) |
| `dx_w_2006` | rate of diagnosis among women in 2006 | Beta | 0.248 | (0.169, 0.337) | 0.271 | (0.218, 0.319) |
| `Rdx_m:w_2006` | relative rate of diagnosis among men vs women in 2006 | Gamma | 0.377 | (0.207, 0.597) | 0.471 | (0.333, 0.66) |
| `dx_wq_2011` | rate of diagnosis among non-FSW women in 2011 | Gamma | 0.637 | (0.466, 0.834) | 0.544 | (0.421, 0.657) |
| `Rdx_m:wq_2011` | relative rate of diagnosis among men vs non-FSW women in 2011 | Gamma | 0.529 | (0.351, 0.742) | 0.467 | (0.385, 0.555) |
| `aRdx_fsw:wq_2011` | additional relative diagnosis among FSW vs non-FSW women in 2011 | Gamma | 0.521 | (0.206, 0.98) | 0.542 | (0.301, 0.854) |
| `aRdx_wq_16:11` | additional relative diagnosis among non-FSW women in 2016 vs 2011 | Gamma | 0.204 | (0.118, 0.313) | 0.186 | (0.12, 0.264) |

continued …

continued …

| | | | | | | |
|---|---|---|---|---|---|---|
| aRdx_fsw:wq_2016 | additional relative diagnosis among FSW vs non-FSW women in 2016 | Gamma | 0.619 | (0.291, 1.07) | 0.589 | (0.416, 0.848) |
| tx_2010 | rate of ART initiation among diagnosed and eligible in 2010 | Gamma | 1.5 | (0.509, 3.02) | 1.5 | (0.937, 2.1) |
| tx_2012 | rate of ART initiation among diagnosed and eligible in 2012 | Gamma | 8.75 | (6.01, 12.0) | 7.75 | (5.91, 9.8) |
| Rtx_fsw:wq | relative rate of ART initiation among FSW vs non-FSW women | Uniform | 0.75 | (0.512, 0.988) | 0.757 | (0.522, 0.966) |
| ivx | duration on ART before achieving VLS initially | Gamma | 0.62 | (0.33, 1.0) | 0.75 | (0.589, 0.887) |
| Runvx_m:wq | relative rate of viral unsuppression among men vs non-FSW women | Uniform | 1.25 | (1.01, 1.49) | 1.3 | (1.0, 1.5) |
| Runvx_fsw:wq | relative rate of viral unsuppression among FSW vs non-FSW women | Uniform | 1.25 | (1.01, 1.49) | 1.24 | (1.01, 1.47) |
| revx_2010 | rate of viral re-suppression in 2010 | Gamma | 0.729 | (0.5, 1.0) | 0.609 | (0.492, 0.788) |

FSW: female sex worker; p12m: past 12 months; $\beta$: per-act probability of HIV transmission; GUD: any genital ulcer disease in p12m; ART: antiretroviral therapy; VLS: viral load suppression; additional relative (aR): relative value beyond one, *e.g.*, $R = 1.5 \longrightarrow aR = 0.5$; prevalence interpolator (iP): *e.g.*, $P_0 = 0.2$, $P_1 = 0.4$, $iP_x = 0.5 \longrightarrow P_x = 0.3$; all rates in per-year; all durations in years; all parameters reflect stratum averages.

#### A.4 Model Calibration

We considered uncertainty in 74 model input parameters (Table A.5). For each uncertain parameter, we specified a univariate prior distribution based on the available data and/or assumptions (§ A.3). Model calibration aims to reduce this uncertainty — *i.e.*, estimate the (joint) parameter posterior distribution — by comparing model outputs to "calibration targets" under different combinations of input parameters. Table A.5 summarizes the calibrated parameters, including the mean (95% CI) for prior and posterior distributions; § A.4.1 describes our approach to calibration, and § A.4.2 details the calibration targets used. These targets include estimates of HIV incidence, prevalence, and the cascade of care for the population overall, and stratified by risk group where possible. Results of model calibration are given in § B.1.

##### A.4.1 Approach

We used the Incremental Mixture Importance Sampling (IMIS) procedure [174] for model calibration, with added sampling constraints and adjusted weights as described below. Let $\theta$ denote the complete set of 74 calibrated model parameters (Table A.5), and $T$ the complete set of 78 calibration targets (§ A.4.2).

**Prior Sampling & Constraints** In order to obtain good coverage of the sampling space, most (59) calibrated parameters were initially sampled using Latin hypercube sampling [175]. The remaining 15 calibrated parameters were sampled randomly and iteratively until they satisfied a set of constraints:

a. from § A.3.14, resample `F_swr` only:
```
K_swo_fsw_h * F_swo + K_swr_fsw_h * F_swr < 2*365
```
where: `K_swx_fsw_h = C1m_swx_fsw_h * dur_swx / (dur_swx + 1/12)`

b. from § A.3.5.2, let "`c_`" denote `PF_condom_`:
```
c_msp_2006 < c_msp_2016
c_cas_2006 < c_cas_2016
c_swo_2002 < c_swo_2011 < c_swo_2014
c_swr_2002 < c_swr_2011 < c_swr_2014
c_msp_2006 < c_cas_2006
c_msp_2016 < c_cas_2016
c_swr_2002 < c_swo_2002
c_swr_2011 < c_swo_2011
c_swr_2014 < c_swo_2014
```

c. from § A.3.4.1:
```
1 <= (Rbeta_acute * dur_acute) <= 63
```

d. from § A.3.5.3:
```
P_gud_fsw_l > .07
(P_gud_fsw_l * RP_gud_fsw_h:l) < 1
```

As shown in Figure A.9, this approach reduces distortion of sampled *vs* prior distributions, as compared to forward or backward conditional sampling. When sampling from the multivariate Gaussian distributions during each IMIS step [174], we attempted up to 1000 times per sample to find a parameter set $\theta$ that satisfied all constraints; if no such parameter set could be identified, we set the weight of this $\theta$ to zero and continued.

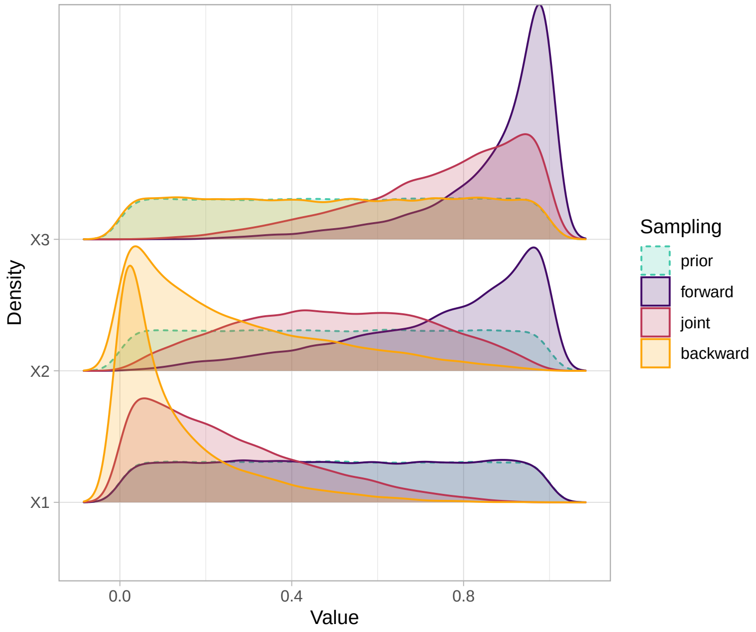

Figure A.9: Illustration of different sampling biases when enforcing $X_1 < X_2 < X_3$

Sampling method: *joint:* sample $X_1, X_2, X_3$ simultaneously; then discard any samples failing $X_1 < X_2 < X_3$; *forward:* sample $X_1$; then sample $X_2$ until $X_1 < X_2$; then sample $X_3$ until $X_2 < X_3$; *backward:* sample $X_3$; then sample $X_2$ until $X_2 < X_3$; then sample $X_1$ until $X_1 < X_2$.

**Likelihoods** We defined the log likelihood $L_i$ of a given parameter set $\theta_i$ as the sum of independent log likelihoods for each calibration target $T_j$:

$$L_i = \sum_j f_j \log p(T_j \mid \theta_i) \tag{A.25}$$

where $f_j$ is a scale factor (usually 1) applied to target $T_j$ to increase or decrease its influence. Any log likelihood which was beyond computational precision was replaced with an arbitrarily large negative number: $-10^6$.

**Weights** Due to the high number of calibration targets and thus high variance in log likelihoods, IMIS weights defined per [174] exactly were usually degenerate, having all-but-one near-zero values. As such, our weight definitions used the following transformation of log likelihoods instead of actual likelihoods — *i.e.*, $\exp(L)$:

$$\tilde{L}_i = \frac{Q_L^{0.9}}{L_i} \tag{A.26}$$

where $Q_L^{0.9}$ is the 90% quantile of log likelihoods $L$. Figure A.10 illustrates the shape of this transform *vs* actual likelihoods, for dummy log likelihoods uniformly distributed in log space $\in [-10^6, -10^1]$. With this transformation: the 90% quantile becomes 1, a 10-fold higher $L$ becomes 10, and a 10-fold lower $L$ becomes 0.1. These transformed log likelihoods $\tilde{L}$ were used instead of actual likelihoods within the weight definitions for all stages: after initial sampling, within each IMIS step, and for the final resampling.

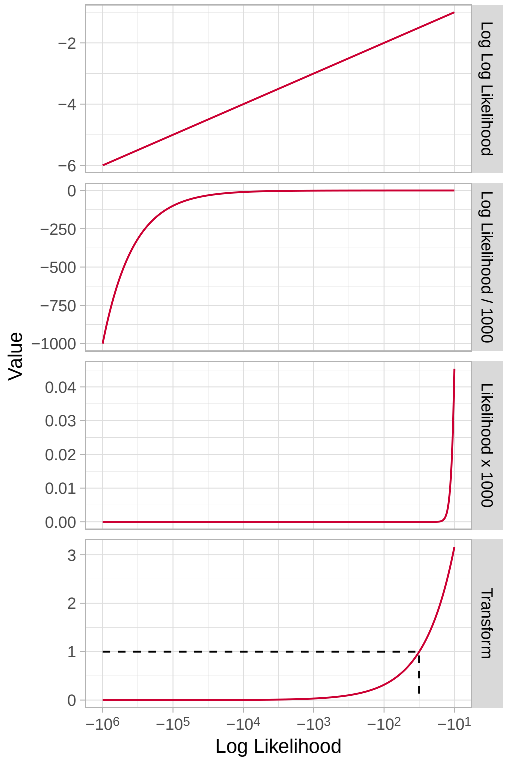

Figure A.10: Shape of log likelihood transform used for IMIS weight definitions

Dashed line indicates 90% quantile of log likelihoods, corresponding to a value of 1 after transformation

**Iterations** We ran 100 independent batches of the basic (without optimization) IMIS, each with 1000 initial samples, 100 resamples per IMIS step, and 100 total IMIS steps (1,100,000 total model runs), from which we resampled 1000 posterior parameter sets ("model fits") without replacement. In the notation of [174], we used 100 batches of: $N_0 = 1000$, $B = 100$, $J = 1000$. We used batches to allow model fitting in parallel. We did not use any stopping criterion, but verified visually that log likelihoods plateaued within each batch.

#### A.4.2 Calibration Targets

The data sources for Eswatini calibration targets are mainly the same as for Eswatini-specific parameters (see § A.3.2). We assumed that population-level surveys in 2006 (DHS) [2], 2011 (SHIMS1) [3,4], 2016 (SHIMS2) [5], and 2021 (SHIMS3) [21] reached FSW and their clients, although respondents may not report selling or buying sex in the context of these surveys.

Table A.6: Estimates of HIV prevalence in Eswatini

| Population [a] | Year | N | Raw % | Adj % | (95% CI) | Used | Ref | Notes |
|---|---|---|---|---|---|---|---|---|
| Overall | 2006 | 8,187 | 25.9 | — | (24.4, 27.3) | ✓ | [2] | b |
| | 2011 | 18,172 | 32.1 | 28.0 | (27.0, 29.0) | ✓ | [3] | cd |
| | 2016 | 8,533 | 27.2 | — | (25.8, 28.7) | ✓ | [5] | b |
| | 2021 | 12,043 | 23.7 | — | (22.6, 24.9) | ✓ | [21] | e |
| Women Overall | 2006 | 4,424 | 31.1 | — | (29.4, 32.9) | ✓ | [2] | b |
| | 2011 | 9,843 | 38.8 | 34.2 | (33.0, 35.4) | ✓ | [3] | cd |
| | 2016 | 4,878 | 34.3 | — | (32.6, 36.0) | ✓ | [5] | b |
| | 2021 | 6,985 | 31.6 | — | (29.8, 33.4) | ✓ | [21] | e |
| Men Overall | 2006 | 3,763 | 19.7 | — | (17.9, 21.4) | ✓ | [2] | b |
| | 2011 | 8,329 | 24.1 | 20.7 | (19.6, 21.8) | ✓ | [3] | cd |
| | 2016 | 3,655 | 18.8 | — | (17.3, 20.4) | ✓ | [5] | b |
| | 2021 | 5,058 | 15.6 | — | (14.3, 16.9) | ✓ | [21] | e |
| LR Overall | 2006 | 7,589 | 24.9 | — | — | ✗ | [2] | |
| | 2011 | 16,145 | 31.9 | — | — | ✗ | [3] | |
| | 2016 | 7,887 | 32.2 | — | — | ✗ | [5] | |
| Non-LR Overall | 2006 | 579 | 38.3 | — | — | ✗ | [2] | |
| | 2011 | 1,887 | 33.3 | 29.0 | (25.9, 32.2) | ✗ | [3] | cd |
| | 2016 | 914 | 28.7 | — | (25.8, 31.7) | ✗ | [5] | g |
| LR Women | 2006 | 4,346 | 30.7 | 26.8 | (22.7, 28.7) | * | [2] | f |
| | 2011 | 9,843 | 38.2 | 30.8 | (28.9, 32.8) | * | [3] | cf |
| | 2016 | 5,203 | 36.5 | 31.5 | (30.0, 33.1) | * | [5] | f |
| Non-LR Women | 2006 | 72 | 53.0 | — | (41.5, 64.3) | * | [2] | g |
| | 2011 | 373 | 54.5 | 48.1 | (41.5, 54.8) | * | [3] | cd |
| | 2016 | 263 | 45.3 | — | (39.3, 51.3) | * | [5] | g |
| LR Men | 2006 | 3,243 | 17.1 | 14.1 | (6.5, 16.7) | * | [2] | f |
| | 2011 | 6,733 | 23.2 | 19.0 | (18.0, 20.1) | * | [3] | cf |
| | 2016 | 2,684 | 25.1 | 16.9 | (15.7, 18.1) | * | [5] | f |
| Non-LR Men | 2006 | 506 | 36.1 | — | (32.0, 40.3) | * | [2] | g |
| | 2011 | 1,515 | 28.1 | 24.1 | (21.4, 26.9) | * | [3] | cd |
| | 2016 | 651 | 22.8 | — | (19.7, 26.1) | * | [5] | g |
| FSW Overall | 2011 | 328 | 70.3 | 60.5 | (52.1, 69.0) | ✓ | [18] | h |
| | 2014 | 781 | 37.8 | — | — | ✗ | [19] | i |
| | 2021 | 676 | 60.8 | 58.8 | (53.9, 63.6) | ✓ | [18] | h |

[a] LR: lower risk, reporting 0-1 partners p6m; Non-LR: lower risk, reporting 2+ partners p6m; FSW: female sex worker; [b] 95% CI as reported from sampling adjustment; [c] adjusted from ages 18–49 to 15–49 (see § A.4.2.1); [d] 95% CI expanded via inferred sampling adjustment; [e] N for survey overall; [f] adjusted for biased reporting of risk behaviours (see § A.3.10 and § A.4.2.1); [g] 95% CI inferred from N; [h] RDS-adjusted; [i] self-reported; * used within prevalence ratio only; all estimates used the BAB distribution.

###### A.4.2.1 HIV Prevalence

Table A.6 summarizes the available HIV prevalence data for Eswatini. Uncertainty around each estimate was modelled using a BAB distribution. We made several adjustments to these estimates as described below.

**Sampling Error** Population-level HIV prevalence estimates in 2006 and 2016 included expanded 95% CI (*vs* standard binomial 95% CI) due to sampling error for women, men, and the population overall (Table B.2 in [2] and Table C.2 in [5]). This expanded 95% CI corresponds to a reduction in effective $N$ *vs* the sample $N$ for the binomial distribution, by a factor of 41–75%. We applied this factor to equivalently expand the estimated 95% CI for the corresponding lower risk and non-lower risk women, men, and population overall in 2006 and 2016, and also for all 2011 HIV prevalence estimates [3].

**Biased Partner Number Reporting** As discussed in § A.3.10, we assumed that the proportion of the population reporting 0–1 sexual partners p6m ("lower risk") is overestimated, and the proportion reporting 2+ ("non-lower risk") is underestimated. While overall HIV prevalence estimates would not be affected by this reporting bias, HIV prevalence among the lower risk group would be overestimated. To correct this overestimate, we further assumed that HIV prevalence among "observed" non-lower risk (had 2+ partners p6m, reported 2+) was representative of HIV prevalence among "unobserved" non-lower risk (had 2+, reported 0–1). Thus, HIV prevalence among the "true" lower risk (had 0–1, reported 0–1) can be estimated as:

$$H_{01} = \frac{H - H_{2+}W'_{2+}}{W'_{01}} \tag{A.27}$$

where $H$ denotes HIV prevalence, and $W'$ denotes the adjusted proportions calculated in § A.3.10.

**Age Range** The model aims to capture the Swati population aged 15–49. While the 2006, 2016, and 2021 surveys provide data for ages 15–49, the 2011 survey was limted to ages 18–49. Since HIV prevalence is much lower among ages 15–17, the 2011 estimates would be biased high. We therefore adjusted all 2011 HIV prevalence estimates in as follows. Drawing on age-stratified data in 2006 [2] and 2011 [3], we assumed that HIV prevalence among ages 15–17 was 5% among women, 2% among men, and 3.5% overall. Next, we estimated the fraction of women aged 15–17 among all women aged 15–49 (13.5%), and likewise for men (15.4%) and overall (14.4%) [176]. We then estimated HIV prevalence among women, men, and overall for ages 15–49 using a weighted average of the 15–17 and 18–49 estimates. Finally, we computed the resulting relative reduction in HIV prevalence for women overall, and applied this reduction equally to the HIV prevalence estimates for lower risk and non-lower risk women, and likewise for men and the population overall.

The raw (unadjusted) estimates suggest that HIV prevalence strongly peaked between 2006 and 2016. After adjustment for respondent ages, 2011 estimates remained highest, but the magnitude of differences with 2006 and 2016 was reduced substantially. The largest reduction in HIV prevalence via adjustment was among lower risk women in 2011: from 38.2% to 30.8%, due to the modelled "addition" of women/girls aged 15–17 (lower HIV prevalence), and "subtraction" of women with 2+ partners p6m (higher HIV prevalence).

**Prevalence Ratios** Since risk heterogeneity is a key determinant of epidemic dynamics, it is important to capture HIV prevalence ratios across risk groups. For this objective, directly specifying prevalence ratio targets

Table A.7: Estimated HIV prevalence ratios in Eswatini

| Numerator [a] | Denominator [a] | Year | Ratio | (95% CI) | Used | Ref | Notes |
|---|---|---|---|---|---|---|---|
| Non-LR Women | LR Women | 2006 | 2.02 | (1.84, 2.34) | ✓ | [2] | b |
| | | 2011 | 1.54 | (1.47, 1.66) | ✓ | [3] | b |
| | | 2016 | 1.42 | (1.37, 1.51) | ✓ | [5] | b |
| Non-LR Men | LR Men | 2006 | 2.57 | (2.16, 5.28) | ✓ | [2] | b |
| | | 2011 | 1.24 | (1.20, 1.34) | ✓ | [3] | b |
| | | 2016 | 1.32 | (1.26, 1.45) | ✓ | [5] | b |
| FSW Overall | Women Overall | 2011 | 2.16 | (1.87, 2.50) | ✓ | [3,18] | b |
| | | 2021 | 1.86 | (1.68, 2.06) | ✓ | [21,22] | b |
| HR FSW | LR FSW | 2011 | 1.05 | (0.85, 1.27) | ✗ | [18] | c |

[a] LR: lower risk, reporting 0-1 partners p6m; Non-LR: lower risk, reporting 2+ partners p6m; FSW: female sex worker; HR/LR FSW: higher/lower risk FSW, as defined in § A.3.9; [b] mean and 95% CI estimated via Monte Carlo sampling; [c] per analysis in § A.3.9; see Table A.6 for more notes on data sources and adjustments.

is more efficient than using independent prevalence targets for lower risk and non-lower risk. Based on the available data, we defined the prevalence ratio targets in Table A.7.

###### A.4.2.2 HIV Incidence

Population-level HIV incidence was first measured in the 2011 Swaziland HIV Incidence Measurement Survey (SHIMS) via 6-month cohort (gold standard) [4,20], in which 145 seroconversions were observed among 11,232 re-tested (LTFU was 5.6%). SHIMS2 and SHIMS3 in 2016–17 and 2021 used the HIV-1 Limiting Antigen Enzyme Immunoassay (LAg EIA) "recency test", which detects infections acquired within the past 141 days, 95%CI: (119, 160) [23]; this LAg EIA incidence measure was validated during SHIMS1 [20]. Recency testing was also recently integrated into Eswatini standard of care [52].

Table A.8 summarizes the available HIV incidence data for Eswatini. Uncertainty around each estimate was modelled using a skewnormal or inverse gaussian distribution. As with prevalence, the 2011 estimates were adjusted for the missing 15–17 age range, this time assuming 2% and 0.4% annual incidence among women and men aged 15–17, respectively (extrapolating from age-stratified incidence estimates from [4]). The 2011 estimates for lower risk women and men were also adjusted for biased partner number reporting using the same approach as for HIV prevalence. Two incidence ratios were also defined (Table A.9).

No study of FSW in Eswatini estimated incidence directly, but [22] reported that 30 of 676 prevalent HIV infections among FSW were identified as recent via LAg EIA per national guidelines [5,52]. Using Eq. (A.14) with $\rho$ = 30/676 = 4.44% and $T$ = 130 days, we computed an incidence rate of $\lambda$ = 11.7% per year. We further estimated uncertainty for this rate by combining the 95% CI from $\rho \sim \text{Binom}(p = 4.44\%, N = 676)$ and $T \in (118,\ 140)$, yielding 95% CI for $\lambda$ of (8.3, 16.9).

Table A.8: Estimates of HIV incidence in Eswatini

| Population [a] | Year | N | Raw % | Adj % | (95% CI) | Used | Ref | Notes |
|---|---|---|---|---|---|---|---|---|
| Overall | 2016 | 9,476 | 1.48 | — | (0.96, 1.99) | ✓ | [5] | bc |
| | 2021 | 12,043 | 0.77 | — | (0.39, 1.15) | ✓ | [21] | d |
| Women Overall | 2011 | 5,486 | 3.1 | 2.94 | (2.52, 3.47) | ✓ | [4] | ef |
| | 2016 | 5,227 | 1.99 | — | (1.16, 2.80) | ✓ | [5] | bc |
| | 2021 | 6,985 | 1.45 | — | (0.69, 2.20) | ✓ | [21] | d |
| Men Overall | 2011 | 5,746 | 1.7 | 1.50 | (1.16, 1.84) | ✓ | [4] | ef |
| | 2016 | 4,249 | 0.99 | — | (0.39, 1.59) | ✓ | [5] | bc |
| | 2021 | 5,058 | 0.20 | — | (0.00, 0.48) | ✓ | [21] | d |
| LR Women | 2011 | 4,924 | 3.21 | 1.58 | (0.40, 2.24) | * | [4] | efg |
| Non-LR Women | 2011 | 93 | 10.10 | 9.62 | (4.76, 18.29) | * | [4] | ef |
| LR Men | 2011 | 3,855 | 1.64 | 0.76 | (0.01, 1.17) | * | [4] | efg |
| Non-LR Men | 2011 | 874 | 3.87 | 3.42 | (2.21, 4.94) | * | [4] | ef |
| FSW Overall | 2021 | 676 | 11.71 | — | (8.31, 16.92) | ✓ | [22] | b |

[a] LR: lower risk, reporting 0-1 partners p6m; Non-LR: lower risk, reporting 2+ partners p6m; FSW: female sex worker; [b] via HIV-1 Limiting Antigen recency testing; [c] 95% CI as reported from sampling adjustment; [d] N for survey overall; [e] via 6 month cohort (94.4% follow-up); [f] adjusted from ages 18–49 to 15–49 (see § A.4.2.1); [g] adjusted for biased reporting of risk behaviours (see § A.3.10 and § A.4.2.1); * used within incidence ratio only; all estimates used the skew normal distribution.

Table A.9: Estimated HIV incidence ratios in Eswatini

| Numerator [a] | Denominator [a] | Year | Ratio | (95% CI) | Used | Ref | Notes |
|---|---|---|---|---|---|---|---|
| Non-LR Women | LR Women | 2011 | 5.74 | (2.47, 22.26) | ✓ | [3] | b |
| Non-LR Men | LR Men | 2011 | 4.16 | (1.69, 23.09) | ✓ | [3] | b |

[a] LR: lower risk, reporting 0-1 partners p6m; Non-LR: lower risk, reporting 2+ partners p6m; FSW: female sex worker; [b] mean and 95% CI estimated via Monte Carlo sampling; see Table A.8 for more notes on data sources and adjustments.

###### A.4.2.3 HIV Cascade of Care

Table A.10 summarizes the available data for the HIV cascade of care in Eswatini, including estimates stratified by risk group where possible. Both conditional (*e.g.*, on ART among diagnosed, "90-90-90") and unconditional (*e.g.*, on ART among PLHIV, "90-81-73") cascade data were included, which is redundant but may improve calibration quality. Unlike HIV prevalence and incidence calibration targets, no adjustments were applied to these data. A recent meta-analysis [177] suggested substantial under-reporting of known HIV+ status, including 9 (4, 15)% among the population overall (10 studies), and 32 (22, 44)% among FSW specifically (2 studies). However, data from SHIMS2 [5] suggested much lower under-reporting (2.2%) in Eswatini.

Table A.10: Estimated HIV cascade of care in Eswatini

| Step [a] | Population [a] | Year | N | % | (95% CI) | Used | Ref | Notes |
|---|---|---|---|---|---|---|---|---|
| Diagnosed among PLHIV | Overall | 2011 | 5,807 | 62.6 | (61.4, 63.8) | ✓ | [81] | bc |
| | | 2016 | 2,417 | 86.1 | (84.7, 87.6) | ✓ | [5] | e |
| | Women overall | 2011 | 3,810 | 69.1 | (67.6, 70.6) | ✓ | [81] | b |
| | | 2016 | 1,690 | 90.2 | (88.6, 91.8) | ✓ | [5] | e |
| | Men overall | 2011 | 1,997 | 50.1 | (47.9, 52.3) | ✓ | [81] | b |
| | | 2016 | 727 | 77.3 | (74.0, 80.6) | ✓ | [5] | e |
| | FSW | 2011 | 313 | 74.1 | (61.7, 89.8) | ✓ | [178] | d |
| | | 2021 | 411 | 88.3 | (85.1, 91.2) | ✓ | [22] | bf |
| On ART among Diagnosed | Overall | 2011 | 3,635 | 52.1 | (50.5, 53.7) | ✓ | [81] | bcd |
| | | 2016 | 2,113 | 87.8 | (86.0, 89.6) | ✓ | [5] | e |
| | Women overall | 2011 | 2,633 | 48.0 | (46.1, 49.9) | ✓ | [81] | bd |
| | | 2016 | 1,532 | 87.5 | (85.4, 89.6) | ✓ | [5] | e |
| | Men overall | 2011 | 1,002 | 62.7 | (59.7, 65.7) | ✓ | [81] | bd |
| | | 2016 | 581 | 88.4 | (85.2, 91.6) | ✓ | [5] | e |
| | FSW | 2011 | 174 | 36.9 | (30.1, 44.2) | ✓ | [178] | |
| | | 2021 | 363 | 97.5 | (95.7, 98.9) | ✓ | [22] | bf |
| On ART among PLHIV | Overall | 2011 | 5,807 | 31.9 | (30.7, 33.1) | ✓ | [81] | bc |
| | | 2016 | 2,417 | 75.6 | (73.6, 77.5) | ✓ | [5] | e |
| | Women overall | 2011 | 3,810 | 33.2 | (31.7, 34.7) | ✓ | [81] | b |
| | | 2016 | 1,690 | 78.9 | (76.8, 81.1) | ✓ | [5] | e |
| | Men overall | 2011 | 1,997 | 31.4 | (29.4, 33.4) | ✓ | [81] | b |
| | | 2016 | 727 | 68.3 | (64.7, 72.0) | ✓ | [5] | e |
| | FSW | 2011 | 313 | 27.4 | (20.9, 35.7) | ✓ | [178] | d |
| | | 2021 | 411 | 86.1 | (82.6, 89.3) | ✓ | [22] | bf |
| VLS among On ART | Overall | 2016 | 1,858 | 90.3 | (89.0, 91.6) | ✓ | [5] | e |
| | Women overall | 2016 | 1,342 | 91.4 | (89.9, 92.8) | ✓ | [5] | e |
| | Men overall | 2016 | 516 | 87.6 | (84.4, 90.9) | ✓ | [5] | e |
| VLS among PLHIV | Overall | 2016 | 2,417 | 68.2 | (66.1, 70.4) | ✓ | [5] | e |
| | | 2021 | 2,854 | 86.6 | (85.0, 88.1) | ✓ | [21] | g |
| | Women overall | 2016 | 1,690 | 72.1 | (69.7, 74.5) | ✓ | [5] | e |
| | | 2021 | 2,207 | 88.6 | (87.0, 90.2) | ✓ | [21] | g |
| | Men overall | 2016 | 727 | 59.9 | (56.1, 63.7) | ✓ | [5] | e |
| | | 2021 | 789 | 82.4 | (79.3, 85.5) | ✓ | [21] | g |

[a] PLHIV: people living with HIV; ART: antiretroviral therapy; VLS: HIV viral load suppressed, defined as $\leq$ 1000 RNA copies/mL in [5]; FSW: female sex worker; [b] 95% CI inferred from N; [c] estimated from combining women & men; [d] estimated from conditional steps, with 95% CI via simulation; [e] 95% CI as reported from sampling adjustment; [f] not RDS-adjusted; [g] N estimated from HIV prevalence; [2] did not provide any appropriate cascade data.

#### Appendix B

### Supplementary Results

#### B.1 Model Calibration

This section presents the results of model calibration under the *Effective Partnerships Adjustment*, including: § B.1.1: posterior distributions of calibrated parameters (Table A.5); § B.1.2: model outputs *vs* associated calibration targets; and § B.1.3 modelled patterns of transmission among risk groups in Eswatini over time.

##### B.1.1 Posterior Parameter Distributions

Figure B.1 illustrates the distributions of calibrated model parameters, stratified by prior (IMIS iteration 0) *vs* posterior (resampled 1000 parameter sets). Most prior *vs* posterior distributions were significantly different (Anderson-Darling Test [179]). Such differences tended to favour increased overall transmission (*e.g.*, `beta_0`, `dur_acute`, `Rbeta_acute`), including via casual partnerships (*e.g.*, `C12m_cas_wm`, `RC_cas_cli.wm`, `RF_cas.msp`), but decreased accumulation of HIV prevalence among FSW (*e.g.*, `F_swr`, `dur_fsw`). These findings may reflect the difficulty of recreating high overall HIV prevalence despite relatively small prevalence ratios (~2) for FSW *vs* women overall. Relative transmission from men to women (`Rbeta_vi_rec`) was also high, perhaps because our model lacks age, and age-gender differences in sexual activity and/or mixing might better explain gender differences in HIV prevalence.

Figure B.2 further illustrates bivariate rank correlations among posterior parameter values. Several sex work condom use parameters had strong positive correlations, likely induced via relational constraints (see § A.4.1). By contrast, the strongest negative correlation was between the rate of diagnosis for non-FSW women and a global relative rate for diagnosis, since these parameters served very similar roles.

##### B.1.2 Model Fits *vs* Calibration Targets

This section presents the estimates of key model outputs from the 1000 model fits (posterior parameter sets), with comparison to the associated calibration targets.

**Log Likelihoods** Figure B.3 illustrates the distributions of log likelihoods for initial prior / Latin hypercube samples (100,000), all IMIS iteration samples (1,000,000), and posterior samples (1000). See § A.4.1 regarding our adjusted IMIS weights due to the large variance in log likelihoods.

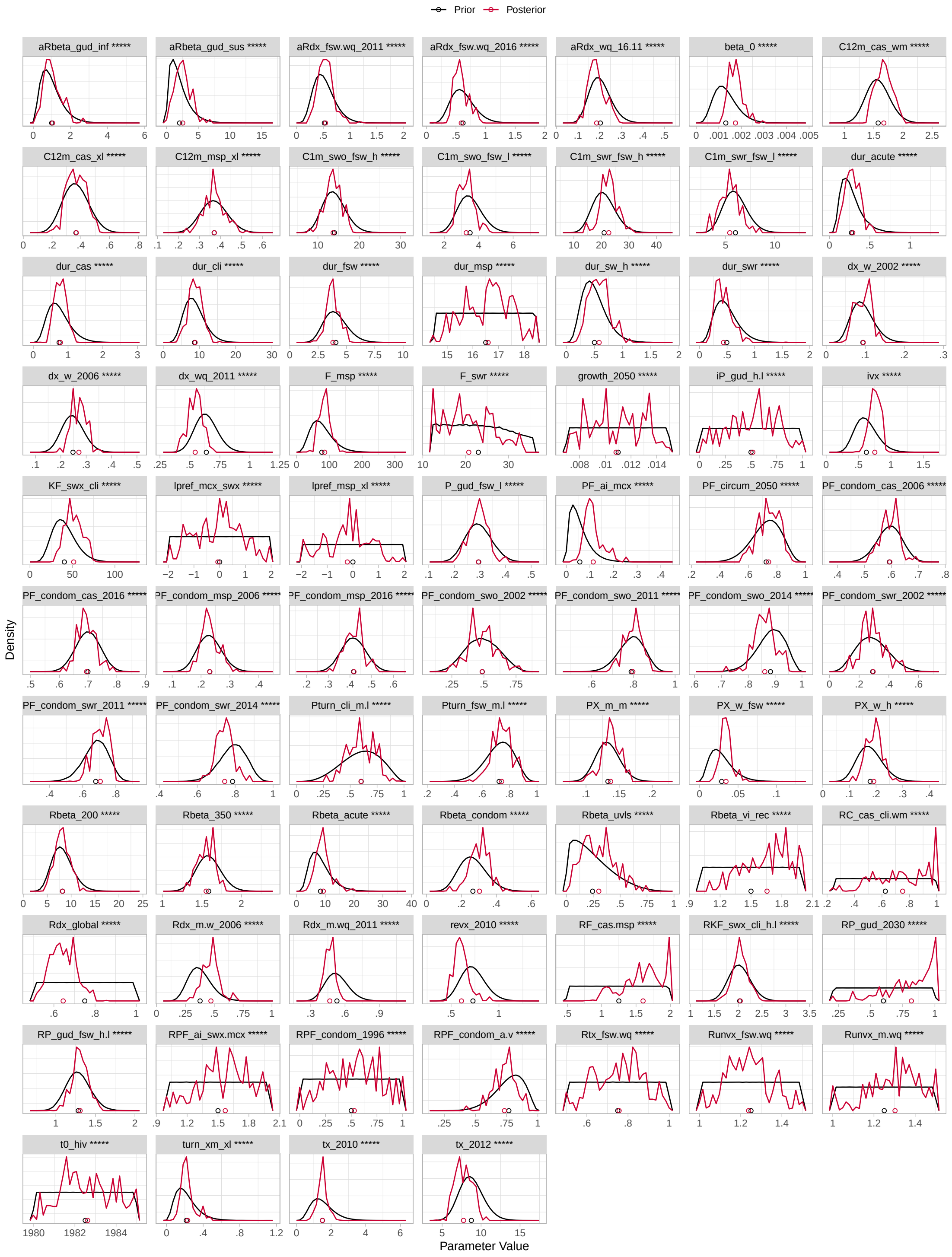

Figure B.1: Distributions of calibrated model parameters, stratified by prior (all initial samples) *vs* posterior (1000 resamples)

Table A.5 gives parameter definitions; lines and circles: normalized density and means for each parameter; * denote significance of QN rank score test [180] for comparing distributions, where: $p < 0.1$: *, $p < 0.01$: **, etc.

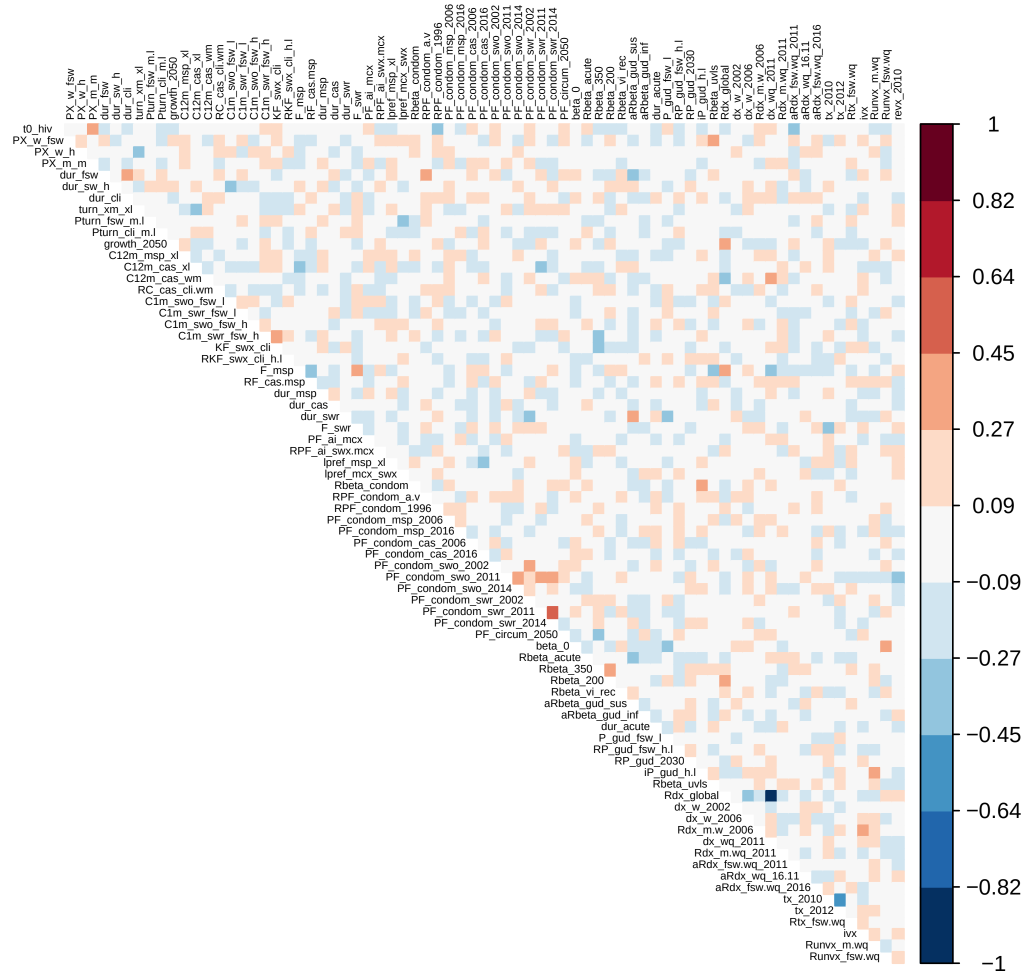

Figure B.2: Rank correlations among posterior samples for calibrated model parameters

Table A.5 gives parameter definitions.

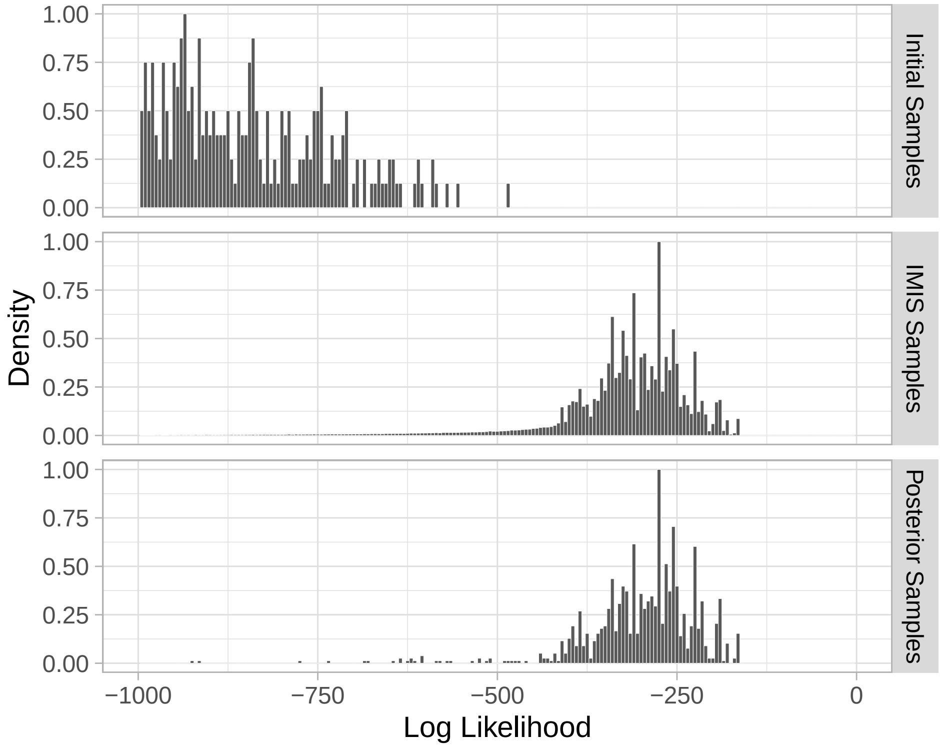

Figure B.3: Distribution of log likelihoods across stages of calibration

Only log likelihoods ≥ −1000 shown.

###### B.1.2.1 HIV Prevalence & Incidence

Figure B.4 illustrates modelled HIV prevalence (a), prevalence ratios (b), incidence (c), and incidence ratios (d) among selected risk groups, with the associated calibration targets. Overall, model estimates agree well with the available calibration targets, with the following shortcomings. Relative to the calibration targets, the model tends to overestimate HIV prevalence among FSW in 2014, slightly overestimate HIV prevalence among men overall, and overestimate HIV prevalence ratios for non-lowest *vs* lowest activity men. The discrepancies for men could arise due to population-level surveys failing to reach men at higher risk (*e.g.*, with high mobility) [181], considering that participation rates were consistently lower for men *vs* women (§ A.3.2).

Few data are available to validate the modelled early epidemic dynamics. Modelled incidence among women and men peaked rapidly after introduction of HIV (Figure B.4c), corresponding to rapid acquisition and saturation among FSW and clients. Modelled incidence and prevalence continued to increase approximately linearly over 1990–2010, reflecting a balance of would-be exponential epidemic growth *vs* build-up of mitigating factors, such as increasing condom use (Figure B.10), male circumcision (Figure B.9), ART coverage (Figure B.5, and accumulation of seroconcordant partnerships (Figure B.11). These trends can be compared with HIV prevalence from Eswatini antenatal care clinics over the same period (Figure B.4e), which suggest similar

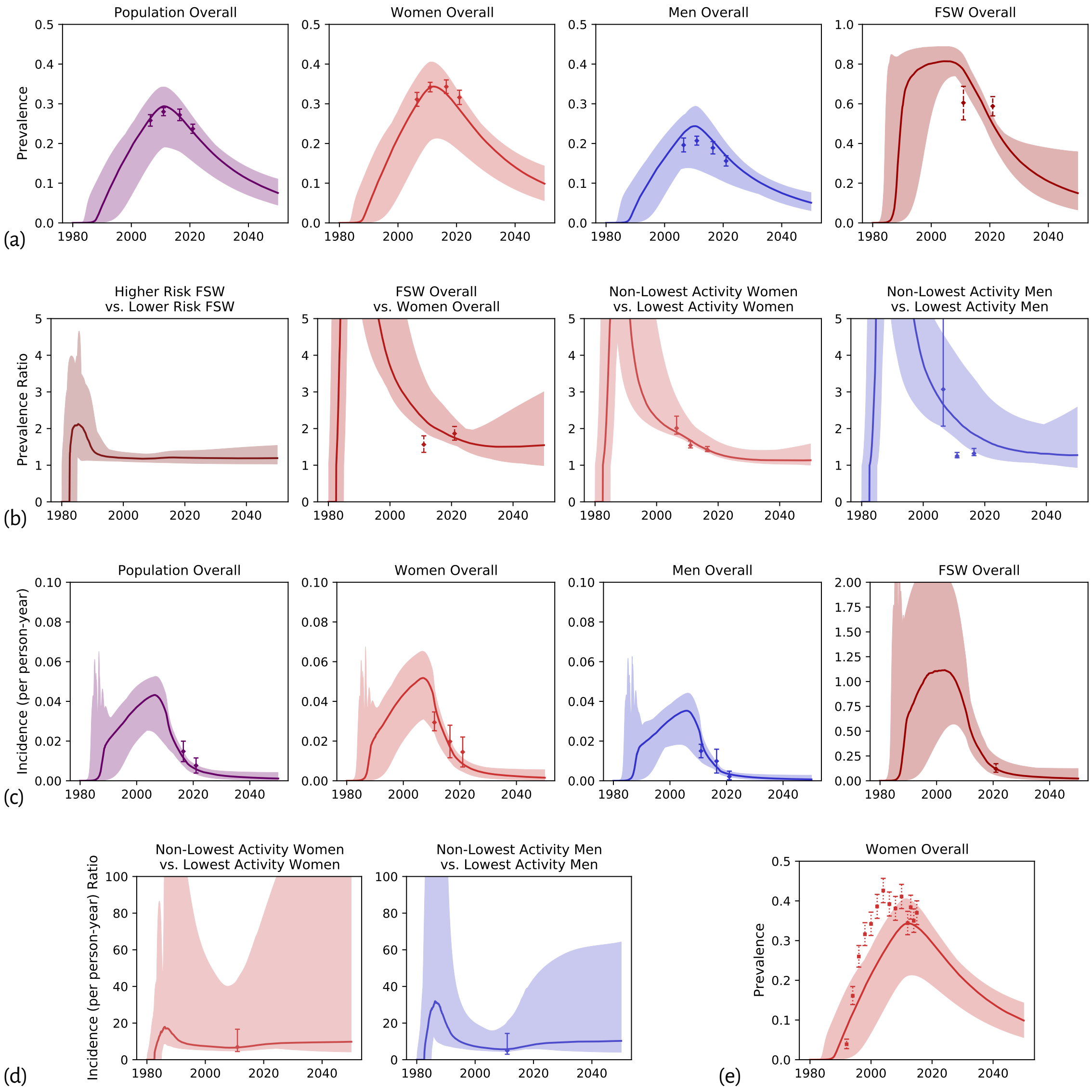

Figure B.4: Modelled HIV prevalence, incidence, and ratios thereof among selected risk groups, and associated calibration targets

1000 model fits (posterior parameter sets); ribbon and curve: range and median of model fits; points and error bars: mean and 95% CI for each calibration target; (e) targets from antenatal care data [61,98] (not used for calibration)

trends.[22] Decline of HIV incidence and prevalence after 2010 can likely be attributed to rapid ART scale-up (see § B.1.2.2) and further increases in condom use (Figure B.10). Although modelled incidence declined rapidly, prevalence remained relatively higher due to increased survival of PLHIV with ART. In some model fits, prevalence among FSW declined faster than among women overall, likely due to high turnover of women in sex work.

###### B.1.2.2 ART Cascade

Figure B.5 illustrates the modelled ART cascade among selected risk groups, including both conditional and unconditional cascade steps, and the associated calibration targets. The model estimates agree quite well with these targets, for all risk groups. The non-monotonic increasing proportions virally suppressed among treated PLHIV reflect major changes in treatment eligibility (see § A.3.8.2), which caused influxes of newly ART-eligible PLHIV to temporarily decrease the proportions virally suppressed among treated PLHIV. Figure B.6 also illustrates rate of HIV diagnosis (a) and ART initiation (b) among selected risk groups over time.

###### B.1.2.3 Additional Model Outputs

Figure B.7 illustrates the relative sizes of higher and lower risk FSW and clients over time, showing relative stability despite disproportionate HIV-attributable mortality. Figure B.8 illustrates the total population size over time, showing agreement with available data [28] (see § A.3.12.1). Figure B.9 illustrates the modelled proportion of men aged 15–49 who are circumcised, including uncertainty about future trends (see § A.3.5.1). Figure B.10 illustrates modelled trends in condom use within different partnership types (see § A.3.5.2). Figure B.11 illustrates the modelled transmission-driven seroconcordance proportion for different partnership types (see § A.2.4), as defined in Eq. (A.9) with denominator (b).

##### B.1.3 Who Infected Whom

As further model validation, and to gain insights into the modelled networks of transmission, this section presents several summaries of "who infected whom" — *i.e.*, distributions of yearly infections stratified by the transmitting group, acquiring group, and partnership type. Throughout the section, the numbers of yearly infections shown are obtained from the median value across all 1000 model fits.

Figure B.12 illustrates the total numbers and proportions of modelled yearly infections transmitted from (a) and acquired among (b) modelled risk groups. Figure B.13 then gives the *ratio* of yearly infections transmitted *vs* acquired. Figure B.14 stratifies yearly infections by partnership type, while Figure B.15 illustrates the complete transmission network every 10 years from 1990.

[22] Antenatal care data were not used as calibration targets because such data are known to overestimate HIV prevalence among women overall due to non-representative sampling [182,183].

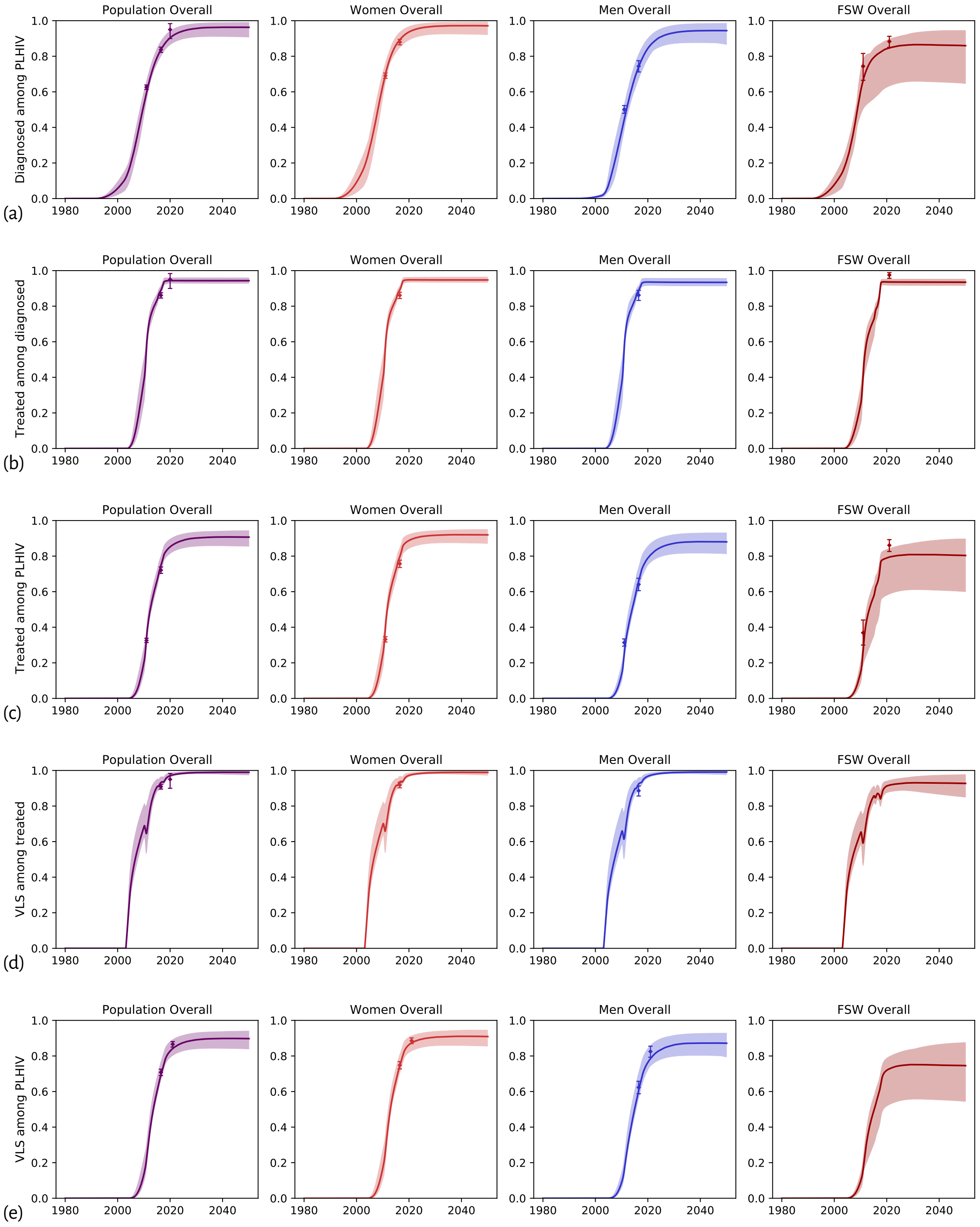

Figure B.5: Modelled ART cascade among selected risk groups and associated calibration targets

PLHIV: people living with HIV; VLS: viral load unsuppressed; 1000 model fits (posterior parameter sets); ribbon and curve: range and median of model fits; points and error bars: mean and 95% CI for each calibration target.

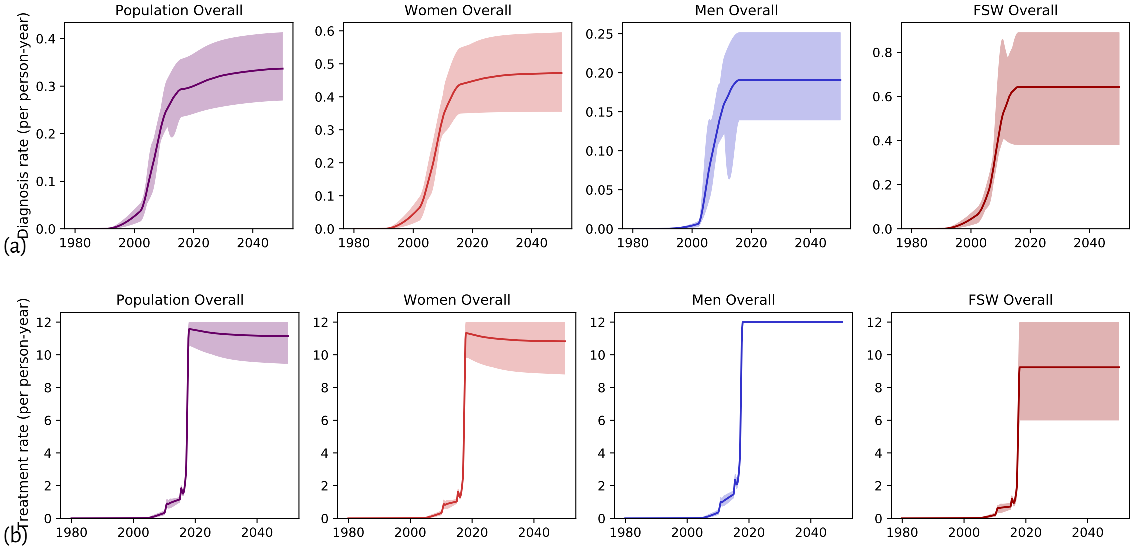

Figure B.6: Modelled rates of HIV diagnosis and ART initiation among selected risk groups over time

1000 model fits (posterior parameter sets); ribbon and curve: range and median of model fits.

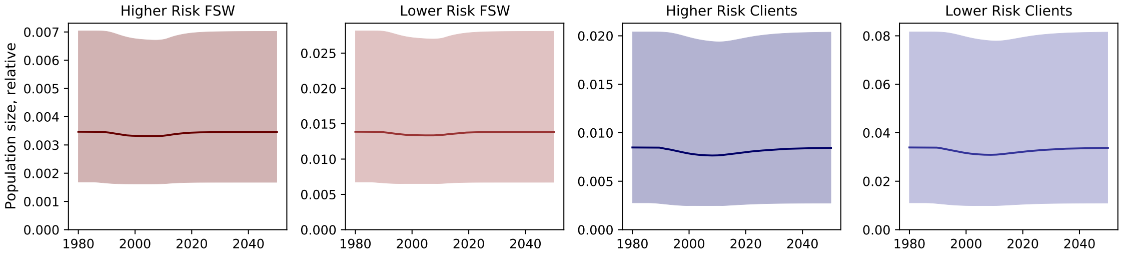

Figure B.7: Modelled relative sizes of selected risk groups over time

1000 model fits (posterior parameter sets); ribbon and curve: range and median of model fits.

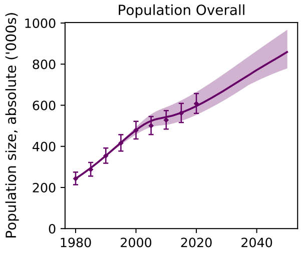

Figure B.8: Modelled total population aged 15–49 and associated calibration targets

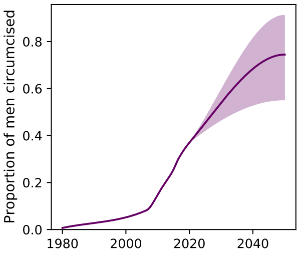

Figure B.9: Modelled proportion of men aged 15–49 who are circumcised

1000 model fits (posterior parameter sets); ribbon and curve: range and median of model fits; points and error bars: mean and 95% CI for each calibration target.

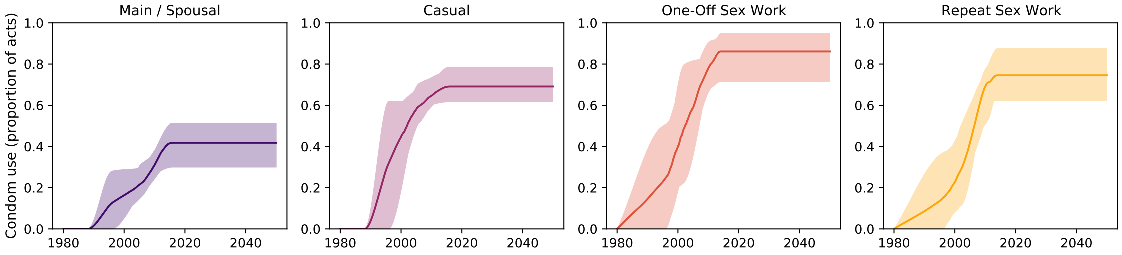

Figure B.10: Modelled condom use within different partnership types

1000 model fits (posterior parameter sets); ribbon and curve: range and median of model fits.

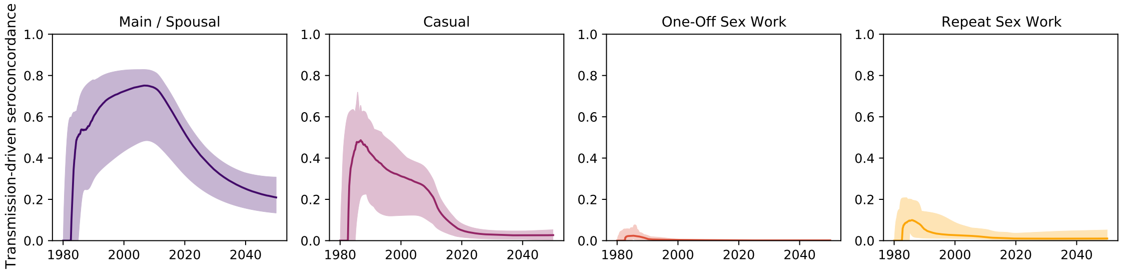

Figure B.11: Modelled transmission-driven seroconcordance within different partnership types

1000 model fits (posterior parameter sets); ribbon and curve: range and median of model fits.

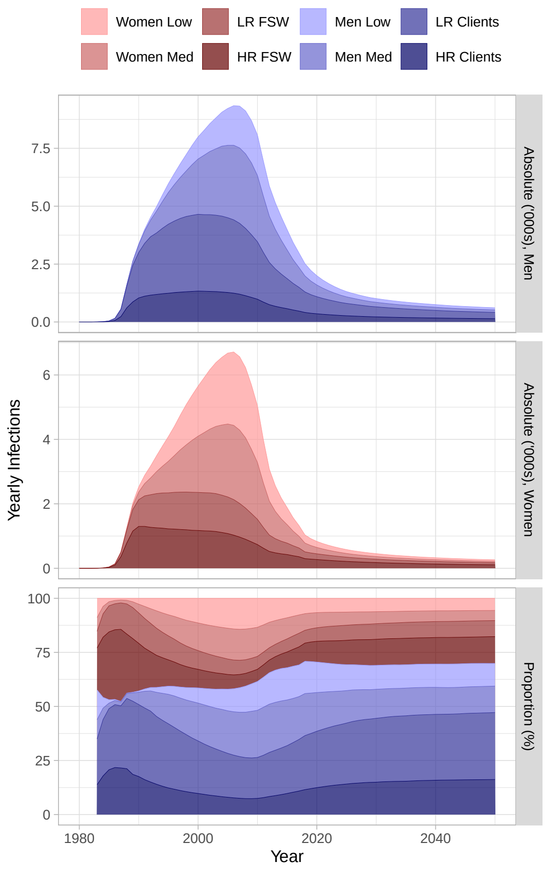

(a) Transmitted from

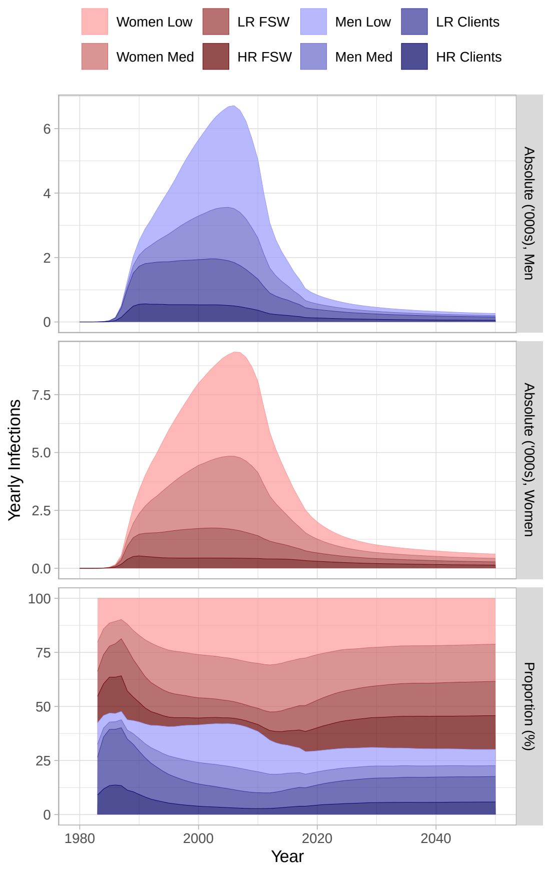

(b) Acquired among

Figure B.12: Modelled yearly HIV infections (a) transmitted from and (b) acquired among risk groups

Low: lowest activity; Med: medium activity; LR/HR: lower/higher risk; FSW: female sex workers; Clients: of FSW; median numbers of infections across all model fits shown.

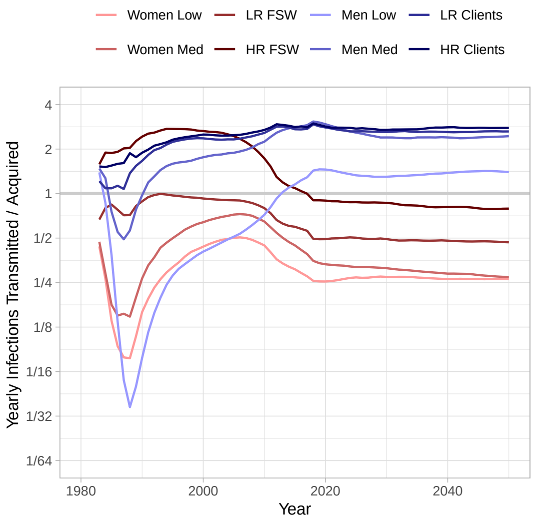

Figure B.13: Ratio of modelled yearly infections transmitted from *vs* acquired among risk groups

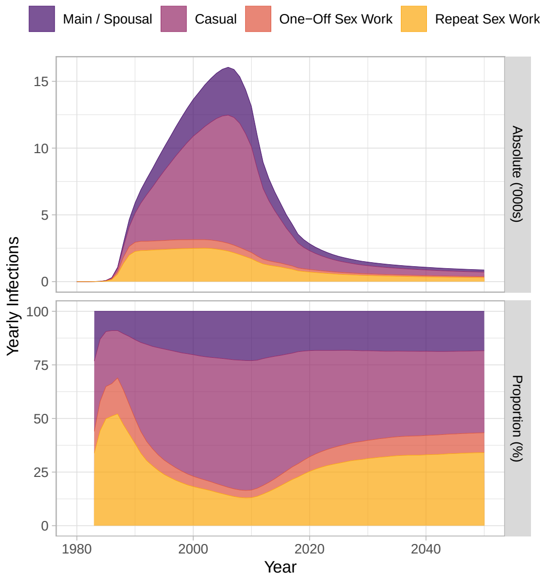

Figure B.14: Modelled yearly HIV infections transmitted via different partnership types

Low: lowest activity; Med: medium activity; LR/HR: lower/higher risk; FSW: female sex workers; Clients: of FSW; median numbers of infections across all model fits shown.

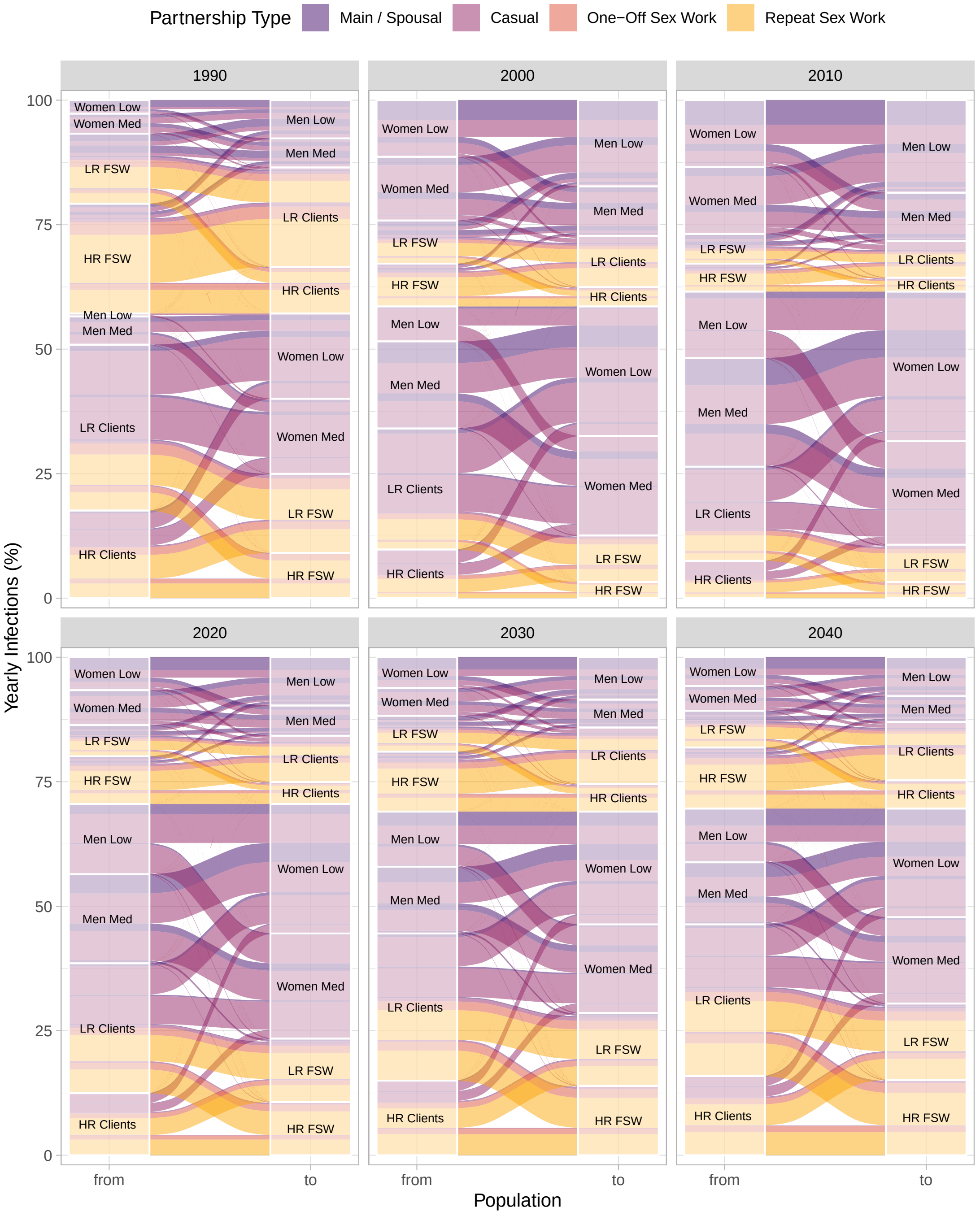

Figure B.15: Alluvial diagram showing proportions of all yearly infections (ribbons) transmitted from (left) to (right) modelled risk groups, stratified by partnership type (color) and year (facets)

Low: lowest activity; Med: medium activity; LR/HR: lower/higher risk; FSW: female sex workers; Clients: of FSW; median numbers of infections across all model fits shown.

#### B.2 Objectives

##### B.2.1 Objective 1

Figure B.17 illustrates cascade attainment over time across the base case and counterfactual scenarios (Table 2) for FSW, clients, everyone else, and the population overall. Transient declines in VLS among treated around 2010 correspond to expanding ART eligibility.

Figure B.16 illustrates overall HIV incidence over time across scenarios. As in § B.1.3, Figure B.18 illustrates the distributions of additional infections over time *vs* the base case across counterfactual scenarios (Table 2), stratified by partnership type, transmitting group, and acquiring group.

##### B.2.2 Objective 2

Figure B.19 illustrates the distributions of cascade attainment by 2020 for FSW, clients, everyone else, and the population overall, in the 10,000 randomly generated counterfactual scenarios for the Objective 2 regression analysis. Figure B.20 illustrates the corresponding directed acyclic graph as reflected in Eq. (2). Figure B.21 then illustrates the simulated *vs* regression-estimated cumulative additional infections (CAI) and additional incidence rate (AIR), which supports the use of linear regression despite minor heteroskedasticity.

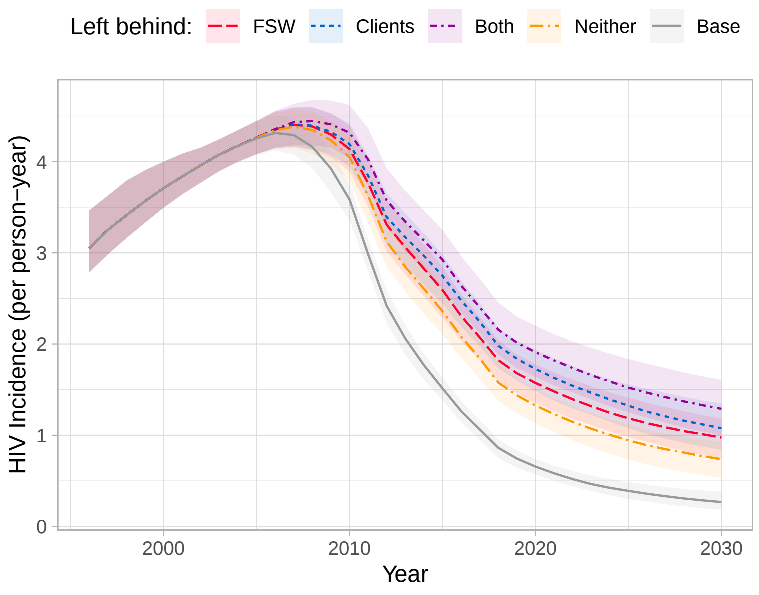

Figure B.16: Overall HIV incidence over time across scenarios

Base case: 95-95-95 by 2020; counterfactual scenarios: 80-80-90 overall by 2020, with reduced cascade (60-40-80: left behind) among FSW, clients of FSW, both, or neither; ribbon and curve: range and median of model fits.

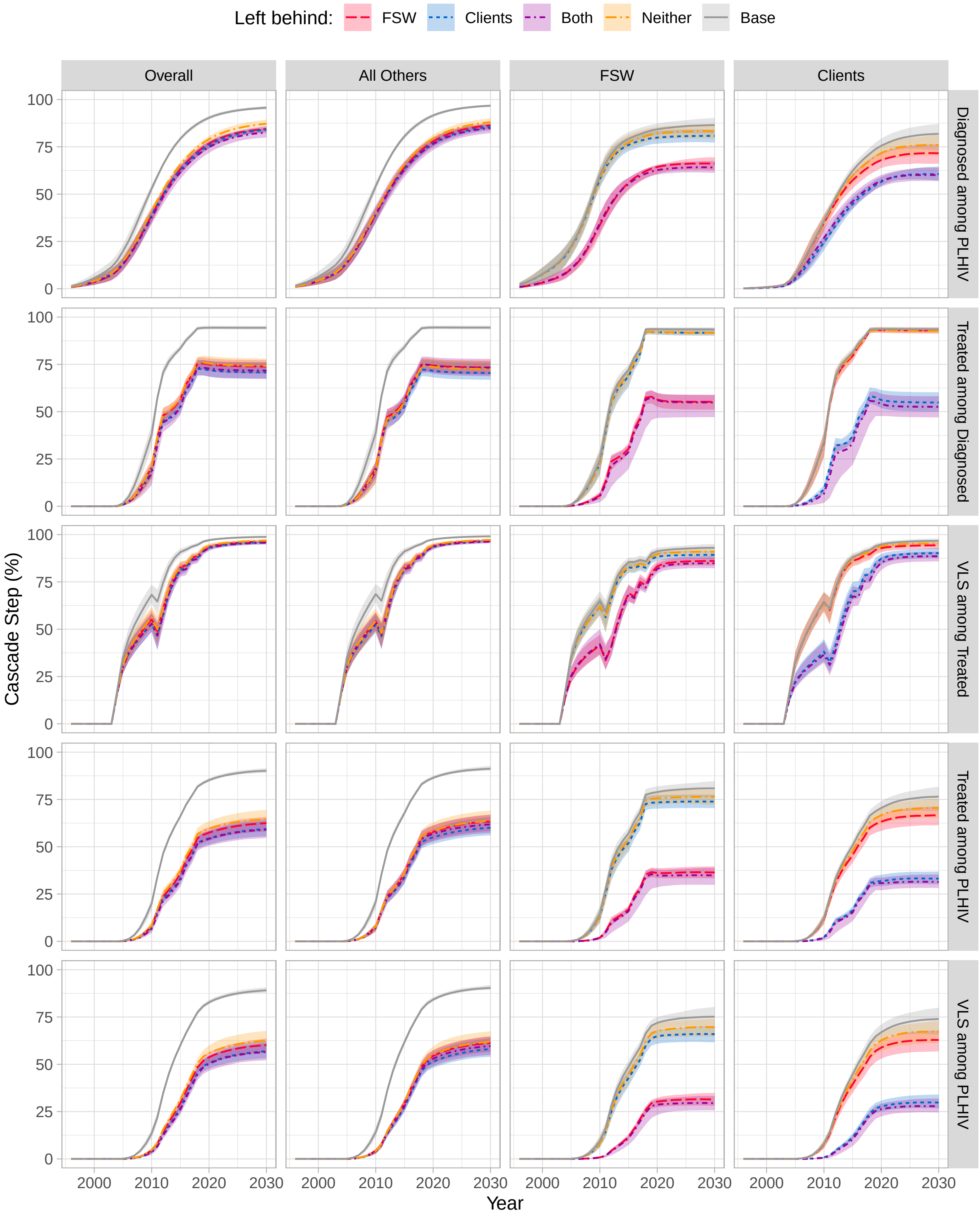

Figure B.17: Cascade attainment over time across scenarios

FSW: female sex workers; Clients: of FSW; All Others: all women and men not involved in sex work; PLHIV: people living with HIV; VLS: viral load unsuppressed; Base case: 95-95-95 by 2020; counterfactual scenarios: 80-80-90 overall by 2020, with reduced cascade (60-40-80: left behind) among FSW, clients of FSW, both, or neither; ribbon and curve: range and median of model fits.

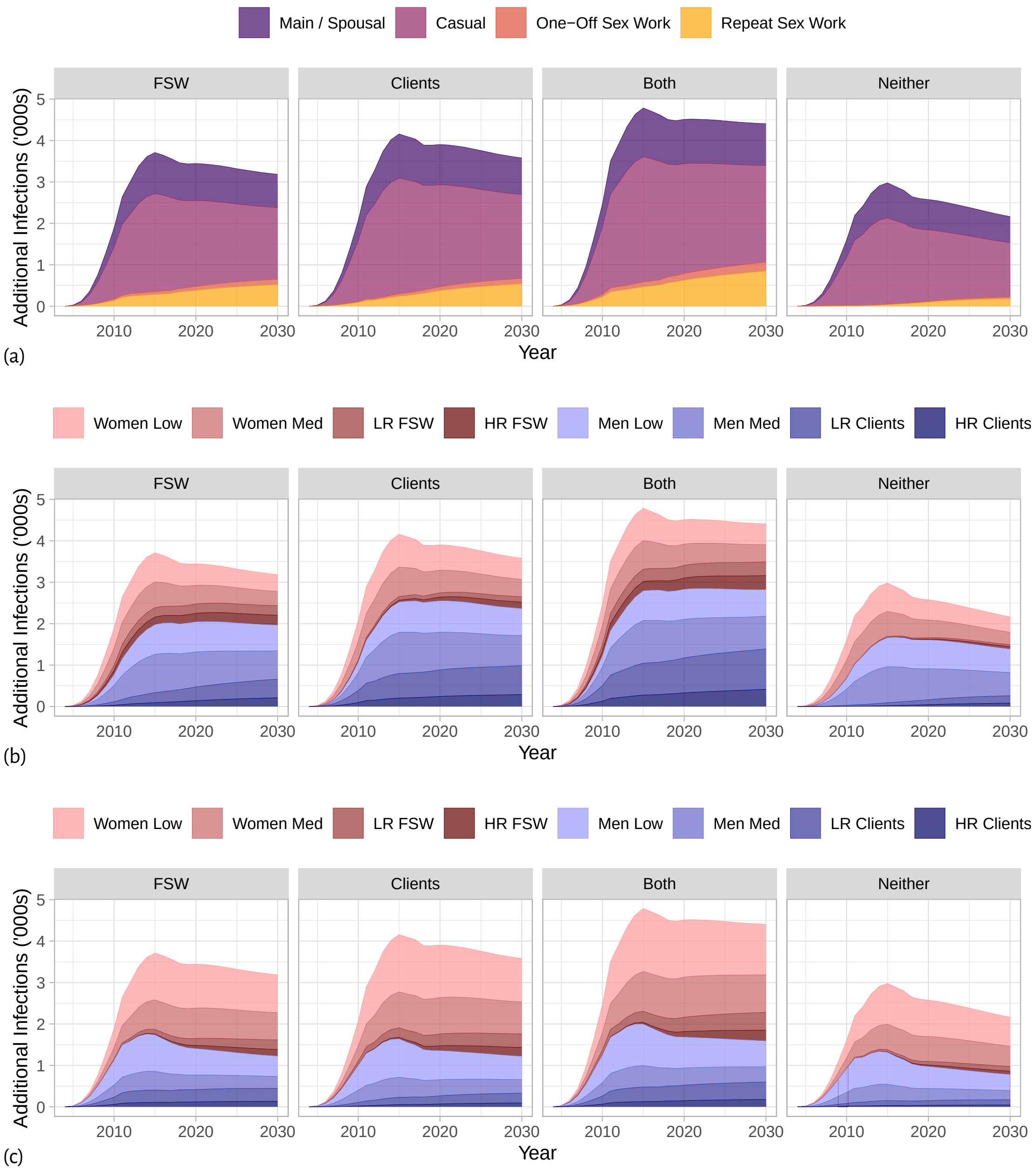

Figure B.18: Additional infections in each counterfactual scenario *vs* the base case, stratified by: (a) partnership type, (b) transmitting group, and (c) acquiring group

Base case: 95-95-95 by 2020; counterfactual scenarios: 80-80-90 overall by 2020, with reduced cascade (60-40-80: left behind) among FSW, clients of FSW, both, or neither; median numbers of infections across all model fits shown.

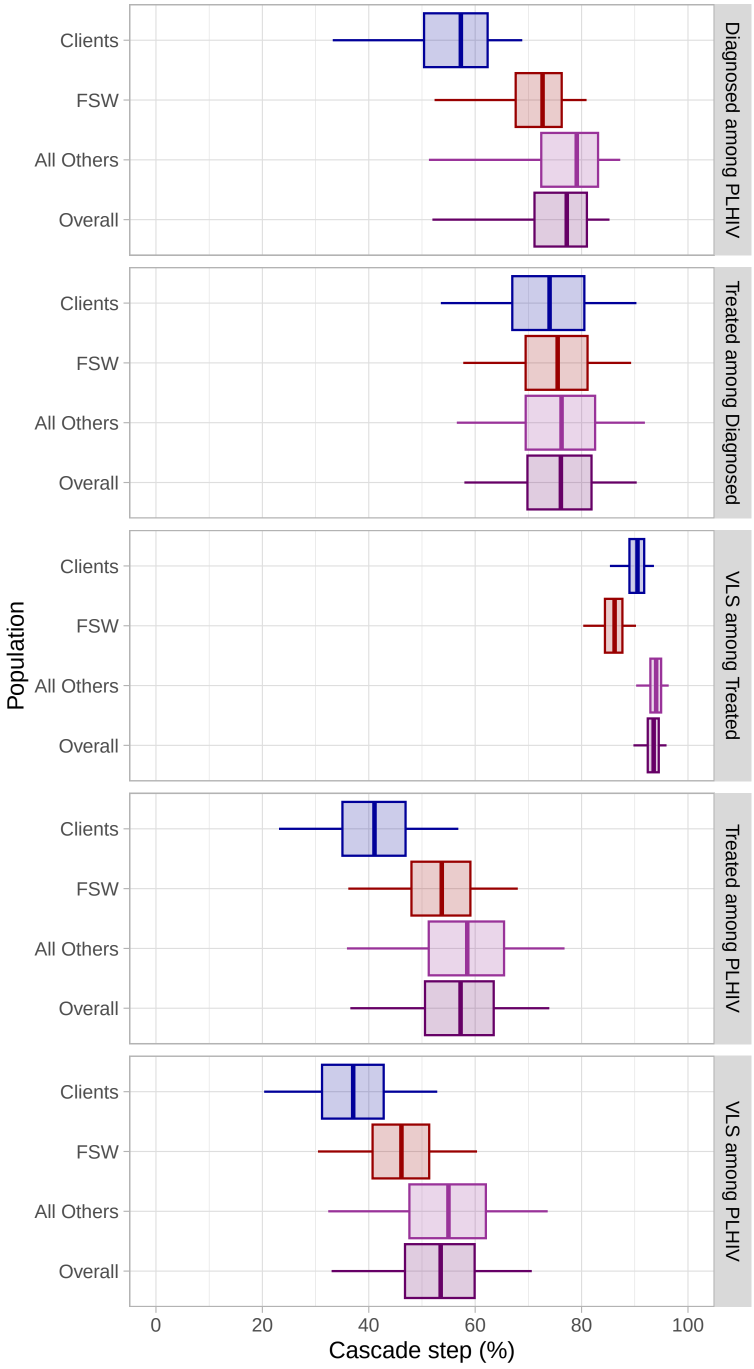

Figure B.19: Cascade attainment by 2020 across 10,000 randomly generated counterfactual scenarios

FSW: female sex workers; Clients: of FSW; All Others: all women and men not involved in sex work; PLHIV: people living with HIV; VLS: viral load unsuppressed; whiskers, boxes, and midlines: 95% CI, 50% CI, median of model fits.

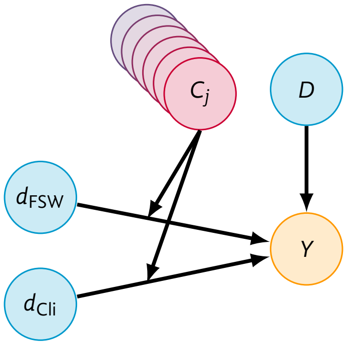

Figure B.20: Directed acyclic graph (DAG) for inferring the epidemic conditions under which differential viral suppression across subpopulations matters most

$Y$: cumulative additional infections (CAI) or additional incidence rate (AIR) by 2030; $D$: difference in population-overall viral non-suppression in counterfactual *vs* base case scenario; $d_i$: difference in subpopulation-$i$-specific viral non-suppression *vs* population overall within counterfactual scenario; $C_j$: epidemic conditions (effect modifiers of $d_i$).

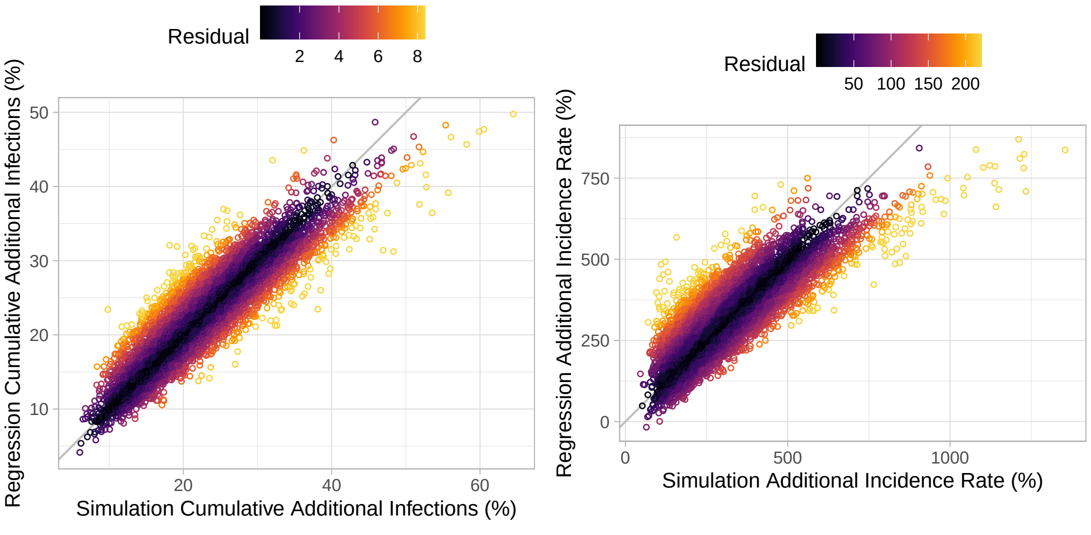

(a) Cumulative additional infections

(b) Additional incidence rate

Figure B.21: Simulated *vs* regression-estimated outcomes, and corresponding residuals for Objective 2.

Outcomes computed for 2030 *vs* base case; regression models defined in Eq. (2).

#### B.3 Research in Context

This section presents results of the scoping review for Box 1, adapted and updated from [108].

##### B.3.1 Definitions & Criteria

Table B.1 gives the inclusion/exclusion criteria for our review, while key definitions are given below:

**Non-linear model** Any model where: the number of infections projected at time $t$ is an iterative function of the number of infections previously projected by the model before time $t$.

**Sub-Saharan Africa** (SSA) any country in the UN regions of East, South, Central, and West Africa, plus South Sudan (see Table B.2 for full country list).

**90-90-90+ scenario** Among all people living with HIV (regardless of CD4 count): 90+% are diagnosed, of whom 90+% are on ART, of whom 90+% are virally suppressed — or 73+% are effectively modelled as virally suppressed — achieved for the population overall by any year, and without any other interventions beyond status quo. We further distinguish between *named* 90-90-90+ scenarios and *effectively* 90-90-90+ scenarios based on modelled rates.

**Quantified prevention impacts** HIV incidence rate reduction or cumulative infections averted among the whole population in 90-90-90+ *vs* status quo scenario over any time horizon.

**Evidence before this study** We adapted and updated a scoping review of HIV transmission modelling studies examining the prevention impacts of antiretroviral therapy (ART) cascade scale-up in Sub-Saharan Africa. Our search yielded (prior + update) 1367 + 373 unique studies, of which: 7 + 5 examined the prevention impacts of achieving the UNAIDS 90-90-90 goals or greater (Table B.3). Some of these studies considered differences in baseline rates of diagnosis, treatment initiation, and/or treatment failure/discontinuation — mainly by sex, age, and occasionally risk. Three studies specifically examined scenarios of unequal cascade attainment across subpopulations, predicting substantially more infections when subpopulations at higher risk were left behind. However, these studies did not maintain consistent overall cascade across scenarios — *i.e.,* each scenario reached a different cascade for the population overall — and considered only hypothetical scenarios.
**Added value of this study** We developed and calibrated a detailed model of heterosexual HIV transmission and observed ART cascade scale-up in Eswatini, drawing on population-level and female sex worker (FSW)-specific surveys. Eswatini achieved 95-95-95 overall and similar among FSW by 2020. We estimated what would have happened if overall cascade scale-up had been slower (reaching only 80-80-90 by 2020), and inequalities in the cascade among FSW and/or clients had not been addressed (reaching 60-40-80 by 2020). We found that such cascade inequalities would have led to 40–149% additional HIV infections in the overall population by 2030, as compared to equitable scale-up.
**Implications of all the available evidence** The population-level prevention benefits of achieving 95-95-95 overall are undermined by inequalities in the HIV cascade, especially when populations experiencing disproportionate transmission risks are left behind, including but not limited to FSW and their clients.

Box 1: Research in context

Table B.1: Criteria for inclusion and exclusion

| Include | Exclude |
|---|---|
| **Publication Type** | |
| • English language<br>• peer-reviewed research article | • non-English<br>• non-peer-reviewed<br>• review article<br>• textbook, grey literature<br>• opinions, comments, correspondence<br>• conference abstracts and proceedings |
| **Modelling of HIV Transmission** | |
| • population-level dynamics<br>• sexual HIV transmission model<br>• non-linear HIV transmission model[*] | • only within-host/cellular/protein modelling<br>• no sexual HIV transmission modelled<br>• HIV transmission model is linear<br>• HIV incidence is fixed or imported |
| **Context & Objectives** | |
| • any region in Sub-Saharan Africa (SSA)[*]<br>• includes 90-90-90+ scenario[*]<br>• includes base-case reflecting status quo<br>• report prevention impacts of 90-90-90+[*] | • only regions outside SSA[*]<br>• only theoretical context<br>• no 90-90-90+ scenario[*]<br>• no base-case scenario<br>• did not report ART prevention impacts[*]<br>• intervention/budget optimization<br>• viral load monitoring or regimen switching |

[*] See definitions in B.3.1.

##### B.3.2 Search Strategy & Hits

We searched MEDLINE and EMBASE via Ovid on: 2020 March 20 (studies published before 2020) and 2023 November 23 (studies published in/after 2020). Table B.2 gives the search terms and hits from each search, where `term/` denotes a MeSH term, and `.mp` searches the main text fields, including title, abstract, and heading words. Duplicate studies were removed automatically by Ovid. Figure B.22 illustrates the PRISMA diagram for study identification.

Table B.2: Search terms and hits

| | Hits | Term |
|---|---|---|
| M1 | 259,932 | `model, theoretical/` |
| M2 | 357,388 | `model, biological/` |
| M3 | 354,078 | `computer simulation/` |
| M4 | 210,102 | `patient-specific modeling/` |
| M5 | 84,649 | `monte carlo method/` |
| M6 | 70,865 | `exp stochastic processes/` |
| M7 | 621,337 | `(model* ADJ3 (math* OR transmission OR dynamic* OR epidemi* OR compartmental OR deterministic OR individual OR agent OR network OR infectious disease* OR markov OR dynamic* OR simulat*)).mp.` |

| | | |
|---|---|---|
| M8 | 1,648,338 | OR/ M1-M7 |
| H1 | 331,119 | exp HIV/ |
| H2 | 748,853 | exp HIV infections/ |
| H3 | 880,774 | (HIV OR HIV1* OR HIV2* OR HIV-1* OR HIV-2*).mp. |
| H4 | 426,755 | hiv infect*.mp. |
| H5 | 627,422 | (human immun*deficiency virus OR human immun* deficiency virus).mp. |
| H6 | 234,482 | exp Acquired Immunodeficiency Syndrome/ |
| H7 | 249,998 | (acquired immun*deficiency syndrome OR acquired immun* deficiency syndrome).mp. |
| H8 | 1,105,191 | OR/ H1-H7 |
| G1 | 4550 | Angola/ OR Angola.mp. |
| G2 | 11,475 | Benin/ OR Benin.mp. |
| G3 | 7688 | Botswana/ OR Botswana.mp. |
| G4 | 13,091 | Burkina Faso/ OR Burkina Faso.mp. |
| G5 | 2713 | Burundi/ OR Burundi.mp. |
| G6 | 21,726 | Cameroon/ OR Cameroon.mp. |
| G7 | 1598 | Cape Verde/ OR Cape Verde.mp. |
| G8 | 28,956 | Central African Republic/ OR Central African Republic.mp. OR CAR.ti. |
| G9 | 4129 | Chad/ OR Chad.mp. |
| G10 | 1320 | Comoros/ OR Comoros.mp. |
| G11 | 18,397 | Democratic Republic of the Congo/ OR Democratic Republic of the Congo.mp. OR DRC.mp. |
| G12 | 1209 | Djibouti/ OR Djibouti.mp. |
| G13 | 1410 | Equatorial Guinea/ OR Equatorial Guinea.mp. |
| G14 | 1941 | Eritrea/ OR Eritrea.mp. |
| G15 | 64,626 | Ethiopia/ OR Ethiopia.mp. |
| G16 | 5383 | Gabon/ OR Gabon.mp. |
| G17 | 7939 | Gambia/ OR Gambia.mp. |
| G18 | 37,148 | Ghana/ OR Ghana.mp. |
| G19 | 370,653 | Guinea/ OR Guinea.mp. |
| G20 | 3084 | Guinea-Bissau/ OR Guinea-Bissau.mp. |
| G21 | 11,852 | Cote d'Ivoire/ OR Cote d'Ivoire.mp. OR Ivory Coast.mp. |
| G22 | 60,705 | Kenya/ OR Kenya.mp. |
| G23 | 2372 | Lesotho/ OR Lesotho.mp. |
| G24 | 5259 | Liberia/ OR Liberia.mp. |
| G25 | 13,940 | Madagascar/ OR Madagascar.mp. |
| G26 | 22,145 | Malawi/ OR Malawi.mp. |
| G27 | 11,543 | Mali/ OR Mali.mp. |
| G28 | 1997 | Mauritania/ OR Mauritania.mp. |
| G29 | 2928 | Mauritius/ OR Mauritius.mp. |
| G30 | 11,416 | Mozambique/ OR Mozambique.mp. |
| G31 | 5216 | Namibia/ OR Namibia.mp. |
| G32 | 43,198 | Niger/ OR Niger.mp. |
| G33 | 105,242 | Nigeria/ OR Nigeria.mp. |
| G34 | 17,810 | Republic of the Congo/ OR Republic of the Congo.mp. OR Congo-Brazzaville.mp. |
| G35 | 1860 | Reunion/ |
| G36 | 10,849 | Rwanda/ OR Rwanda.mp. |
| G37 | 488 | "Sao Tome and Principe"/ OR "Sao Tome and Principe".mp. |
| G38 | 19,552 | Senegal/ OR Senegal.mp. |
| G39 | 2027 | Seychelles/ OR Seychelles.mp. |
| G40 | 7333 | Sierra Leone/ OR Sierra Leone.mp. |
| G41 | 6180 | Somalia/ OR Somalia.mp. |
| G42 | 145,351 | South Africa/ OR South Africa.mp. |
| G43 | 1909 | South Sudan/ OR South Sudan.mp. |
| G44 | 26,487 | Sudan/ OR Sudan.mp. |
| G45 | 3261 | Swaziland/ OR Swaziland.mp. OR Eswatini/ OR Eswatini.mp. |
| G46 | 43,118 | Tanzania/ OR Tanzania.mp. |
| G47 | 4746 | Togo/ OR Togo.mp. |
| G48 | 50,642 | Uganda/ OR Uganda.mp. |
| G49 | 17,842 | Zambia/ OR Zambia.mp. |
| G50 | 19,262 | Zimbabwe/ OR Zimbabwe.mp. |
| G51 | 625,159 | exp africa south of the sahara/ OR sub-saharan.mp. OR south of the sahara.mp. |

| | | |
|---|---|---|
| G52 | 1,191,770 | OR/ G1-G51 |
| X1 | 2777 | M8 AND H8 AND G52 |
| X2 | 2740 | X1 NOT animal/ |
| X3 | 2733 | limit X2 to english language |
| X5 | 1740 | remove duplicates from X4 |

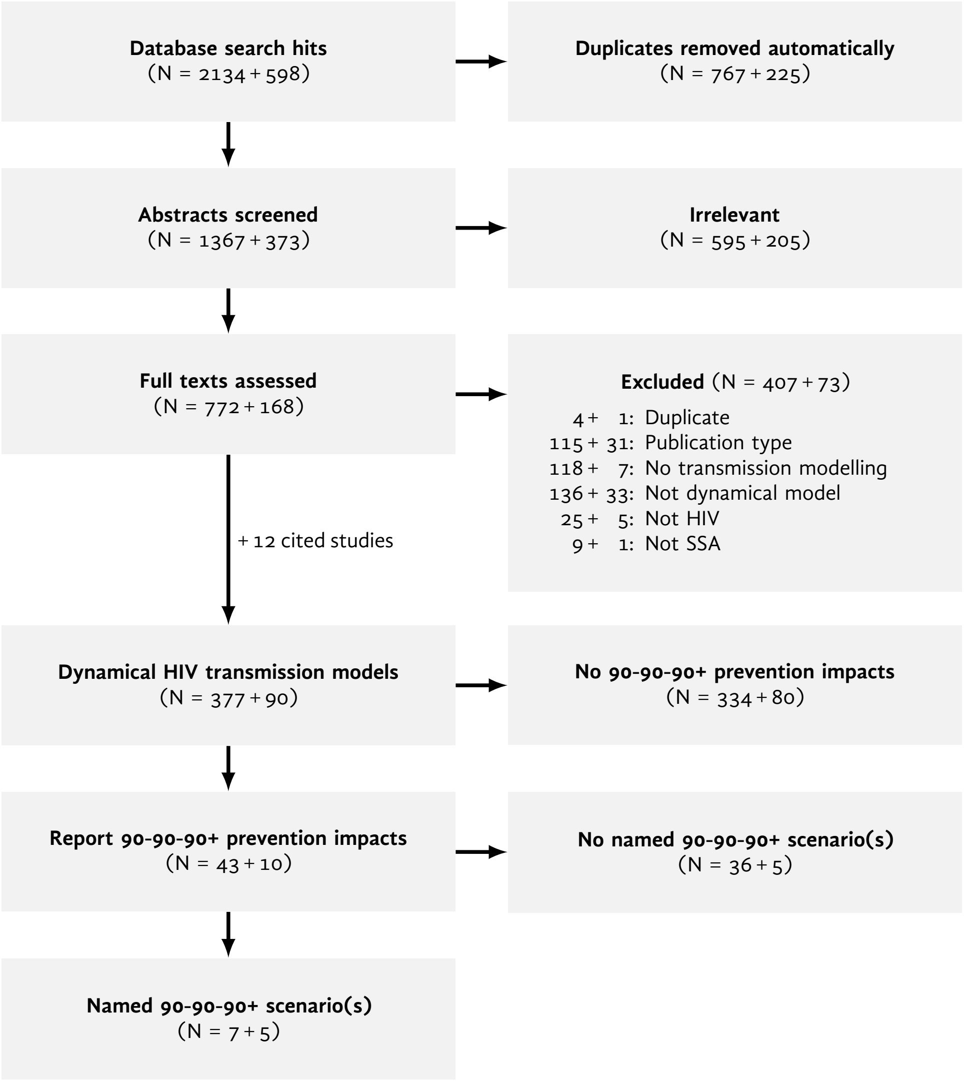

Figure B.22: PRISMA diagram for identifying studies modelling the prevention impacts of 90-90-90+

##### B.3.3 Included Studies

Table B.3 summarizes the characteristics of included studies with named 90-90-90+ scenarios.

Table B.3: Characteristics of studies modelling prevention impacts in named 90-90-90+ scenarios

| | | | Status Quo Rate Diffs [1,2] | | | 90-90-90+ Scenarios | | |
|---|---|---|---|---|---|---|---|---|
| Ref | Model Name | Strata [1] | Diag | Init | Supp | Prioritized | Left Behind | Modelled Context |
| Hontelez et al. [184] | STDSIM | SARK | S | S | — | — | — | Sub-Saharan Africa |
| Maheu-Giroux et al. [185] | — | SARK | SAK | — | K | K | K | Côte d'Ivoire |
| Volz et al. [186] | — | SK | K | K | — | K | — | Nigeria |
| Bansi-Matharu et al. [187] | Synthesis | SARK | SAR | S | — | — | — | Zimbabwe |
| Stuart et al. [188] | Optima | SARK | SAK | SAK | — | — | — | Johannesburg, South Africa |
| Abuelezam et al. [189] | CDM | SARK | — | — | — | SARK | — | South Africa |
| Reidy et al. [190] | Goals | SRK | S | S | — | — | S | Kenya; South Africa; Uganda; Zimbabwe |
| Akullian et al. [191] | EMOD | SARK | SA | SA | — | A | — | Eswatini |
| Marukutira et al. [192] | Optima | SM | SM | SM | M | — | M | Botswana |
| Kripke et al. [193] | Goals | SRK | S | S | — | — | — | Lesotho; Mozambique; Uganda |
| Luong Nguyen et al. [194] | — | R [3] | — | — | — | — | — | Kenya |
| Probert et al. [195] | PopART-IBM | SAR | S | — | S | — | — | Zambia; South Africa |

[1] S: sex; A: age; R: risk; K: any key population(s); M: migrants; [2] Differences in rates of diagnosis, ART initiation, or viral suppression (including treatment failure, discontinuation, loss-to-follow-up);
[3] Risk groups were defined by age/sex.

[7] Swaziland Ministry of Health. *National Guidelines for Antiretroviral Treatment and Post Exposure Prophylaxis*. Mbabane, Swaziland, 2006.

[8] Swaziland Ministry of Health. *The National Guidelines on the Comprehensive HIV Package of Care for Adults and Adolescents in Swaziland*. Mbabane, Swaziland, 2010.

[9] Swaziland Ministry of Health. *Swaziland Integrated HIV Management Guidelines*. Mbabane, Swaziland, 2015.

[10] Swaziland Ministry of Health. *Swaziland Integrated HIV Management Guidelines*. Mbabane, Swaziland, 2018.

[19] PEPFAR. *Characterizing the HIV Prevention and Treatment Needs among Key Populations, including Men who Have Sex with Men and Female Sex Workers in Swaziland: From Evidence to Action*. Mbabane, Swaziland, 2015.

[20] Swaziland Ministry of Health. *Swaziland HIV Incidence Measurement Survey: First Findings Report*. Mbabane, Swaziland, 2012. http://shims.icap.columbia.edu/.

[21] Eswatini Ministry of Health. *Eswatini Population-Based HIV Impact Assessment 3 (SHIMS3): Summary Sheet*. Mbabane, Eswatini, 2022.

[22] Eswatini Ministry of Health. *2020–2021 Integrated Biological-Behavioral Surveillance Survey among Female Sex Workers and Men Who Have sex with Men in Eswatini*. Mbabane, Eswatini, 2022.

[23] Yen T. Duong et al. “Detection of recent HIV-1 infection using a new limiting-antigen avidity assay: Potential for HIV-1 incidence estimates and avidity maturation studies”. *PLOS ONE* 7.3 (2012), e33328. https://doi.org/10.1371/journal.pone.0033328.

[24] Eileen A. Yam et al. “Association between condom use and use of other contraceptive methods among female sex workers in swaziland: A relationship-level analysis of condom and contraceptive use”. *Sexually Transmitted Diseases* 40.5 (2013), pp. 406–412. https://doi.org/10.1097/OLQ.0b013e318283c16d.

[25] Sharon Stucker Weir et al. *Priorities for Local AIDS Control Efforts: A Manual for Implementing the PLACE Method*. 2005. https://www.measureevaluation.org/resources/publications/ms-05-13.html.

[26] Alan Whiteside and Amy Whalley. *Reviewing Emergencies for Swaziland: Shifting the Paradigm in a New Era*. Mbabane, Swaziland: NERCHA, 2007.

[52] PEPFAR. *Eswatini Country Operational Plan 2021*. 2021.

[53] Maria A. Carrasco, Joseph G. Rosen, and Ryan Phelps. “Left behind?: male clients of female sex workers in Zambia”. *AIDS Care* 32.12 (2020), pp. 1498–1505. https://doi.org/10.1080/09540121.2020.1718589.

[54] Julia Cordero-Coma and Richard Breen. “HIV Prevention and Social Desirability: Husband-Wife Discrepancies in Reports of Condom Use”. *Journal of Marriage and Family* 74.3 (2012), pp. 601–613. https://doi.org/10.1111/j.1741-3737.2012.00976.x.

[55] Luc Béhanzin et al. “Assessment of HIV-related risky behaviour: A comparative study of face-to-face interviews and polling booth surveys in the general population of Cotonou, Benin”. *Sexually Transmitted Infections* 89.7 (2013), pp. 595–601. https://doi.org/10.1136/sextrans-2012-050884.

[56] Swaziland Ministry of Health. *Swaziland 1988 Family Health Survey*. Mbabane, Swaziland, 1990.

[57] Swaziland Ministry of Health. *Swaziland Behavioral Surveillance Survey*. Mbabane, Swaziland, 2002.

[58] Anne-Mariea; Marie Côté et al. “Transactional sex is the driving force in the dynamics of HIV in Accra, Ghana”. *AIDS* 18.6 (2004), pp. 917–925. https://doi.org/10.1097/00002030-200404090-00009.

[59] Branwen Nia Owen et al. “What Proportion of Female Sex Workers Practise anal Intercourse and How Frequently? A Systematic Review and Meta-analysis”. *AIDS and Behavior* 24.3 (2020), pp. 697–713. https://doi.org/10.1007/s10461-019-02477-w.

[60] F. N. Fritsch and R. E. Carlson. “Monotone Piecewise Cubic Interpolation”. *SIAM Journal on Numerical Analysis* 17.2 (1980), pp. 238–246. https://doi.org/10.1137/0717021.

[61] National Emergency Response Council on HIV and AIDS. *Swaziland Country Report On Monitoring The Political Declaration On HIV And AIDS*. Mbabane, Swaziland, 2012.

[81] Swaziland Ministry of Health. *Swaziland HIV Incidence Measurement Survey (SHIMS): Descriptive Data Tables*. Mbabane, Swaziland, 2014.

[82] Mathieu Maheu-Giroux et al. “National HIV testing and diagnosis coverage in sub-Saharan Africa: A new modeling tool for estimating the ’first 90’ from program and survey data”. *AIDS* 33 (2019), S255–S269. https://doi.org/10.1097/QAD.0000000000002386.

[83] Government of Swaziland. *Policy Document on HIV/AIDS and STD Prevention and Control*. Mbabane, Swaziland, 1998.

[84] Human Sciences Research Council. *An Audit of HIV/AIDS Policies in Botswana, Lesotho, Mozambique, South Africa, and Swaziland*. Ed. by Nompumelelo Zungu-Dirwayi et al. Cape Town, South Africa: HSRC Publishers, 2004.

[85] Nokwazi Mathabela et al. *UNGASS 2010 Swaziland Progress Report*. Mbabane, Swaziland, 2010.

[86] National Emergency Response Council on HIV and AIDS. *The Kingdom of Swaziland National AIDS Spending Assessment 2005/06 and 2006/07*. Mbabane, Swaziland, 2009.

[87] National Emergency Response Council on HIV and AIDS. *National Multi-sectorial Strategic Framework for HIV and AIDS 2009-2014*. Mbabane, Swaziland, 2009.

[88] Françoise Jenniskens. *Maximizing ART for Better Health and Zero New HIV Infections*. Mbabane, Eswatini, 2015.

[89] WHO. *Scaling up antiretroviral therapy in resource-limited settings : treatment guidelines for a public health approach*. Geneva, Switzerland, 2013.

[90] WHO. *Chronic HIV Care with ARV Therapy and Prevention*. Geneva, Switzerland, 2007.

[91] WHO. *Consolidated guidelines on the use of antiretroviral drugs for treating and preventing HIV infection*. Geneva, Switzerland, 2013.

[98] Swaziland Ministry of Health. *Annual HIV Programs Report*. Mbabane, Swaziland, 2015.

[99] Kathryn E. Lancaster et al. “HIV care and treatment experiences among female sex workers living with HIV in sub-Saharan Africa: A systematic review”. *African Journal of AIDS Research* 15.4 (2016), pp. 377–386. https://doi.org/10.2989/16085906.2016.1255652.

[100] WHO. *HIV Drug Resistance Report 2021*. Geneva, Switzerland, 2021.

[101] Anthony Hauser et al. “Bridging the gap between hiv epidemiology and antiretroviral resistance evolution: Modelling the spread of resistance in south africa”. *PLOS Computational Biology* 15.6 (2019), e1007083. https://doi.org/10.1371/journal.pcbi.1007083.

[102] National Emergency Response Council on HIV and AIDS. *Swaziland Global Aids Response Progress Reporting 2014*. 2014.

[129] UNAIDS/WHO Working Group on Global HIV/AIDS and STI Surveillance. *Guidelines on Estimating the Size of Populations Most at Risk to HIV*. Geneva, Switzerland, 2010.

[130] Abu S Abdul-Quader, Andrew L Baughman, and Wolfgang Hladik. "Estimating the size of key populations: Current status and future possibilities". *Current Opinion in HIV and AIDS* 9.2 (2014), pp. 107–114. http://doi.org/10.1097/COH.0000000000000041.

[131] L. Morison et al. "Commercial sex and the spread of HIV in four cities in sub-Saharan Africa". *AIDS* 15.S4 (2001), S61–S69. https://doi.org/10.1097/00002030-200108004-00007.

[132] World Health Organization. *Swaziland, Mortality Country Fact Sheet 2006*. 2006.

[138] Roy M Anderson and Robert M May. *Infectious diseases of humans: dynamics and control*. Oxford University Press, 1991.

[152] Olive Shisana and Leickness Simbayi. *South African national HIV prevalence, HIV incidence, behaviour and communication survey, 2005*. Cape Town, South Africa, 2005.

[178] PEPFAR. *HIV among Female Sex Workers and Men Who Have Sex with Men in Swaziland*. Mbabane, Swaziland, 2014.
